# Health and economic impacts of Lassa vaccination campaigns in West Africa

**DOI:** 10.1101/2024.02.26.24303394

**Authors:** David R M Smith, Joanne Turner, Patrick Fahr, Lauren A Attfield, Paul R Bessell, Christl A Donnelly, Rory Gibb, Kate E Jones, David W Redding, Danny Asogun, Oladele Oluwafemi Ayodeji, Benedict N Azuogu, William A Fischer, Kamji Jan, Adebola T Olayinka, David A Wohl, Andrew A Torkelson, Katelyn A Dinkel, Emily J Nixon, Koen B Pouwels, T Déirdre Hollingsworth

## Abstract

Lassa fever is a zoonotic disease identified by the World Health Organization (WHO) as having pandemic potential. This study estimates the health-economic burden of Lassa fever throughout West Africa and projects impacts of a series of vaccination campaigns. We also model the emergence of “Lassa-X” – a hypothetical pandemic Lassa virus variant – and project impacts of achieving 100 Days Mission vaccination targets. Our model predicted 2.7M (95% uncertainty interval: 2.1M-3.4M) Lassa virus infections annually, resulting over ten years in 2.0M (793.8K-3.9M) disability-adjusted life years (DALYs). The most effective vaccination strategy was a population-wide preventive campaign primarily targeting WHO-classified “endemic” districts. Under conservative vaccine efficacy assumptions, this campaign averted $20.1M ($8.2M-$39.0M) in lost DALY value and $128.2M ($67.2M-$231.9M) in societal costs (International dollars 2021). Reactive vaccination in response to local outbreaks averted just one-tenth the health-economic burden of preventive campaigns. In the event of Lassa-X emerging, spreading throughout West Africa and causing approximately 1.2M DALYs within two years, 100 Days Mission vaccination averted 22% of DALYs given a vaccine 70% effective against disease, and 74% of DALYs given a vaccine 70% effective against both infection and disease. These findings suggest how vaccination could alleviate Lassa fever’s burden and assist in pandemic preparedness.

## Main

Lassa fever is a viral haemorrhagic disease endemic to West Africa, where infections are common but widely undetected. Lassa fever is caused by *Lassa mammarenavirus* (LASV) and several lines of evidence, including detailed genomic analyses, suggest that the vast majority of human LASV infections are caused by zoonotic transmission from the Natal multimammate mouse (*Mastomys natalensis*).^1,2^ The virus can also spread through human-to-human contact, although this has predominantly been observed in healthcare settings with inadequate infection prevention and control practices.^3^

Most LASV infections are believed to be asymptomatic or cause only mild febrile illness,^4^ but Lassa fever nonetheless has a large negative impact on population health and economies. Among patients presenting to hospital, the case-fatality ratio is estimated to be around 15%, and long-term sequelae such as bilateral sensorineural hearing loss are common in Lassa fever survivors.^5,6^ Monetary costs per hospitalisation are estimated to be high and are often paid (partly) out-of-pocket by patients.^7^ For example, a study from Nigeria found that the average patient’s out-of-pocket expenditure on Lassa fever treatment was approximately 480% of the monthly minimum wage in 2011.^8^

No licensed vaccines against Lassa fever are currently available, although several candidates are under development. A recent phase 1 randomised trial of a measles-vectored Lassa vaccine showed an acceptable safety and tolerability profile, a substantial increase in LASV-specific non-neutralising IgG concentrations and a moderate T-cell response,^9^ in line with the response observed in non-human primates.^10^ Several other vaccines are currently at early stages of development, with five phase 1 trials and one phase 2 trial registered by October 2022.^11^

Lassa fever is listed by the World Health Organization (WHO) as one of the diseases posing the greatest risk to public health due to its epidemic potential and the absence of effective countermeasures.^12^ In response to such concerns, in 2022 the Group of Seven forum, Group of Twenty forum and various international governments endorsed the 100 Days Mission, a pandemic response roadmap aiming at the delivery of vaccines within 100 days of the emergence of novel pathogens with pandemic potential.^13^

In anticipation of one or more Lassa vaccine candidates being licensed in the near future, in this study we estimate the current health-economic burden of Lassa fever in West Africa and project the potential impacts of different reactive and preventive vaccination campaigns. We also project potential impacts of vaccination in line with the 100 Days Mission in response to the emergence of “Lassa-X”, a hypothetical future variant of LASV with pandemic potential. **Table 1** summarises our main findings and their implications for public policy.

**Table 1.**
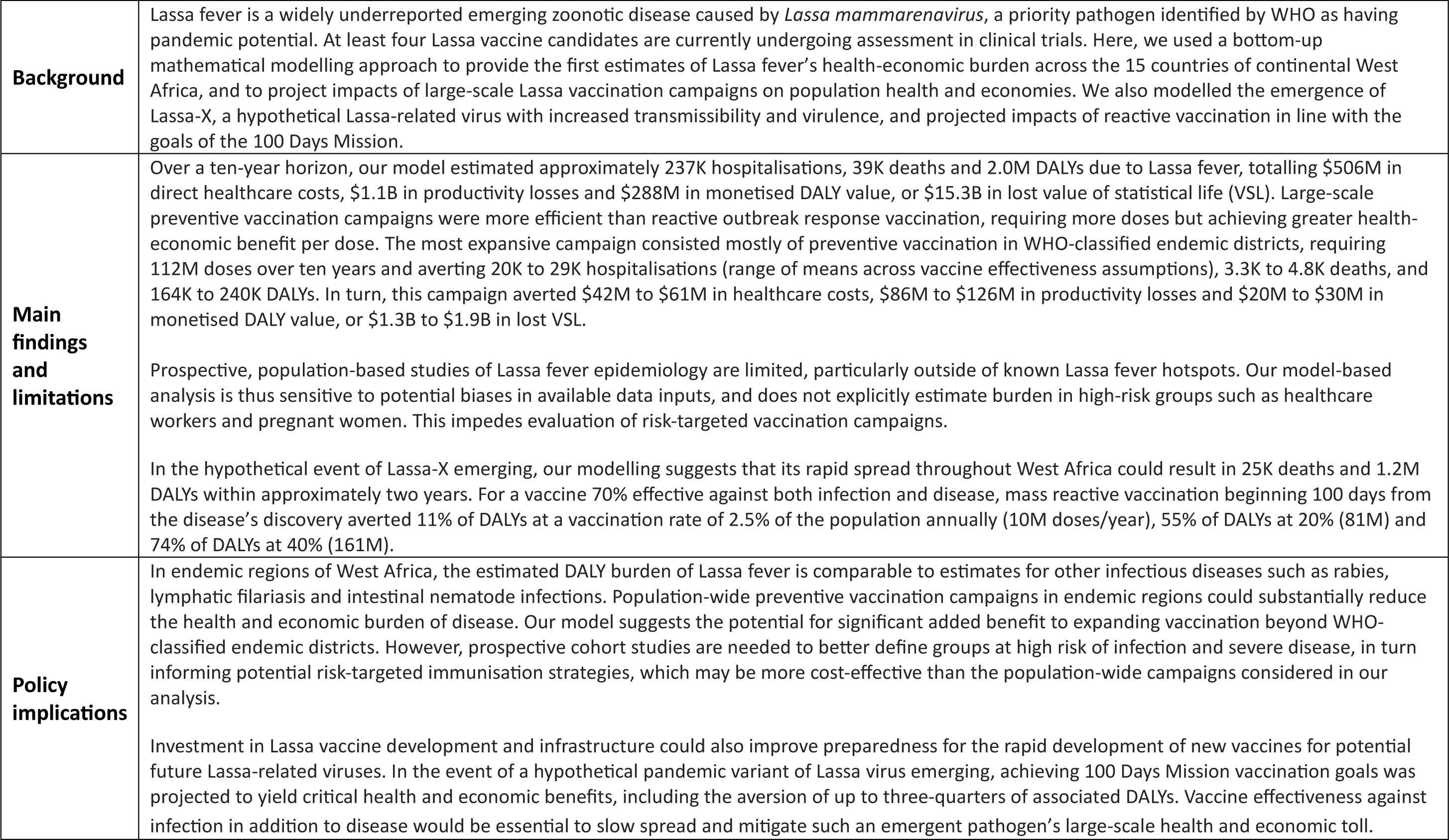
Policy summary. Monetary costs are reported in International dollars 2021. DALY = disability-adjusted life year, WHO = World Health Organization, VSL = value of statistical life, K = thousand, M = million, B = billion.

## Results

### Model overview

We developed an epidemiological model projecting human Lassa fever burden over a 10-year time horizon across the 15 countries of continental West Africa (Benin, Burkina Faso, Côte d’Ivoire, The Gambia, Ghana, Guinea, Guinea Bissau, Liberia, Mali, Mauritania, Niger, Nigeria, Senegal, Sierra Leone and Togo) and their 183 Level 1 sub-national administrative units. These units have different names in different countries (e.g. regions in Guinea, counties in Liberia, departments in Benin) but herein are collectively referred to as *districts*. Due to large gaps in Lassa fever surveillance and limited case reporting throughout much of its endemic range,^3^ we favoured a bottom-up modelling approach, synthesizing best available ecological, epidemiological, clinical and economic data to project the cumulative health and economic burden of disease.

Our model consists of six main components (see model schematic in **Extended Data Figure 1**). First, a previously published geospatial risk map is used to predict the risk of zoonotic LASV transmission from *M. natalensis* to humans (“spillover”) at the level of 0.05° x 0.05° spatial pixels throughout West Africa.^14^ Second, modelled spillover risk estimates are used as inputs in a generalised linear model (GLM) to predict human LASV seroprevalence. Third, modelled human LASV seroprevalence estimates are used as inputs in a serocatalytic model including country-level population projections to predict spillover infection incidence. Fourth, spillover infections are aggregated at the district level, and a stochastic branching process model is used to simulate onward human-to-human LASV transmission. Fifth, a computational algorithm is applied retrospectively to spillover infections and ensuing transmission chains to simulate a range of reactive and preventive vaccination campaigns and to project the number of infections averted by vaccination. (Separate model components used to simulate Lassa-X transmission and vaccination are described below.) Sixth, modelled estimates of LASV infection, and of infections averted due to vaccination or occurring in vaccinated individuals, are used as inputs in a probabilistic decision-analytic model used to project the health burden of Lassa fever and associated economic costs, and the health and economic burden averted due to vaccination over ten years.

### Lassa fever burden

Our model predicts a heterogeneous distribution of zoonotic LASV infection throughout West Africa (**Figure 1**). In the absence of vaccination, the mean annual number of LASV infections throughout the region was estimated at 2.7M (95%UI: 2.1M–3.4M), or 27.2M (20.9M–34.0M) over the full 10-year simulation period (**Extended Data Table 1**). Just over half of all infections occurred in Nigeria (mean 52.9%) and the vast majority (mean 93.7%) resulted from zoonotic spillover as opposed to human-to-human transmission, due to LASV’s low estimated basic reproduction number (*R*0). At the district level, annual LASV infection incidence was highest in Margibi, Liberia (1,198 [943–1,475] infections /100,000 population), followed by Denguélé, Côte d’Ivoire (1,032 [880–1,200] /100,000) and Nasarawa, Nigeria (978 [803–1,162] /100,000). Over ten years, LASV infection throughout West Africa led to an estimated 5.4M (2.7M–9.9M) mild/moderate symptomatic cases, 237.0K (148.6K–345.6K) hospitalisations and 39.3K (12.9K–83.3K) deaths, resulting in 2.0M (793.8K-3.9M) disability-adjusted life years (DALYs). See **Supplementary Appendix E** for more detailed estimates of Lassa fever burden.

**Figure 1.**
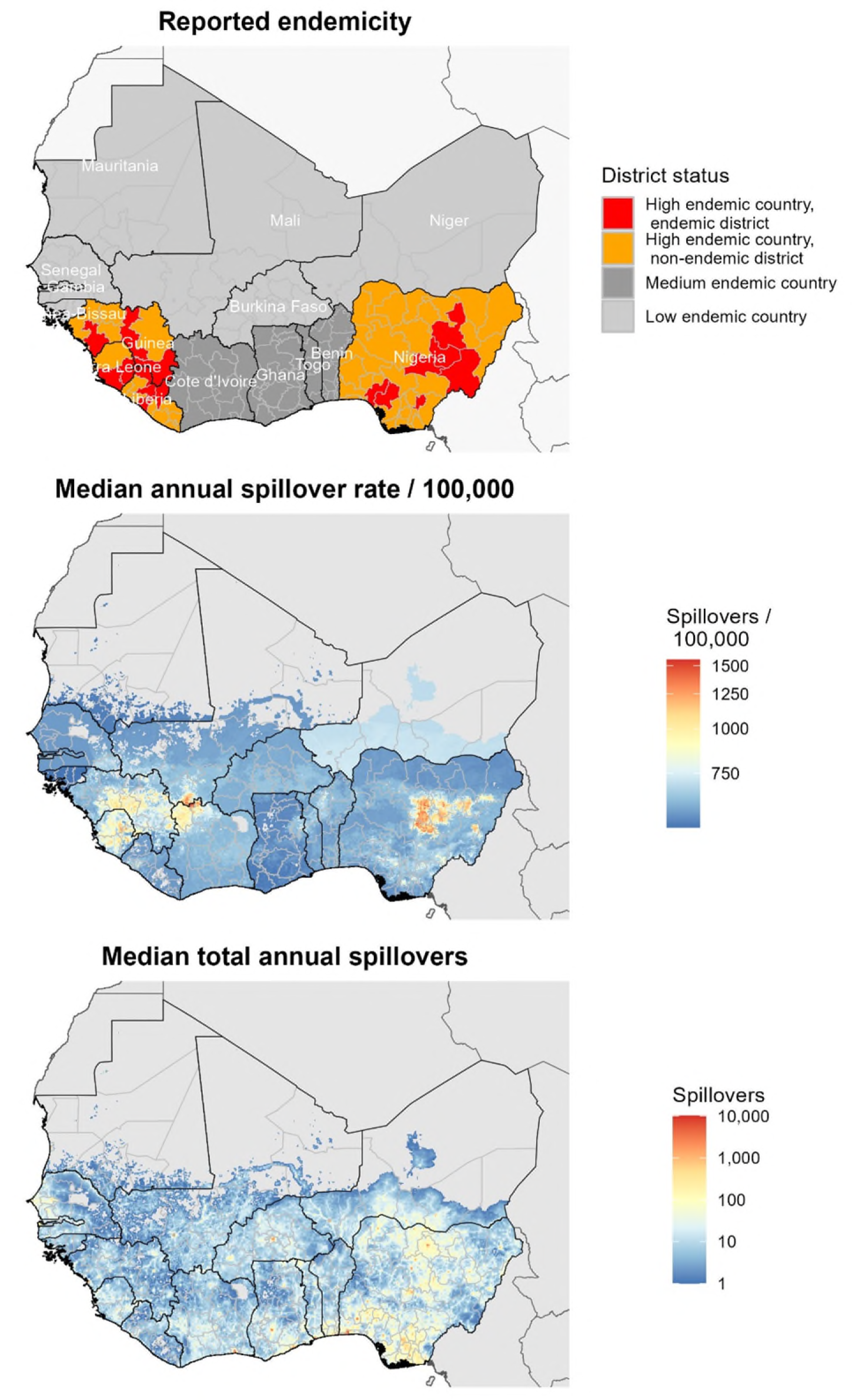
Maps of West Africa showing reported Lassa fever endemicity and estimated LASV spillover incidence. Top: map showing the classification of Lassa fever endemicity for different countries and “districts”, as defined by US CDC and WHO (see **Supplementary appendix C.2**). Middle: the median annual incidence of zoonotic LASV infection per 100,000 population as estimated by our model at the level of 5km grid cells. Bottom: the median total annual number of zoonotic LASV infections as estimated by our model at the level of 5km grid cells. LASV: Lassa virus, US CDC: United States Centers for Disease Control & Prevention, WHO: World Health Organization.

Over ten years, Lassa fever treatment was projected to incur $338.9M ($206.6M-$506.3M) in government-reimbursed treatment costs and $166.9M ($116.0M-$289.3M) in out-of-pocket medical costs, resulting in catastrophic expenditures for 232.3K (145.6K-338.7K) individuals and pushing 167.0K (104.7K-243.6K) individuals below the international poverty line (**Supplementary tables E.3 and E.4**). Missed work due to illness totalled $1.1B ($380.5M-$2.2B) in productivity losses, primarily due to mortality in actively employed adults. Productivity losses outranked treatment costs in driving an estimated $1.6B ($805.1M-$2.8B) in total cumulative societal costs. Hospitalisation costs, not outpatient costs, were the main driver of treatment costs, but mild to moderate disease in the community resulted in greater productivity losses than severe disease in hospital (**Supplementary figure E.2**). Lassa fever DALYs were valued at $287.7M ($115.4M-$562.9M) using country-specific cost-effectiveness thresholds. Finally, an alternative measure of Lassa fever’s economic burden, the value of statistical life (VSL) lost due to Lassa fever mortality, was projected at $15.3B ($5.0B–$32.4B). Uncertainty in health-economic outcomes was primarily driven by uncertainty in risks of hospitalisation and death (**Supplementary figure D.2**)

### Simulating Lassa vaccination campaigns

Vaccination is introduced into the population via a series of six scenarios designed to reflect realistic assumptions about vaccine stockpile, administration and efficacy (**Extended Data Table 2**). In all six scenarios we include reactive vaccination, in which Lassa fever outbreaks trigger the local deployment of a limited vaccine stockpile in affected districts. In scenarios 2 through 6 we also include preventive vaccination in the form of mass, population-wide campaigns rolled out over three years and focusing primarily on regions classified as Lassa fever “endemic”. The 15 countries included in our model are categorised as high endemic, medium endemic or low endemic according to classifications published by the US CDC, while districts within high-endemic countries are further classified as endemic or non-endemic according to classifications published by WHO (see **Figure 1** and **Supplementary appendix C.2**). Two main mechanisms of vaccine efficacy are considered: protection against infection prevents individuals from acquiring LASV infection from either *M. natalensis* or other humans, and protection against disease prevents vaccinated individuals who become infected from progressing to disease, thus averting outpatient consultation, hospitalisation, chronic sequelae and death. In our simulations we project impacts of a vaccine that is 70% or 90% effective only against disease, or 70% or 90% effective against both infection and disease. We do not consider other potential mechanistic impacts of vaccination, such as reduced infectiousness or altered behaviour among vaccinated individuals, as such factors are less relevant given low estimated rates of human-to-human LASV transmission.

### Health-economic impacts of vaccination against Lassa fever

The considered vaccination scenarios varied considerably in their projected impacts, with scenario 4 leading to the greatest reductions in Lassa fever burden over ten years (**Extended Data Figure 2** and **Table 2**). In this scenario, in addition to reactive vaccination triggered in districts experiencing local outbreaks, preventive vaccination was administered to 80% of the population in WHO-classified endemic districts, as well as to 5% of the population in all other districts throughout West Africa. For a vaccine 70% effective against disease with no impact on infection, over ten years this strategy averted a mean 456.0K (226.4K-822.7K) mild/moderate symptomatic cases, 19.9K (12.7K-28.8K) hospitalisations, 3.3K (1.1K-7.0K) deaths and 164.1K (66.7K-317.7K) DALYs. Over this period, this strategy further prevented 19.8K (12.6K-28.5K) and 14.2K (9.0K-20.5K) individuals, respectively, from experiencing catastrophic or impoverishing out-of-pocket healthcare expenditures, and averted $128.2M ($67.2M-$231.9M) in societal costs, or $1.3B ($436.8M-$2.8B) in VSL lost.

Other vaccination scenarios used fewer doses of vaccine and, in turn, averted less of Lassa fever’s health-economic burden. Scenario 3, which limited preventive vaccination to high-endemic countries, was the scenario resulting in the second greatest health-economic benefits, including the aversion of 141.4K (57.6K-273.2K) DALYs and $112.8M ($59.2M-$203.8M) in societal costs. Scenarios 2, 5 and 6 varied considerably in terms of which individuals were vaccinated but ultimately resulted in similar cumulative health-economic benefits across the region, because the overall number of doses delivered under each scenario was essentially the same. By contrast, scenario 1 included only reactive and not preventive vaccination, averting just 13.7K (5.5K-26.8K) DALYs and $10.3M ($5.3M–$18.8M) in societal costs, thus having approximately one-tenth the overall health-economic benefits of scenario 4.

A vaccine effective against infection in addition to disease was found to have moderately increased impact. In scenario 4 for instance, $20.1M ($8.2M-$39.0M) in DALY value was averted by a vaccine 70% effective only against disease, while $27.1M ($11.0M-$52.5M) was averted when also 70% effective against infection (**Table 2**). By comparison, a vaccine 90% effective only against disease averted $25.8M ($10.5M-$50.1M) in DALY value (**Supplementary table E.9**), having similar impact to a vaccine 70% effective against both infection and disease. In the best-case scenario of a vaccine 90% effective against both infection and disease, scenario 4 averted up to 3.1M (2.4M-3.7M) infections, 240.1K (97.5K-464.9K) DALYs valued at $29.5M ($12.0M-$57.2M), and $1.9B ($638.5M-$4.1B) in VSL lost.

**Table 2.**
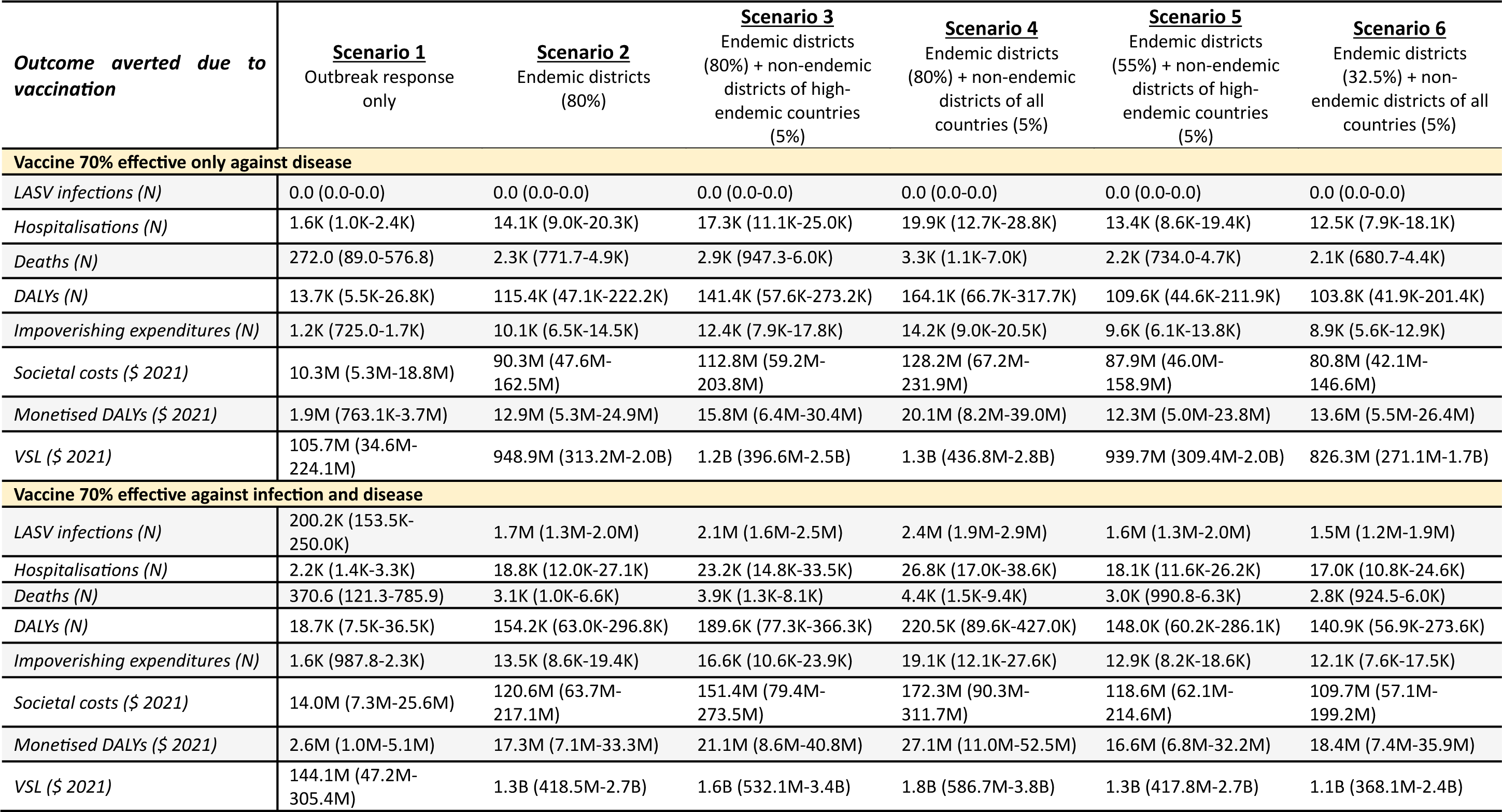
Projected ten-year impacts of Lassa vaccination. The mean (95% uncertainty interval) health and economic burden of Lassa fever averted due to vaccination over ten years from the initiation of vaccine rollout, for the six vaccination scenarios described in **Extended Data Table 2**. Columns represent vaccination scenarios and rows represent outcomes averted. The table compares a vaccine 70% effective only against disease (top) with a vaccine 70% effective against both infection and disease (bottom). Societal costs combine outpatient treatment costs, hospital treatment costs and productivity losses. Costs are reported in International dollars (2021) and future monetary costs are discounted at 3%/year. LASV = Lassa virus, DALY = disability-adjusted life year, VSL = value of statistical life, K = thousand, M = million, B = billion.

| **Outcome averted due to vaccination** | **Scenario 1**<br>Outbreak response only | **Scenario 2**<br>Endemic districts (80%) | **Scenario 3**<br>Endemic districts (80%) + non-endemic districts of high-endemic countries (5%) | **Scenario 4**<br>Endemic districts (80%) + non-endemic districts of all countries (5%) | **Scenario 5**<br>Endemic districts (55%) + non-endemic districts of high-endemic countries (5%) | **Scenario 6**<br>Endemic districts (32.5%) + non-endemic districts of all countries (5%) |
| --- | --- | --- | --- | --- | --- | --- |
| **Vaccine 70% effective only against disease** |  |  |  |  |  |  |
| *LASV infections (N)* | 0.0 (0.0-0.0) | 0.0 (0.0-0.0) | 0.0 (0.0-0.0) | 0.0 (0.0-0.0) | 0.0 (0.0-0.0) | 0.0 (0.0-0.0) |
| *Hospitalisations (N)* | 1.6K (1.0K-2.4K) | 14.1K (9.0K-20.3K) | 17.3K (11.1K-25.0K) | 19.9K (12.7K-28.8K) | 13.4K (8.6K-19.4K) | 12.5K (7.9K-18.1K) |
| *Deaths (N)* | 272.0 (89.0-576.8) | 2.3K (771.7-4.9K) | 2.9K (947.3-6.0K) | 3.3K (1.1K-7.0K) | 2.2K (734.0-4.7K) | 2.1K (680.7-4.4K) |
| *DALYs (N)* | 13.7K (5.5K-26.8K) | 115.4K (47.1K-222.2K) | 141.4K (57.6K-273.2K) | 164.1K (66.7K-317.7K) | 109.6K (44.6K-211.9K) | 103.8K (41.9K-201.4K) |
| *Impoverishing expenditures (N)* | 1.2K (725.0-1.7K) | 10.1K (6.5K-14.5K) | 12.4K (7.9K-17.8K) | 14.2K (9.0K-20.5K) | 9.6K (6.1K-13.8K) | 8.9K (5.6K-12.9K) |
| *Societal costs (\$ 2021)* | 10.3M (5.3M-18.8M) | 90.3M (47.6M-162.5M) | 112.8M (59.2M-203.8M) | 128.2M (67.2M-231.9M) | 87.9M (46.0M-158.9M) | 80.8M (42.1M-146.6M) |
| *Monetised DALYs (\$ 2021)* | 1.9M (763.1K-3.7M) | 12.9M (5.3M-24.9M) | 15.8M (6.4M-30.4M) | 20.1M (8.2M-39.0M) | 12.3M (5.0M-23.8M) | 13.6M (5.5M-26.4M) |
| *VSL (\$ 2021)* | 105.7M (34.6M-224.1M) | 948.9M (313.2M-2.0B) | 1.2B (396.6M-2.5B) | 1.3B (436.8M-2.8B) | 939.7M (309.4M-2.0B) | 826.3M (271.1M-1.7B) |
| **Vaccine 70% effective against infection and disease** |  |  |  |  |  |  |
| *LASV infections (N)* | 200.2K (153.5K-250.0K) | 1.7M (1.3M-2.0M) | 2.1M (1.6M-2.5M) | 2.4M (1.9M-2.9M) | 1.6M (1.3M-2.0M) | 1.5M (1.2M-1.9M) |
| *Hospitalisations (N)* | 2.2K (1.4K-3.3K) | 18.8K (12.0K-27.1K) | 23.2K (14.8K-33.5K) | 26.8K (17.0K-38.6K) | 18.1K (11.6K-26.2K) | 17.0K (10.8K-24.6K) |
| *Deaths (N)* | 370.6 (121.3-785.9) | 3.1K (1.0K-6.6K) | 3.9K (1.3K-8.1K) | 4.4K (1.5K-9.4K) | 3.0K (990.8-6.3K) | 2.8K (924.5-6.0K) |
| *DALYs (N)* | 18.7K (7.5K-36.5K) | 154.2K (63.0K-296.8K) | 189.6K (77.3K-366.3K) | 220.5K (89.6K-427.0K) | 148.0K (60.2K-286.1K) | 140.9K (56.9K-273.6K) |
| *Impoverishing expenditures (N)* | 1.6K (987.8-2.3K) | 13.5K (8.6K-19.4K) | 16.6K (10.6K-23.9K) | 19.1K (12.1K-27.6K) | 12.9K (8.2K-18.6K) | 12.1K (7.6K-17.5K) |
| *Societal costs (\$ 2021)* | 14.0M (7.3M-25.6M) | 120.6M (63.7M-217.1M) | 151.4M (79.4M-273.5M) | 172.3M (90.3M-311.7M) | 118.6M (62.1M-214.6M) | 109.7M (57.1M-199.2M) |
| *Monetised DALYs (\$ 2021)* | 2.6M (1.0M-5.1M) | 17.3M (7.1M-33.3M) | 21.1M (8.6M-40.8M) | 27.1M (11.0M-52.5M) | 16.6M (6.8M-32.2M) | 18.4M (7.4M-35.9M) |
| *VSL (\$ 2021)* | 144.1M (47.2M-305.4M) | 1.3B (418.5M-2.7B) | 1.6B (532.1M-3.4B) | 1.8B (586.7M-3.8B) | 1.3B (417.8M-2.7B) | 1.1B (368.1M-2.4B) |

Geographic variation in vaccine impact depended primarily on which districts were classified as endemic and hence targeted for vaccination (**Extended Data Figure 2**). Overall impacts of vaccination were greatest in Nigeria, but impacts per 100,000 population were greatest in other endemic countries (Guinea, Liberia and Sierra Leone), because Nigeria had a larger number of individuals but a smaller share of its total population living in districts classified as endemic. In turn, approximately 16% of the total population of Nigeria and 33% of the combined population of Guinea, Liberia and Sierra Leone were vaccinated by ten years under scenarios 3 and 4 (**Figure 2**). Given a vaccine 70% effective only against disease, these scenarios averted 10.5% of DALYs in Nigeria, 20.3% in Liberia, 23.6% in Guinea and 28.1% in Sierra Leone. For a vaccine 90% effective against infection and disease, these scenarios averted 15.3% of DALYS in Nigeria, 29.4% in Liberia, 34.1% in Guinea and 40.7% in Sierra Leone.

**Figure 2.**
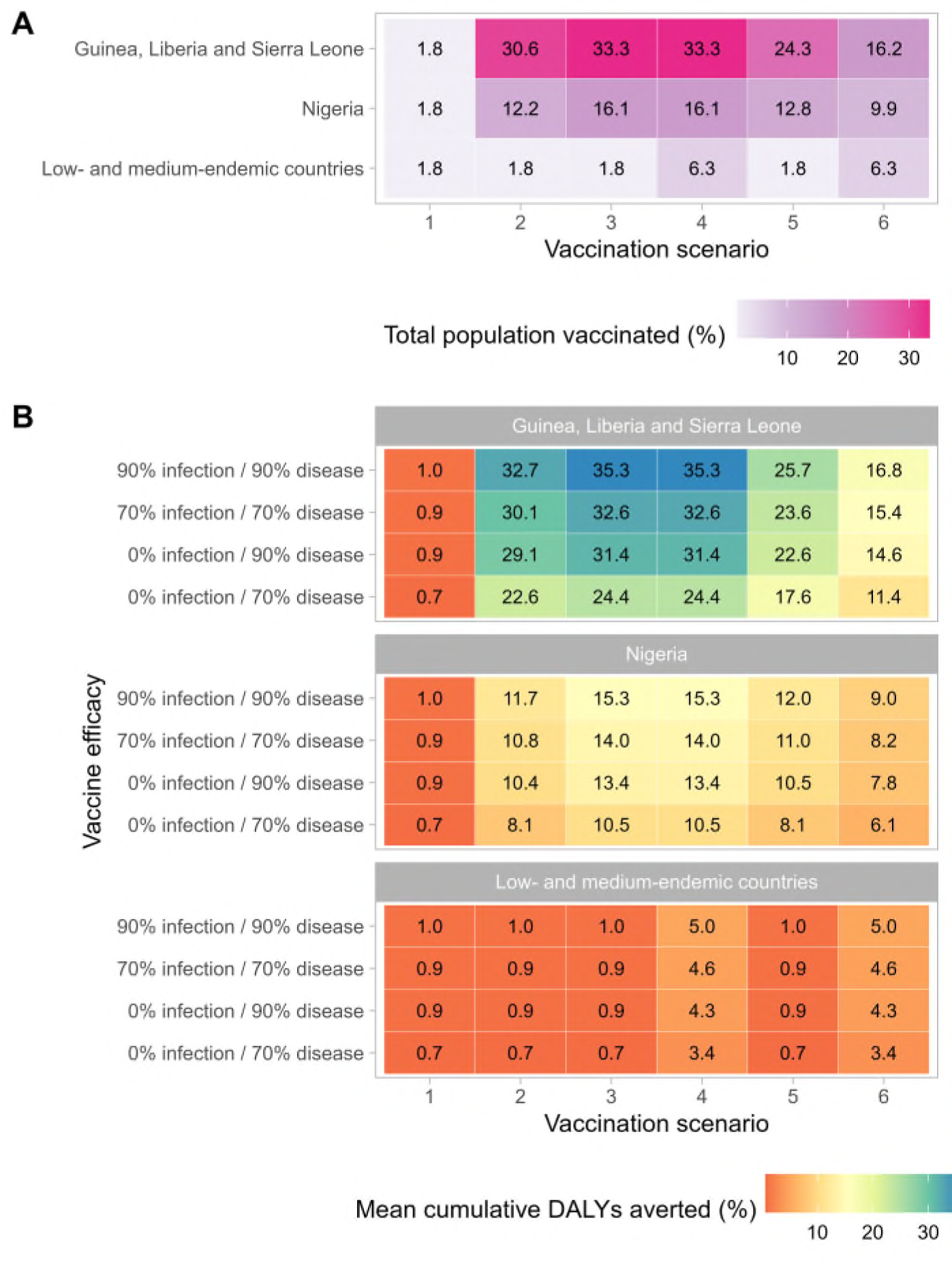
Vaccination coverage and corresponding reductions in Lassa fever burden vary greatly across countries. (**A**) The share of the total population vaccinated by ten years in each vaccination scenario (x-axis) and aggregated across three geographic levels (y-axis). (**B**) The share of cumulative DALYs due to Lassa fever averted over ten years by vaccination. Impacts vary greatly depending on the vaccination scenario (x-axis), assumed vaccine efficacy (y-axis) and the geographic location (panels). DALY: disability-adjusted life year.

### Threshold vaccine costs

Projected economic benefits of Lassa vaccination were used to calculate the threshold vaccine cost (TVC). This can be interpreted as the maximum cost per dose at which vaccination has a benefit-to-cost ratio above one, in the specific context of our modelled vaccination campaigns and corresponding dosage assumptions (i.e. a single-dose primary series followed by a single-dose booster after five years, with 10% dose wastage). TVCs were similar across all five preventive campaigns (scenarios 2 through 6), but lower for reactive vaccination (scenario 1) (**Supplementary table E.12**). Estimated TVCs ranged from $0.51 ($0.30-$0.80) to $21.15 ($7.28-$43.97) depending on the economic perspective considered, the vaccination campaign evaluated and the vaccine’s efficacy against infection and disease. TVCs were lowest from the perspective considering only healthcare costs and monetised DALYs (range of means: $0.51-$0.91), but more than doubled given a perspective considering all societal costs (healthcare costs and productivity losses) in addition to monetised DALYs ($1.18-$2.20), and increased by more than twenty-fold when considering healthcare costs and VSL ($10.54-$21.15).

### Modelling Lassa-X

In addition to our analysis of Lassa fever, we model the emergence of “Lassa-X”, a hypothetical future variant of LASV with pandemic potential due to both elevated clinical severity and increased propensity for human-to-human transmission. In this analysis, Lassa-X was assumed to emerge in humans following a single spillover event, where the probability of emergence in each district is directly proportional to the estimated share of all zoonotic LASV infections occurring in each district. We assumed that prior LASV immunity, whether natural or vaccine-derived, offers no protection against Lassa-X. We conceptualised Lassa-X as having Ebola-like transmission characteristics and, under baseline assumptions, a ten-fold increase in hospitalisation risk relative to Lassa fever. Lassa-X transmission parameters were quantified using Ebola case data from the 2013/16 West Africa epidemic, resulting in simulated Lassa-X outbreaks lasting for approximately two years before subsiding. A range of reactive 100 Days Mission vaccination scenarios were then evaluated, considering different delays to vaccine initiation, rates of vaccine uptake, and degrees of efficacy against infection and disease. Finally, as for Lassa fever, we used a probabilistic decision-analytic model to project the health and economic burden of Lassa-X, and burden averted as a result of vaccination.

### Projected burden of Lassa-X

Under our modelling assumptions, the emergence of Lassa-X led to explosive outbreaks throughout West Africa (**Figure 3**), spreading to 88.3% (63.9%–94.0%) of the 183 districts included in our model (**Supplementary figure F.1**). In total, there were 1.7M (230.1K–4.2M) Lassa-X infections, and Nigeria accounted for by far the greatest share of infections, followed by Niger and Ghana (**Supplementary tables G.1 and G.2**). The projected burden of Lassa-X infection was associated with a high degree of uncertainty, driven predominantly by the highly stochastic nature of simulated outbreaks (**Supplementary figure G.2**).

**Figure 3.**
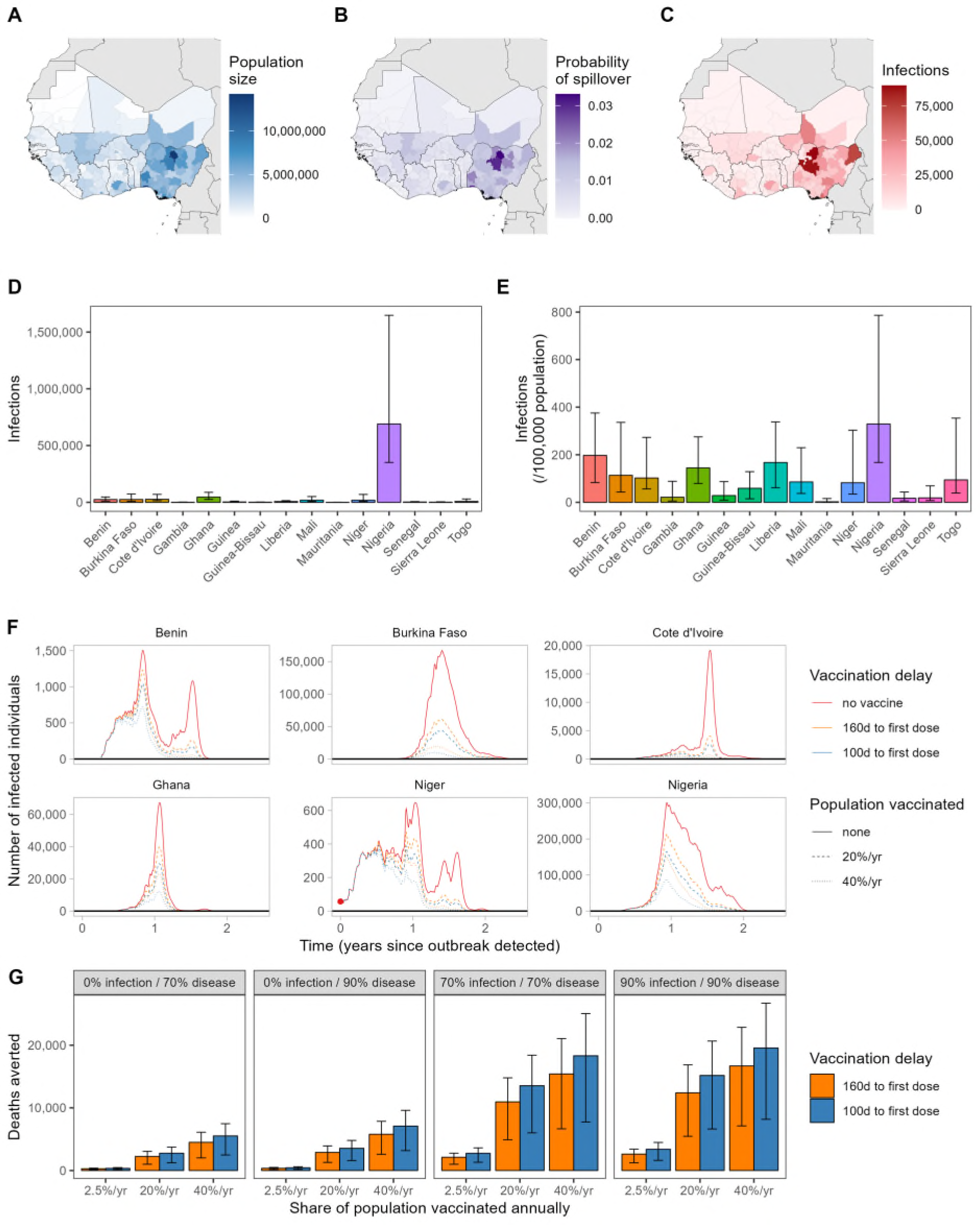
Projected burden of Lassa-X infection and impacts of vaccination. Maps of West Africa showing, for each district: (**A**) the population size, (**B**) the probability of Lassa-X spillover and (**C**) the mean cumulative number of Lassa-X infections over the entire outbreak (approximately two years). The second row depicts (**D**) the median cumulative incidence of Lassa-X infection over the entire outbreak and (**E**) the median cumulative incidence over the entire outbreak per 100,000 population in the absence of vaccination. Interquartile ranges are indicated by error bars. (**F**) The total number of Lassa-X infections over time in six selected countries in one randomly selected outbreak simulation in which the initial Lassa-X spillover event occurred in Niger (the red dot highlights the initial detection of the epidemic at time 0). Lines show how a vaccine with 70% efficacy against infection and disease influences infection dynamics, where line colour represents the delay to vaccine rollout and line dashing represents the rate of vaccination (the proportion of the population vaccinated over a 1-year period). (**G**) The mean cumulative number of deaths averted due to vaccination over the entire outbreak and across all countries, depending on vaccine efficacy (panels), the rate of vaccination (x-axis) and the delay to vaccine rollout (colours). Interquartile ranges are indicated by error bars.

In our baseline analysis, Lassa-X resulted in 149.7K (19.7K–374.4K) hospitalisations and 24.8K (2.4K-76.0K) deaths, causing 1.2M (95% UI: 132.5K–3.7M) DALYs valued at $191.1M ($18.4M–$575.2M). Out-of-pocket treatment costs were estimated at $118.5M ($12.2M–$317.3M), resulting in catastrophic healthcare expenditures for 147.4K (18.5K-372.5K) individuals, and pushing 103.1K (13.6K-254.3K) individuals below the poverty line. Lassa-X also resulted in $737.2M ($56.4M-$2.4B) in productivity losses to the greater economy and $10.1B ($625.9M-34.1B) in VSL lost. In alternative scenarios where Lassa-X infection was just as likely or one-tenth as likely to result in hospitalisation as LASV infection, estimates of the health-economic burden were approximately one and two orders of magnitude lower, respectively (**Supplementary table G.4**).

### Vaccination to slow the spread of Lassa-X

Impacts of vaccination on the health-economic burden of Lassa-X depend on the delay until vaccination initiation, the rate of vaccine uptake in the population, and the efficacy of vaccination against infection and/or disease (**Table 3**). In the most ambitious vaccination scenario considered, vaccine administration began 100 days after initial detection of the first hospitalised case of Lassa-X at a rate equivalent to 40% of the population per year across all countries in West Africa. Assuming a vaccine 70% effective only against disease, this vaccination scenario averted 276.6K (38.0K-755.9K) DALYs. However, in contrast to LASV vaccination, vaccine impact was more than three-fold greater when effective against infection as well as disease. For a vaccine 70% effective against both, this most ambitious vaccination scenario averted 1.2M (201.3K-2.7M) infections and 916.4K (108.0K-2.6M) DALYs, representing approximately 74% of the DALY burden imposed by Lassa-X. Vaccinating at half the rate (20% of the population/year) averted approximately 55% of the DALYs imposed by Lassa-X, while vaccinating at a low rate (2.5% of the population/year) averted just 11% of DALYs (**Supplementary tables G.5 to G.8**). Benefits of delivering vaccines at a higher rate outweighed benefits of initiating vaccination earlier (100 days vs. 160 days from outbreak detection), which in turn outweighed benefits of a vaccine with greater efficacy against infection and disease (90% vs. 70%).

**Table 3.** Projected impacts of 100 Days Mission vaccination campaigns in response to Lassa-X. The health-economic burden of Lassa-X averted due to vaccination, comparing a vaccine 70% effective only against disease (top) with a vaccine 70% effective against both infection and disease (bottom). Columns represent the vaccination scenarios considered and rows represent the outcomes averted. All figures represent means (95% uncertainty intervals) across all simulations, for the baseline scenario assuming a ten-fold greater risk of hospitalisation relative to Lassa virus infection. Societal costs combine outpatient treatment costs, hospital treatment costs and productivity losses. Costs are reported in International dollars (2021) and future monetary costs are discounted at 3%/year. DALY = disability-adjusted life year, VSL = value of statistical life, K = thousand, M = million, B = billion.

| Outcome averted due to vaccination | Vaccination scenario |  |  |  |  |  |
| --- | --- | --- | --- | --- | --- | --- |
|  | 2.5% of population vaccinated/year |  | 20% of population vaccinated/year |  | 40% of population vaccinated/year |  |
|  | 160d delay to first dose | 100d delay to first dose | 160d delay to first dose | 100d delay to first dose | 160d delay to first dose | 100d delay to first dose |
| **Vaccine 70% effective only against disease** |  |  |  |  |  |  |
| *Lassa-X infections (N)* | 0.0 (0.0-0.0) | 0.0 (0.0-0.0) | 0.0 (0.0-0.0) | 0.0 (0.0-0.0) | 0.0 (0.0-0.0) | 0.0 (0.0-0.0) |
| *Hospitalisations (N)* | 1.7K (294.0-3.9K) | 2.1K (361.5-4.7K) | 13.6K (2.4K-31.2K) | 16.6K (2.9K-37.3K) | 27.1K (4.7K-62.3K) | 33.3K (5.8K-74.8K) |
| *Deaths (N)* | 281.2 (35.6-811.0) | 344.5 (43.9-984.0) | 2.2K (284.6-6.5K) | 2.8K (350.9-7.9K) | 4.5K (569.2-13.0K) | 5.5K (704.9-15.7K) |
| *DALYs (N)* | 14.1K (1.9K-39.2K) | 17.3K (2.4K-47.2K) | 112.9K (15.4K-313.3K) | 138.1K (18.9K-377.6K) | 225.9K (30.9K-626.2K) | 276.6K (38.0K-755.9K) |
| *Impoverishing expenditures (N)* | 1.2K (206.2-2.7K) | 1.4K (255.7-3.2K) | 9.3K (1.6K-21.3K) | 11.4K (2.0K-25.3K) | 18.7K (3.3K-42.7K) | 22.9K (4.1K-50.5K) |
| *Societal costs (\$ 2021)* | 11.9M (1.8M-31.9M) | 14.6M (2.2M-38.8M) | 95.0M (14.3M-255.1M) | 116.8M (17.7M-310.5M) | 190.0M (28.6M-510.0M) | 233.7M (35.5M-620.8M) |
| *Monetised DALYs (\$ 2021)* | 2.2M (325.8K-6.6M) | 2.7M (398.8K-7.9M) | 17.9M (2.6M-52.8M) | 21.8M (3.2M-63.2M) | 35.8M (5.3M-105.8M) | 43.6M (6.4M-126.5M) |
| *VSL (\$ 2021)* | 109.8M (11.3M-346.3M) | 135.7M (13.8M-420.9M) | 878.2M (90.6M-2.8B) | 1.1B (110.3M-3.4B) | 1.8B (181.6M-5.5B) | 2.2B (221.7M-6.7B) |
| **Vaccine 70% effective against infection and disease** |  |  |  |  |  |  |
| *Lassa-X infections (N)* | 141.4K (27.5K-323.4K) | 183.8K (37.3K-399.2K) | 737.6K (146.5K-1.7M) | 916.0K (189.7K-2.0M) | 1.0M (200.5K-2.3M) | 1.2M (201.3K-2.7M) |
| *Hospitalisations (N)* | 12.8K (2.4K-31.3K) | 16.6K (3.1K-38.8K) | 66.1K (11.6K-152.2K) | 81.8K (14.4K-184.8K) | 93.0K (15.4K-214.5K) | 110.5K (16.8K-252.3K) |
| *Deaths (N)* | 2.1K (288.7-6.3K) | 2.7K (380.1-7.9K) | 11.0K (1.4K-31.4K) | 13.6K (1.6K-38.6K) | 15.4K (1.8K-44.6K) | 18.3K (2.0K-53.1K) |
| *DALYs (N)* | 106.7K (15.6K-304.9K) | 138.0K (20.6K-383.0K) | 550.4K (74.1K-1.5M) | 679.1K (90.0K-1.9M) | 773.2K (97.8K-2.1M) | 916.4K (108.0K-2.6M) |
| *Impoverishing expenditures (N)* | 8.8K (1.7K-21.9K) | 11.4K (2.2K-26.9K) | 45.5K (8.1K-105.0K) | 56.2K (10.0K-126.9K) | 63.8K (10.5K-147.3K) | 75.9K (11.6K-173.4K) |
| *Societal costs (\$ 2021)* | 88.8M (14.4M-244.5M) | 115.8M (19.1M-310.0M) | 464.4M (69.3M-1.3B) | 576.9M (80.4M-1.6B) | 656.0M (85.6M-1.8B) | 782.9M (91.4M-2.2B) |
| *Monetised DALYs (\$ 2021)* | 17.0M (2.6M-51.6M) | 21.9M (3.5M-64.4M) | 88.1M (12.2M-267.6M) | 107.9M (13.9M-321.8M) | 123.2M (14.9M-374.0M) | 144.5M (16.1M-429.3M) |
| VSL<br>(\$ 2021) | 809.8M<br>(95.2M-2.6B) | 1.1B<br>(126.4M-3.3B) | 4.3B<br>(423.4M-13.8B) | 5.4B<br>(482.0M-16.9B) | 6.1B<br>(518.6M-19.7B) | 7.3B<br>(557.3M-23.5B) |

## Discussion

This is to our knowledge the first burden of disease study for Lassa fever and the first to project impacts of Lassa vaccination campaigns on population health and economies.^15^ We estimated that 2.1M to 3.4M human LASV infections occur annually throughout West Africa, resulting in 15K to 35K hospitalisations and 1.3K to 8.3K deaths. These figures are consistent with recent modelling work estimating 900K to 4.4M human LASV infections per year,^14^ and an annual 5K deaths reported elsewhere.^3,16^ We further estimated that Lassa fever causes 2.0M DALYs, $1.6B in societal costs and $15.3B in lost VSL over ten years. Our modelling suggests that administering Lassa vaccines preventively to districts of Nigeria, Guinea, Liberia and Sierra Leone currently classified as “endemic” by WHO would avert a substantial share of the burden of disease in those areas. In our most expansive rollout scenario – in which vaccine reaches approximately 80% of individuals in endemic districts and 5% of individuals elsewhere over a 3-year period – a vaccine 70% effective against disease is projected to avert 164K DALYs, $128M in societal costs and $1.3B in VSL lost over ten years. This corresponds to a 10.5% reduction in Lassa fever DALYs in Nigeria given vaccination among 16.1% of the population, and a 24.4% reduction in DALYs across Guinea, Liberia and Sierra Leone given vaccination among 33.3% of the population. However, for the same rollout scenario, a vaccine 90% effective against both infection and disease could avert 240K DALYs, $188M in societal costs and $1.9B in VSL lost, corresponding to a 15.3% reduction in Lassa fever DALYs in Nigeria and a 35.3% reduction across Guinea, Liberia and Sierra Leone.

Impacts of the Lassa vaccination campaigns included in our analysis were modest in countries other than Nigeria, Guinea, Liberia and Sierra Leone. This is due primarily to these simulated campaigns reflecting a constrained global vaccine stockpile (<20M doses annually) and hence limited allocation to districts not currently classified as endemic by WHO. While our most optimistic vaccination scenario was projected to prevent as many as 1.9M (62%) infections in endemic-classified districts (**Supplementary figure E.4**), these areas cover just shy of 10% of the approximately 400M individuals living in West Africa. Yet our model predicts high Lassa fever incidence and disease burden in several “non-endemic” areas. This is consistent with seroprevalence data highlighting extensive underreporting of LASV infection across the region, particularly in Ghana, Côte d’Ivoire, Burkina Faso, Mali, Togo and Benin.^14,17–19^ Underreporting of Lassa fever is likely due to a combination of limited surveillance resources in affected countries, the mild and non-specific symptom presentation of most cases, seasonal fluctuations in infection incidence coincident with other febrile illnesses (malaria in particular), and stigma associated with infection, making robust estimation of Lassa fever burden a great challenge.^20^ Conversely, low case numbers in some areas estimated to be suitable for transmission^21^ may reflect truly limited burden, driven in part by significant spatiotemporal heterogeneity in LASV infection prevalence and the low dispersal rate of *M. natalensis*.^22^

It is important to put Lassa fever’s projected health-economic burden and impacts of vaccination in context, in particular given limited economic resources available for investment in infectious disease prevention in West Africa, and hence opportunity costs to investing in Lassa vaccination in lieu of other interventions. In Nigeria, 2021, we estimated an annual 48 (95% UI: 19-93) Lassa fever DALYs/100,000 population. This compares to previous estimates for various emerging, neglected and vaccine-preventable diseases, including trachoma (22 DALYs/100,000 population in Nigeria, 2019), yellow fever (25), rabies (34), lymphatic filariasis (54), intestinal nematode infections (63), diphtheria (80) and typhoid fever (93).^23^ We further predicted mean TVCs up to $2.20 per dose for preventive campaigns when considering societal costs and monetised DALYs. A global costing analysis across 18 common vaccines estimated a per-dose cost of $2.63 in low-income countries from 2011 to 2020, including supply chain and service delivery costs,^24^ suggesting that it may be feasible to achieve a maximum price per dose in line with our TVC estimates. However, real-world costs for any potential forthcoming vaccines are not yet known, and it is important to consider that vaccines currently undergoing clinical trials have distinct dosage regimens,^11^ and that our TVC estimates are specific to our model assumptions: a single-dose primary series with a booster dose after five years and 10% dose wastage. All else being equal, undiscounted TVC estimates for preventive campaigns would be roughly doubled or reduced by one-third, respectively, for a vaccine not requiring a booster dose or one requiring a 2-dose primary series.

The real-world cost-effectiveness of any forthcoming Lassa vaccine will depend not only on its dosage, price and clinical efficacy – estimates of which are not yet available – but also on the alternative interventions available. Novel small-molecule antivirals and monoclonal antibodies are in various stages of development,^25,26^ and may represent promising alternatives for prevention of severe Lassa fever. Our results further highlight how the choice of perspective can lead to divergent conclusions regarding vaccine cost-effectiveness.^27^ For instance, TVCs were roughly one order of magnitude greater when considering VSL instead of societal costs and monetised DALYs, up to $21.15 from $2.20 per dose. This disparity is consistent with a comparative analysis of health risk valuation, highlighting greatest TVC estimation when using VSL.^28^ Although our estimates of vaccine-averted DALYs, societal costs and lost VSL may complement one another to inform priority setting and decision making,^29^ caution is needed when comparing and potentially combining distinct economic metrics (and hence perspectives). In particular, the value inherent to VSL may encapsulate both economic productivity and health-related quality of life, so VSL must be considered independently of productivity losses and monetised life-years. Ultimately, defining the full value of vaccination in endemic areas will require ongoing engagement and priority setting across stakeholders,^30^ and may benefit from considering broader macroeconomic impacts of vaccination not included in our analysis.^31^ Yet even if a particular vaccine is identified as a priority by local stakeholders and is predicted to be cost-effective using context-specific willingness-to-pay thresholds and an appropriate perspective, investment will only be possible if vaccination is affordable, i.e. if sufficient economic resources are available to cover vaccine programme costs.

One major potential benefit to present investment in Lassa vaccination is increased readiness to rapidly develop and deploy vaccines against future LASV variants with pandemic potential. The COVID-19 pandemic demonstrated that prior research on coronaviruses and genetic vaccine technologies gave researchers an important head start on SARS-CoV-2 vaccine development in early 2020.^32^ In this context, we projected impacts of ambitious vaccination campaigns in response to the emergence of a hypothetical novel LASV variant with pandemic potential. While it is impossible to predict whether “Lassa-X” will evolve and exactly which characteristics it would have, this modelling represents a plausible scenario for its emergence and spread, totalling on average 1.7M infections, 150K hospitalisations and 25K deaths over roughly two years, resulting in 1.2M DALYs, $1.1B in societal costs and $10.1B in VSL lost. We estimated that a vaccine 70% effective against infection and disease, with delivery starting 100 days from the first detected case, could avert roughly one-tenth of Lassa-X’s health-economic burden assuming delivery of about 10M doses per year, or up to three-quarters of its burden given 160M doses per year. Such ambitious vaccination scenarios are in keeping with the stated goals of the 100 Days Mission,^13^ representing an expansive global effort to rapidly respond to emerging pandemic threats. In contrast to LASV, vaccination against Lassa-X was more than three-fold more impactful when blocking infection in addition to disease, due to indirect vaccine protection successfully slowing its explosive outbreak dynamics.

This work has several limitations. First, our projections of Lassa fever burden build upon recent estimates of spillover risk and viral transmissibility, but do not account for the potential evolution of these parameters over time, for instance due to projected impacts of climate change.^22^ Second, our model appears to overestimate the magnitude of seasonal fluctuations in incidence, potentially biasing not the total number of infections but how they are distributed through time. While peaks in Lassa fever risk during the dry season are well observed, including five-fold greater risk estimated in Nigeria,^33^ a large outbreak in Liberia during the rainy season in 2019/20 highlights that LASV nonetheless circulates year-round.^34^ Third, in assuming no LASV seroreversion among previously infected people, our model potentially underestimates the number of infections occurring annually. However, fitting the infection-hospitalisation ratio to hospital case data from Nigeria limits the sensitivity of model outcomes to this assumption. Fourth, our evaluation of the economic consequences of Lassa-X is conservative, as we do not account for the exportation of cases outside of West Africa, nor potential externalities of such a large epidemic, including negative impacts on tourism and trade, and the oversaturation and potential collapse of healthcare services. Fifth, since poor Lassa fever knowledge has been reported among both healthcare workers and the general population in several endemic areas,^35,36^ increased awareness resulting from vaccination campaigns could have positive externalities not considered in our analysis, including the adoption of infection prevention behaviours and timelier care-seeking. Conversely, poor Lassa fever knowledge could limit vaccine uptake, posing challenges to reaching the vaccine coverage targets considered here.

Finally, for both LASV and Lassa-X we do not stratify risks of infection, hospitalisation or death by sex or age, and infections in each country are assumed to be representative of the general population in terms of age, sex, employment and income. Seroepidemiological data from Sierra Leone show no clear association between antibodies to LASV and age, sex or occupation,^37^ and studies from hospitalised patients in Sierra Leone and Nigeria show conflicting relationships between age and mortality.^3,38,39^ Prospective epidemiological cohort studies such as the ongoing *Enable* programme will help to better characterise Lassa fever epidemiology – including the spectrum of illness, extent of seroreversion, and risk factors for infection and disease – in turn informing future modelling, vaccine trial design and intervention investment.^40^ In particular, better quantification of risk in groups believed to be at high risk of infection (e.g. healthcare workers) and severe disease (e.g. pregnant women) will help to inform targeted vaccination strategies, which are likely to be more cost-effective than the population-wide campaigns considered in our analysis. Nevertheless, a recent stakeholder survey highlights that the preferred vaccination strategy among Lassa fever experts in West Africa is consistent with the vaccine scenarios considered here, i.e. mass, proactive campaigns immunizing a wide range of people in high-risk areas, with corresponding demand forecasts reaching up to 100 million doses.^41^

### Conclusion

Our analysis suggests that vaccination campaigns targeting known Lassa fever hotspots will help to alleviate the large health-economic burden caused by this disease. However, expanding vaccination beyond WHO-classified “endemic” districts will be necessary to prevent the large burden of disease estimated to occur in neighbouring areas not currently classified as endemic. Improved surveillance is greatly needed to better characterise the epidemiology of Lassa fever across West Africa, helping to inform the design of vaccination campaigns that maximise population health by better targeting those at greatest risk of infection and severe outcomes. In the hypothetical event of a novel, highly pathogenic pandemic variant emerging and devastating the region, our modelling also suggests that the ambitious vaccination targets of the 100 Days Mission could have critical impact, helping to prevent up to three-quarters of associated health-economic burden. The probability of such a variant evolving is exceedingly difficult to predict, but investment in Lassa vaccination now could nonetheless have great additional health-economic value if facilitating a more rapid vaccine response in the event of a pandemic Lassa-related virus emerging.

## Methods

### Zoonotic LASV transmission

The incidence of LASV spillover was estimated by extending a previously published geospatial risk model by Basinski et al. (details in **Supplementary appendix A**).^14^ Briefly, this model synthesises environmental features, *M. natalensis* occurrence data and LASV seroprevalence data from both rodents and humans to predict rates of zoonotic LASV infection across West Africa. Environmental features were obtained as classification rasters from the Moderate Resolution Imaging Spectroradiometer dataset, including eleven land cover features and seasonally-adjusted measures of temperature, rainfall and vegetation. Occurrence data include historical captures of *M. natalensis* confirmed with genetic methods or skull morphology across 167 locations in 13 countries from 1977 to 2017. Rodent seropositivity data cover 13 studies testing *M. natalensis* for LASV across six countries from 1972 to 2014, while human seropositivity data cover 94 community-based serosurveys across five countries from 1970 to 2015.

Consistent with Basinski et al.,^14^ we used a GLM to predict human seroprevalence from modelled estimates of spillover risk at the level of 0.05° x 0.05° spatial pixels. To estimate incidence rates, a Susceptible-Infected-Recovered model was used to model transitions between susceptible (seronegative), infected (seropositive) and recovered (seropositive) states. To account for change in human population size over time, this model was augmented with data on per-capita human birth and death rates for each country for each year from 1960-2019. Using a forward Euler model with 4-week time-steps, we estimated the number of new infections in each time-step that reproduced modelled seroprevalence estimates in 2015, and stepped this forward to estimate infections in 2019, dividing by the 2019 population size to give the 2019 incidence rate in each pixel.^42^ Uncertainty in human LASV seroprevalence from the GLM was propagated forward to generate uncertainty in spillover incidence. Final non-aggregated estimates of spillover incidence from our model (at the pixel level) are shown in **Figure 1**, while aggregated estimates at the district level are shown in **Supplementary figure B.1**. Estimates of spillover incidence in endemic districts are shown in **Supplementary figures B.2** and **B.3**.

### Human-to-human transmission

We developed a stochastic branching process model to simulate infections arising from human-to-human transmission following spillover infection (**Supplementary appendix C.1**). To account for uncertainty in estimated annual spillover incidence, 99 distinct transmission simulations were run, with each one using as inputs a set of LASV spillover estimates corresponding to a particular centile. Each set contains 183 values (one for each district) and the same values are used for each of the 10 years of simulation.

To account for seasonality observed in Lassa fever case reports, annual incidence estimates are distributed across each epidemiological year according to a Beta distribution, as considered previously in Lerch *et al*.^43^ An outbreak tree was generated for each spillover event using an estimate of LASV’s basic reproduction number from the literature (*R*_0_ = 0.063),^43^ estimated from case data from a Lassa fever ward in Kenema Government Hospital, Sierra Leone, from 2010 to 2012.^44^ Infections in each outbreak tree are distributed stochastically through time following estimates of LASV’s incubation and infectious periods,^43^ and final outbreak trees are combined to generate the daily incidence of human-source infection in each district in the absence of vaccination. See **Supplementary table C.1** for LASV infection and transmission parameters.

### Lassa vaccination campaigns

We included 6 vaccination scenarios in which limited doses of vaccine are allocated across specific sub-populations of West Africa (see **Extended Data Table 2** and **Supplementary appendix C.2** for more detail). Vaccine doses are allocated preferentially to populations perceived to be at greatest risk of Lassa fever, i.e. those living in districts classified as Lassa fever endemic by WHO.^45^ In some scenarios, a small number of additional doses are allocated to non-endemic districts. In “constrained” scenarios, the total number of vaccine doses is constrained to reflect limited capacity to produce, stockpile and deliver vaccine. For these scenarios, cholera is used as a proxy disease for assumptions relating to vaccine stockpile and target coverage based on recent campaigns in West Africa.

In our vaccination scenarios developed with these constraints in mind, we consider both reactive vaccination (targeting specific districts in response to local outbreaks) and preventive vaccination (mass vaccinating across entire countries or districts regardless of local transmission patterns). Vaccination is assumed to confer immunity for five years after a single-dose primary series, with a single-dose booster administered five years after the initial dose. Vaccination is applied in the model by “pruning” zoonotic infections and ensuing person-to-person transmission chains, i.e. by retrospectively removing infections directly and indirectly averted as a result of vaccination (see **Supplementary appendix C.3** for more detail). We do not consider potential side-effects of vaccination.

### Health-economic burden of Lassa fever

A decision-analytic model describing the clinical progression of Lassa fever was developed to project the health and economic burden of disease and impacts of vaccination (**Supplementary appendix D.1**). Inputs into this model from our spillover risk map and branching process transmission model include, for each year, district and vaccination scenario: the total number of LASV infections, the number of infections averted due to vaccination, and the number of infections occurring in vaccinated individuals. The latter is included to account for vaccine preventing progression from infection to disease (**Supplementary appendix D.2**). Probability distributions for model parameters were estimated using data from the literature and are described in detail in **Supplementary appendix D.3**. Briefly, probabilities of hospitalisation and death were estimated from reported hospital case data in Edo and Ondo, Nigeria, from 2018 to 2021; durations of illness prior to and during hospitalisation were estimated from a prospective cohort study in a hospital in Ondo from 2018 to 2020; and hospital treatment costs were estimated from patients attending a specialist teaching hospital in Edo from 2015 to 2016 (see **Supplementary table D.1**).^5,8,38^

### Model outcomes

Lassa fever health outcomes estimated by our model include mild/moderate symptomatic cases, hospitalised cases, deaths, cases of chronic sequelae (sensorineural hearing loss) following hospital discharge, and disability-adjusted life years (DALYs). Economic outcomes include direct healthcare costs paid out-of-pocket or reimbursed by government, instances of catastrophic or impoverishing out-of-pocket healthcare expenditures, productivity losses, monetised DALYs, and the value of statistical life (VSL) lost (a population-aggregate measure of individuals’ willingness to pay for a reduction in the probability of dying).^46^ We report societal costs as the sum of healthcare costs and productivity losses. All monetary costs are reported in International dollars ($) 2021, and future monetary costs are discounted at 3%/year. Impacts of vaccination are quantified from outputs of the health-economic model as the difference in projected outcomes across parameter-matched runs of the model with and without vaccination.

To calculate threshold vaccine costs (TVC), we first summed relevant monetary costs for each simulation according to the economic perspective considered: healthcare costs and monetised DALYs, societal costs and monetised DALYs, or healthcare costs and VSL. The TVC is then calculated as the monetary costs averted due to vaccination divided by the number of vaccine doses allocated, including booster doses and wasted doses, and discounting future vaccine doses at 3%/year.

### Lassa-X

In addition to our analysis of Lassa fever, we consider the emergence of “Lassa-X”, a hypothetical future variant of LASV with pandemic potential due to both elevated clinical severity and increased propensity for human-to-human transmission. We assume that the clinical characteristics of Lassa-X are identical to Lassa fever (including sequelae risk and hospital case-fatality ratio), except that Lassa-X is accompanied by a ten-fold increase in risk of hospitalisation relative to Lassa fever. Then, to conceive plausible scenarios of Lassa-X transmission informed by empirical data, we assume that the inherent transmissibility of Lassa-X resembles that of Ebola virus during the 2013/16 West Africa outbreak. Ebola virus transmission is chosen as a surrogate for Lassa-X transmission because, like LASV, Ebola virus is a single-stranded RNA virus known to cause outbreaks in West Africa, results in frequent zoonotic spillover to humans from its animal reservoir, causes viral haemorrhagic fever, and spreads from human to human primarily through contact with infectious bodily fluids. Based on this conceptualisation of Lassa-X, we use a five-step approach to model its emergence and subsequent geospatial spread across West Africa, and to estimate the health-economic impacts of reactive “100 Days Mission” vaccination campaigns (described in detail in **Supplementary appendix F**).

### Simulation and statistical reporting

For each of 99 runs of the LASV transmission model and 100 runs of the Lassa-X transmission model, health-economic outcomes were calculated via 100 Monte Carlo simulations, in which input parameters for the health-economic model were drawn probabilistically from their distributions (**Supplementary table D.1**). In our base case we assume the vaccine is 70% effective only against disease. However, we also include scenarios with vaccine that is 90% effective against disease, 70% effective against both infection and disease, and 90% effective against both infection and disease. Final health and economic outcomes, as well as outcomes averted by vaccination, are reported as means and 95% uncertainty intervals (UIs) across all simulations over the ten-year time horizon of the model. In sensitivity analysis, we consider a 0% discounting rate, a lower risk of developing chronic sequelae subsequent to hospital discharge, and either the same or lower hospitalisation risk for Lassa-X relative to LASV. We also conduct univariate sensitivity analysis to identify the parameters driving outcome uncertainty. See **Supplementary appendix D.4** for more details. Estimates of Lassa fever burden are reported in accordance with the Guidelines for Accurate and Transparent Health Estimates Reporting (GATHER) statement. A GATHER checklist is provided in **Supplementary appendix H**.

### Data sharing

All underlying data and code for this article are available at www.github.com/drmsmith/lassaVac/.

### Credit taxonomy for authorship

David R M Smith: Formal Analysis, Investigation, Methodology, Project administration, Software, Visualization, Writing-original draft, Writing-review and editing.

Joanne Turner: Formal Analysis, Investigation, Methodology, Project administration, Software, Visualization, Writing-original draft, Writing-review and editing.

Patrick Fahr: Formal Analysis, Investigation, Methodology, Writing-original draft, Writing-review and editing

Lauren A Attfield: Formal Analysis, Investigation, Methodology

Paul R Bessell: Formal Analysis, Investigation, Methodology, Writing-review and editing

Christl A Donnelly: Methodology, Supervision, Writing-original draft, Writing-review and editing Rory Gibb: Supervision, Writing-review and editing

Kate E Jones: Supervision, Writing-review and editing

David W Redding: Methodology, Supervision, Writing-review and editing Danny Asogun: Writing-review and editing

Oladele Oluwafemi Ayodeji: Writing-review and editing Benedict N Azuogu: Writing-review and editing

William A Fischer II: Writing-review and editing Kamji Jan: Writing-review and editing

Adebola T Olayinka: Writing-review and editing David A Wohl: Writing-review and editing

Andrew A Torkelson: Methodology, Project Administration, Writing-original draft, Writing-review and editing

Katelyn A Dinkel: Methodology

Emily J Nixon: Conceptualization, Formal Analysis, Funding acquisition, Investigation, Methodology, Project administration, Software, Supervision, Writing-original draft, Writing-review and editing.

Koen B Pouwels: Conceptualization, Formal Analysis, Funding acquisition, Investigation, Methodology, Project administration, Software, Supervision, Writing-original draft, Writing-review and editing.

T Deirdre Hollingsworth: Conceptualization, Formal Analysis, Funding acquisition, Investigation, Methodology, Project administration, Software, Supervision, Writing-original draft, Writing-review and editing.

## Data Availability

All underlying data and code for this article are available at www.github.com/drmsmith/lassaVac/

## Acknowledgements

This work was funded by Coalition for Epidemic Preparedness Innovations (CEPI) through the Vaccine Impact Assessment project funding. The authors would like to acknowledge the CEPI project team (Project lead: Arminder Deol and Project co-lead: Christinah Mukandavire) for their continuous support and helpful discussions and the project’s external advisory team for their invaluable feedback. TDH thanks the Li Ka Shing Foundation for institutional funding. KBP is supported by the Medical Research Foundation (MRF-160-0017-ELP-POUW-C0909). CAD is funded by the National Institute for Health Research (NIHR) Health Protection Research Unit in Emerging and Zoonotic Infections (200907), a partnership between the United Kingdom Health Security Agency (UKHSA), the University of Liverpool, the University of Oxford and the Liverpool School of Tropical Medicine. DWR is supported by an MRC UKRI/Rutherford Fellowship (MR/R02491X/1, MR/R02491X/2) and Sir Henry Dale Research Fellowship (funded by the Wellcome Trust and the Royal Society) (220179/Z/20/Z, 220179/A/20/Z). This research was supported by the QMEE CDT, funded by NERC grant number NE/P012345/1 (LAA); the MRC Centre for Global Infectious Disease Analysis (LAA and CAD); the HPRU in Emerging and Zoonotic Infections (CAD) and the Trinity Challenge (The Sentinel Forecasting Project) (KEJ, RG, DWR). DAW is funded by NIH (NIAID: R01AI135105). We would like to thank Anita Lerch for personal communications and providing code that inspired our LASV transmission model. We thank Mark Todd from Dreaming Spires for assisting in optimising the speed for our stochastic branching process model for Lassa. We acknowledge Dr. Ian Smith, Head of Research Software Engineering at University of Liverpool IT Services, for his help running simulations using HTCondor. We thank Aidan Desjardins and Anna Borlase for early discussions on Lassa dynamics, Natasha Salant for beta testing our code, and Claudio Nunes-Alves for providing helpful comments on an earlier draft. The views expressed are those of the authors and not necessarily those of the institutions with which they are affiliated.

## Inclusion & Ethics

This modelling study did not involve the collection or use of any primary individual-level patient data. This study included local researchers throughout the research process, including stakeholder meetings, expert feedback and manuscript revision from Lassa fever researchers in regions where Lassa fever is endemic.

## Extended Data

**Extended Data Figure 1.**
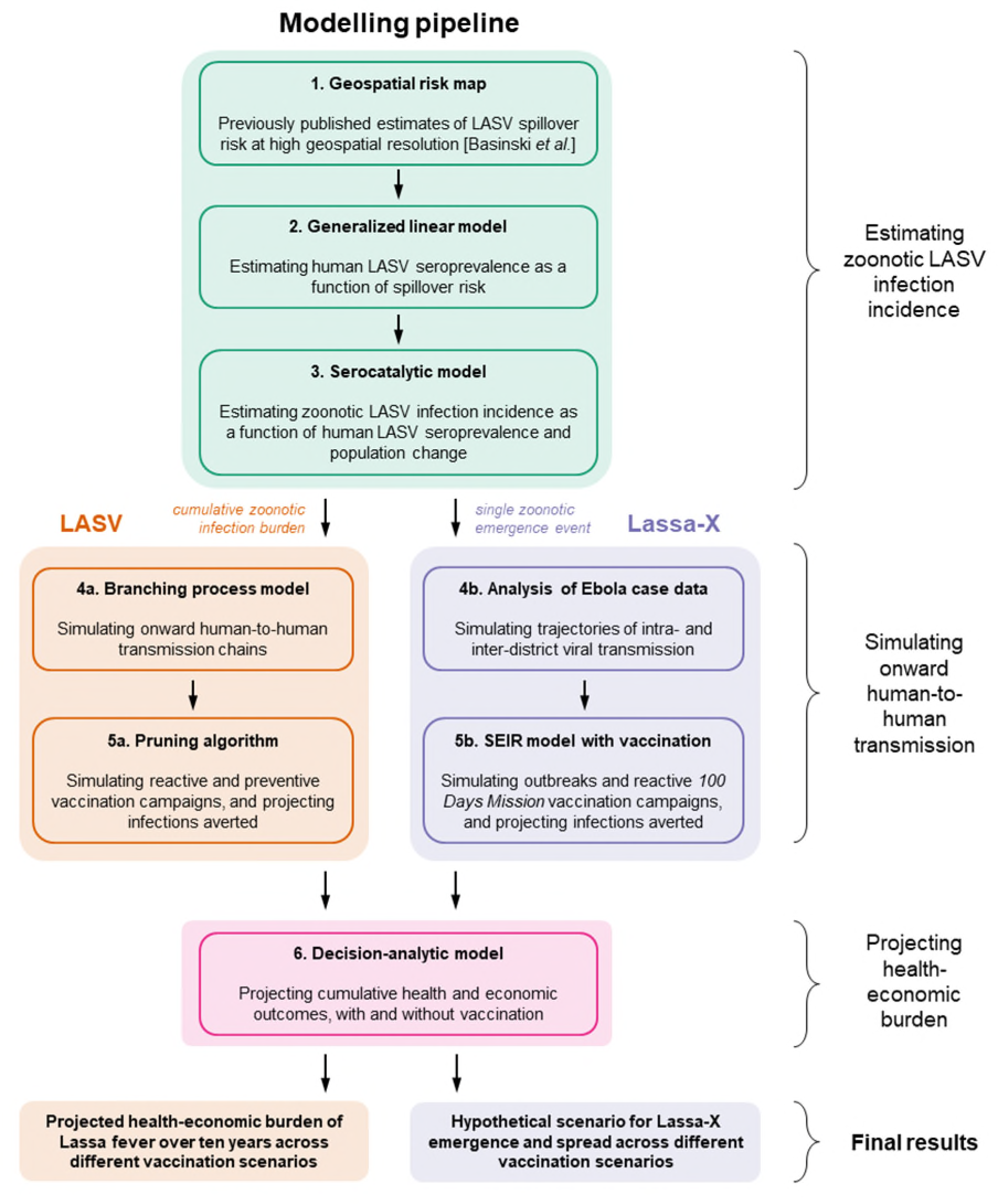
Model schematic. See Methods for details and **Supplementary figure D.1** for a schematic of the decision-analytic model describing disease progression. Pruning in step 5a refers to retrospectively removing infections averted due to vaccination from simulated transmission chains. LASV = *Lassa virus*.

**Extended Data Figure 2.**
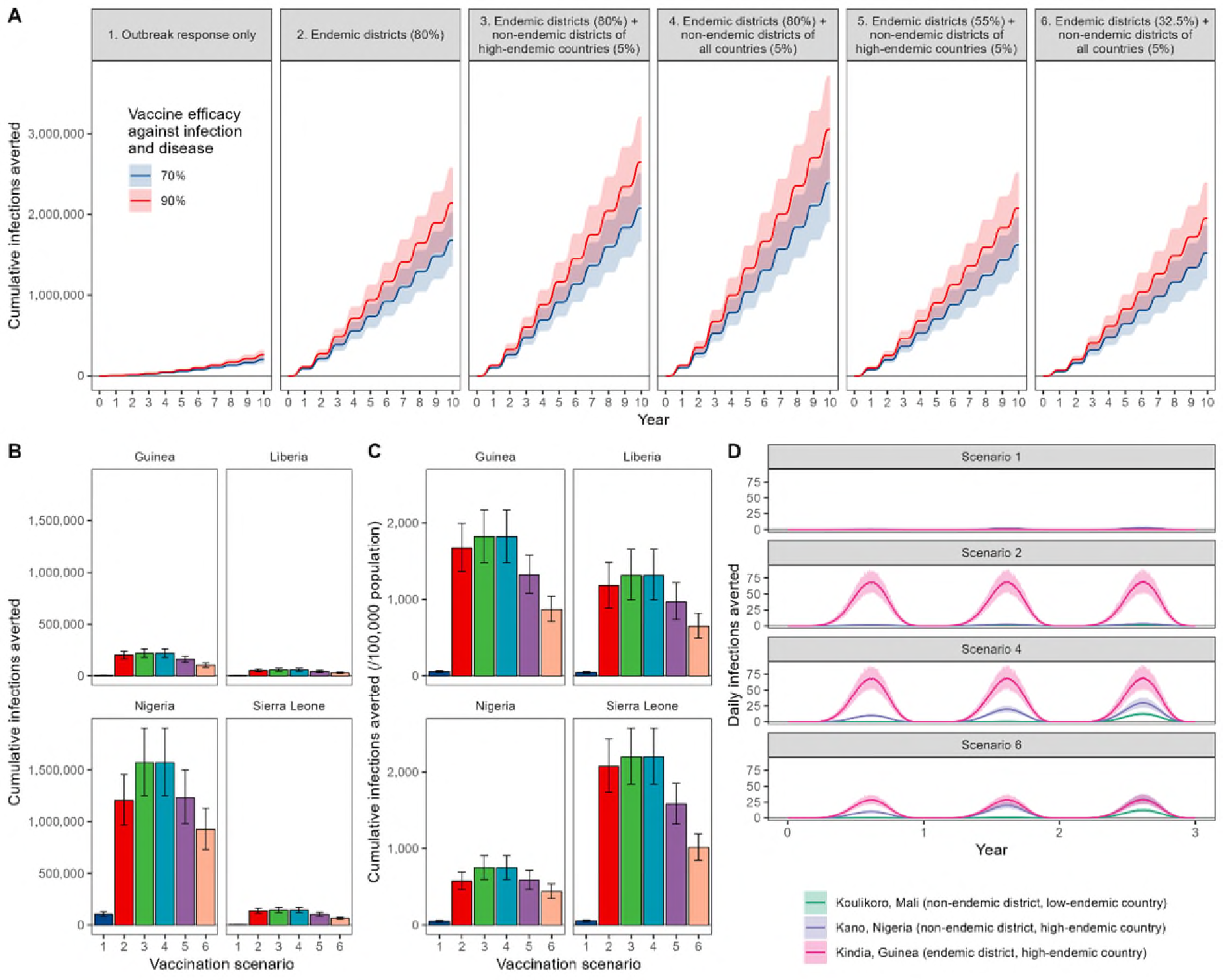
Impacts of a Lassa vaccine effective against infection and disease. (**A**) The mean cumulative number of LASV infections averted due to vaccination across the 15 countries included in the model, comparing vaccine efficacy against infection and disease of 70% (blue) versus 90% (red) across the six considered vaccination scenarios (panels). 95% uncertainty intervals are indicated by shading. (**B**) The mean cumulative number of infections averted over ten years under each vaccination scenario in the four countries classified as high-endemic (Guinea, Liberia, Nigeria and Sierra Leone). 95% uncertainty intervals are indicated by error bars. (**C**)The mean cumulative incidence of infections averted over ten years per 100,000 population under each vaccination scenario in the same four countries. 95% uncertainty intervals are indicated by error bars. (**D**) The mean daily number of infections averted by a vaccine with 70% efficacy against infection and disease over the first three years of vaccine rollout, in three distinct districts under four selected vaccination scenarios. 95% uncertainty intervals are indicated by shading.

**Extended Data Table 1.**
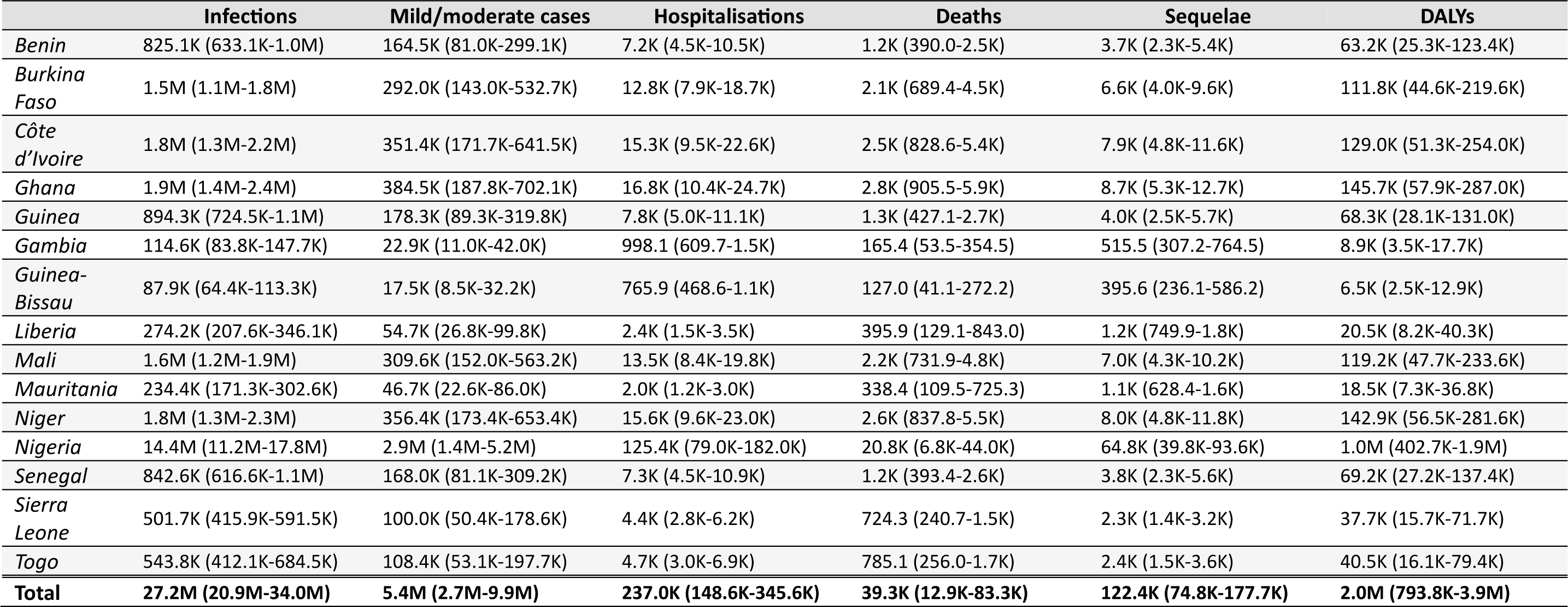
Cumulative health burden of Lassa fever by country over ten years in the absence of vaccination. All figures represent means (95% uncertainty intervals) across 100 runs of the health-economic model for each of 99 runs of the infection model, for the baseline scenario assuming a probability of sequelae of 62% among patients discharged from hospital. DALY = disability-adjusted life year, K = thousand, M = million.

|  | Infections | Mild/moderate cases | Hospitalisations | Deaths | Sequelae | DALYs |
| --- | --- | --- | --- | --- | --- | --- |
| *Benin* | 825.1K (633.1K-1.0M) | 164.5K (81.0K-299.1K) | 7.2K (4.5K-10.5K) | 1.2K (390.0-2.5K) | 3.7K (2.3K-5.4K) | 63.2K (25.3K-123.4K) |
| *Burkina Faso* | 1.5M (1.1M-1.8M) | 292.0K (143.0K-532.7K) | 12.8K (7.9K-18.7K) | 2.1K (689.4-4.5K) | 6.6K (4.0K-9.6K) | 111.8K (44.6K-219.6K) |
| *Côte d'Ivoire* | 1.8M (1.3M-2.2M) | 351.4K (171.7K-641.5K) | 15.3K (9.5K-22.6K) | 2.5K (828.6-5.4K) | 7.9K (4.8K-11.6K) | 129.0K (51.3K-254.0K) |
| *Ghana* | 1.9M (1.4M-2.4M) | 384.5K (187.8K-702.1K) | 16.8K (10.4K-24.7K) | 2.8K (905.5-5.9K) | 8.7K (5.3K-12.7K) | 145.7K (57.9K-287.0K) |
| *Guinea* | 894.3K (724.5K-1.1M) | 178.3K (89.3K-319.8K) | 7.8K (5.0K-11.1K) | 1.3K (427.1-2.7K) | 4.0K (2.5K-5.7K) | 68.3K (28.1K-131.0K) |
| *Gambia* | 114.6K (83.8K-147.7K) | 22.9K (11.0K-42.0K) | 998.1 (609.7-1.5K) | 165.4 (53.5-354.5) | 515.5 (307.2-764.5) | 8.9K (3.5K-17.7K) |
| *Guinea-Bissau* | 87.9K (64.4K-113.3K) | 17.5K (8.5K-32.2K) | 765.9 (468.6-1.1K) | 127.0 (41.1-272.2) | 395.6 (236.1-586.2) | 6.5K (2.5K-12.9K) |
| *Liberia* | 274.2K (207.6K-346.1K) | 54.7K (26.8K-99.8K) | 2.4K (1.5K-3.5K) | 395.9 (129.1-843.0) | 1.2K (749.9-1.8K) | 20.5K (8.2K-40.3K) |
| *Mali* | 1.6M (1.2M-1.9M) | 309.6K (152.0K-563.2K) | 13.5K (8.4K-19.8K) | 2.2K (731.9-4.8K) | 7.0K (4.3K-10.2K) | 119.2K (47.7K-233.6K) |
| *Mauritania* | 234.4K (171.3K-302.6K) | 46.7K (22.6K-86.0K) | 2.0K (1.2K-3.0K) | 338.4 (109.5-725.3) | 1.1K (628.4-1.6K) | 18.5K (7.3K-36.8K) |
| *Niger* | 1.8M (1.3M-2.3M) | 356.4K (173.4K-653.4K) | 15.6K (9.6K-23.0K) | 2.6K (837.8-5.5K) | 8.0K (4.8K-11.8K) | 142.9K (56.5K-281.6K) |
| *Nigeria* | 14.4M (11.2M-17.8M) | 2.9M (1.4M-5.2M) | 125.4K (79.0K-182.0K) | 20.8K (6.8K-44.0K) | 64.8K (39.8K-93.6K) | 1.0M (402.7K-1.9M) |
| *Senegal* | 842.6K (616.6K-1.1M) | 168.0K (81.1K-309.2K) | 7.3K (4.5K-10.9K) | 1.2K (393.4-2.6K) | 3.8K (2.3K-5.6K) | 69.2K (27.2K-137.4K) |
| *Sierra Leone* | 501.7K (415.9K-591.5K) | 100.0K (50.4K-178.6K) | 4.4K (2.8K-6.2K) | 724.3 (240.7-1.5K) | 2.3K (1.4K-3.2K) | 37.7K (15.7K-71.7K) |
| *Togo* | 543.8K (412.1K-684.5K) | 108.4K (53.1K-197.7K) | 4.7K (3.0K-6.9K) | 785.1 (256.0-1.7K) | 2.4K (1.5K-3.6K) | 40.5K (16.1K-79.4K) |
| **Total** | **27.2M (20.9M-34.0M)** | **5.4M (2.7M-9.9M)** | **237.0K (148.6K-345.6K)** | **39.3K (12.9K-83.3K)** | **122.4K (74.8K-177.7K)** | **2.0M (793.8K-3.9M)** |

**Extended Data Table 2.** Lassa vaccination scenarios. Scenario 1 includes reactive vaccination only, which is triggered in response to local outbreaks, while remaining scenarios 2 through 6 include preventive vaccination campaigns in addition to reactive vaccination. Vaccination coverage refers to the percentage of the general population targeted for vaccination in specified districts. Preventive vaccination in scenarios 2 through 4 is unconstrained, i.e. the number of doses reflects desired vaccination coverage levels, while preventive vaccination is constrained in scenarios 5 and 6, i.e. population coverage is constrained by an upper limit of doses to reflect a limited global vaccine stockpile (see **Supplementary table C.2**). The small vaccine pool reserved for reactive vaccination (1 million doses annually, shared across all districts proportionately to population size) is available immediately from year 1, while vaccination for preventive campaigns is rolled out to different countries in different years, generally to high-, medium- and low-endemic countries in years 1, 2 and 3, respectively (see **Supplementary table C.3**). A map showing the classification of endemicity across the countries and districts of West Africa is given in **Figure 1**. For both reactive and preventive vaccination, booster doses are allocated five years after the initial dose, and 90% of available doses are assumed to be delivered (i.e. all coverage targets were reduced by 10% wastage).

|  | Vaccination scenario |  |  |  |  |  |
| --- | --- | --- | --- | --- | --- | --- |
|  | 1 | 2 | 3 | 4 | 5 | 6 |
| Reactive vaccination coverage |  |  |  |  |  |  |
| Any district where outbreak response triggered | 0.2% | 0.2% | 0.2% | 0.2% | 0.2% | 0.2% |
| Preventive vaccination coverage |  |  |  |  |  |  |
| Endemic districts | 0 | 80% | 80% | 80% | 55% | 32.5% |
| Non-endemic districts in high-endemic countries | 0 | 0 | 5% | 5% | 5% | 5% |
| Non-endemic districts in other countries | 0 | 0 | 0 | 5% | 0 | 5% |
| Million vaccine doses allocated over first three years |  |  |  |  |  |  |
| Total | 2.4 | 34.1 | 43.8 | 52.3 | 33.9 | 33.4 |
| Million vaccine doses allocated over all ten years (includes booster doses) |  |  |  |  |  |  |
| Total | 12.1 | 75.5 | 94.8 | 111.8 | 74.9 | 74.1 |

## References

1. Andersen KG, Shapiro BJ, Matranga CB, et al. Clinical Sequencing Uncovers Origins and Evolution of Lassa Virus. Cell 2015; 162(4): 738–50.

2. Kafetzopoulou LE, Pullan ST, Lemey P, et al. Metagenomic sequencing at the epicenter of the Nigeria 2018 Lassa fever outbreak. Science 2019; 363(6422): 74-7.

3. Garry RF. Lassa fever - the road ahead. Nat Rev Microbiol 2023; 21(2): 87–96.

4. World Health Organization. Lassa fever. 2023. https://www.who.int/health-topics/lassa-fever#tab=tab_1 (accessed 20/10/2023.

5. Simons D. Lassa fever cases suffer from severe underreporting based on reported fatalities. Int Health 2023; 15(5): 608–10.

6. Ficenec SC, Percak J, Arguello S, et al. Lassa Fever Induced Hearing Loss: The Neglected Disability of Hemorrhagic Fever. Int J Infect Dis 2020; 100: 82–7.

7. Adetunji AE, Ayenale M, Akhigbe I, et al. Acute kidney injury and mortality in pediatric Lassa fever versus question of access to dialysis. Int J Infect Dis 2021; 103: 124–31.

8. Asogun D, Tobin E, Momoh J, et al. Medical cost of Lassa fever treatment in Irrua Specialist Teaching Hospital, Nigeria. Int J Basic Appl Innov Res 2016; 5(3): 62–73.

9. Tschismarov R, Van Damme P, Germain C, et al. Immunogenicity, safety, and tolerability of a recombinant measles-vectored Lassa fever vaccine: a randomised, placebo-controlled, first-in-human trial. Lancet 2023; 401(10384): 1267–76.

10. Mateo M, Reynard S, Journeaux A, et al. A single-shot Lassa vaccine induces long-term immunity and protects cynomolgus monkeys against heterologous strains. Sci Transl Med 2021; 13(597).

11. Sulis G, Peebles A, Basta NE. Lassa fever vaccine candidates: A scoping review of vaccine clinical trials. Trop Med Int Health 2023; 28(6): 420–31.

12. World Health Organization. Prioritizing diseases for research and development in emergency contexts. 2023. https://www.who.int/activities/prioritizing-diseases-for-research-and-development-in-emergency-contexts (accessed 20/10/2023.

13. Gouglas D, Christodoulou M, Hatchett R. The 100 Days Mission—2022 Global Pandemic Preparedness Summit. Emerg Infect Dis 2023; 29(3): e221142.

14. Basinski AJ, Fichet-Calvet E, Sjodin AR, et al. Bridging the gap: Using reservoir ecology and human serosurveys to estimate Lassa virus spillover in West Africa. PLoS Comput Biol 2021; 17(3): e1008811.

15. Di Bari C, Venkateswaran N, Fastl C, et al. The global burden of neglected zoonotic diseases: Current state of evidence. One Health 2023; 17: 100595.

16. US CDC. Lassa fever. 2022. https://www.cdc.gov/vhf/lassa/index.html (accessed 30/09/2023.

17. Safronetz D, Sogoba N, Diawara SI, et al. Annual Incidence of Lassa Virus Infection in Southern Mali. Am J Trop Med Hyg 2017; 96(4): 944–6.

18. Emmerich P, Gunther S, Schmitz H. Strain-specific antibody response to Lassa virus in the local population of west Africa. J Clin Virol 2008; 42(1): 40–4.

19. Yadouleton A, Picard C, Rieger T, et al. Lassa fever in Benin: description of the 2014 and 2016 epidemics and genetic characterization of a new Lassa virus. Emerg Microbes Infect 2020; 9(1): 1761–70.

20. Bausch DG, Demby AH, Coulibaly M, et al. Lassa fever in Guinea: I. Epidemiology of human disease and clinical observations. Vector Borne Zoonotic Dis 2001; 1(4): 269–81.

21. Mylne AQ, Pigott DM, Longbottom J, et al. Mapping the zoonotic niche of Lassa fever in Africa. Trans R Soc Trop Med Hyg 2015; 109(8): 483–92.

22. Klitting R, Kafetzopoulou LE, Thiery W, et al. Predicting the evolution of the Lassa virus endemic area and population at risk over the next decades. Nat Commun 2022; 13(1): 5596.

23. Angell B, Sanuade O, Adetifa IMO, et al. Population health outcomes in Nigeria compared with other west African countries, 1998-2019: a systematic analysis for the Global Burden of Disease Study. Lancet 2022; 399(10330): 1117–29.

24. Portnoy A, Ozawa S, Grewal S, et al. Costs of vaccine programs across 94 low- and middle-income countries. Vaccine 2015; 33 Suppl 1: A99–108.

25. Amberg SM, Snyder B, Vliet-Gregg PA, et al. Safety and Pharmacokinetics of LHF-535, a Potential Treatment for Lassa Fever, in Healthy Adults. Antimicrob Agents Chemother 2022; 66(11): e0095122.

26. Cross RW, Heinrich ML, Fenton KA, et al. A human monoclonal antibody combination rescues nonhuman primates from advanced disease caused by the major lineages of Lassa virus. Proc Natl Acad Sci U S A 2023; 120(34): e2304876120.

27. Kim DD, Silver MC, Kunst N, Cohen JT, Ollendorf DA, Neumann PJ. Perspective and Costing in Cost-Effectiveness Analysis, 1974-2018. Pharmacoeconomics 2020; 38(10): 1135–45.

28. Park M, Jit M, Wu JT. Cost-benefit analysis of vaccination: a comparative analysis of eight approaches for valuing changes to mortality and morbidity risks. BMC Med 2018; 16(1): 139.

29. Laxminarayan R, Jamison DT, Krupnick AJ, Norheim OF. Valuing vaccines using value of statistical life measures. Vaccine 2014; 32(39): 5065–70.

30. Hutubessy R, Lauer JA, Giersing B, et al. The Full Value of Vaccine Assessments (FVVA): a framework for assessing and communicating the value of vaccines for investment and introduction decision-making. BMC Med 2023; 21(1): 229.

31. Jit M, Hutubessy R, Png ME, et al. The broader economic impact of vaccination: reviewing and appraising the strength of evidence. BMC Med 2015; 13: 209.

32. Mao W, Zimmerman A, Urli Hodges E, et al. Comparing research and development, launch, and scale up timelines of 18 vaccines: lessons learnt from COVID-19 and implications for other infectious diseases. BMJ Glob Health 2023; 8(9).

33. Akhmetzhanov AR, Asai Y, Nishiura H. Quantifying the seasonal drivers of transmission for Lassa fever in Nigeria. Philos Trans R Soc Lond B Biol Sci 2019; 374(1775): 20180268.

34. Jetoh RW, Malik S, Shobayo B, et al. Epidemiological characteristics of Lassa fever cases in Liberia: a retrospective analysis of surveillance data, 2019-2020. Int J Infect Dis 2022; 122: 767-74.

35. Wada YH, Ogunyinka IA, Yusuff KB, et al. Knowledge of Lassa fever, its prevention and control practices and their predictors among healthcare workers during an outbreak in Northern Nigeria: A multi-centre cross-sectional assessment. PLoS Negl Trop Dis 2022; 16(3): e0010259.

36. Aromolaran O, Samson TK, Falodun OI. Knowledge and practices associated with Lassa fever in rural Nigeria: Implications for prevention and control. J Public Health Afr 2023; 14(9): 2001.

37. Grant DS, Engel EJ, Roberts Yerkes N, et al. Seroprevalence of anti-Lassa Virus IgG antibodies in three districts of Sierra Leone: A cross-sectional, population-based study. PLoS Negl Trop Dis 2023; 17(2): e0010938.

38. Duvignaud A, Jaspard M, Etafo IC, et al. Lassa fever outcomes and prognostic factors in Nigeria (LASCOPE): a prospective cohort study. Lancet Glob Health 2021; 9(4): e469–e78.

39. Shaffer JG, Grant DS, Schieffelin JS, et al. Lassa fever in post-conflict Sierra Leone. PLoS Negl Trop Dis 2014; 8(3): e2748.

40. Penfold S, Adegnika AA, Asogun D, et al. A prospective, multi-site, cohort study to estimate incidence of infection and disease due to Lassa fever virus in West African countries (the Enable Lassa research programme)-Study protocol. PLoS One 2023; 18(3): e0283643.

41. Kabore L, Pecenka C, Hausdorff WP. Lassa fever vaccine use cases and demand: Perspectives from select West African experts. Vaccine 2024; 42(8): 1873–7.

42. Attfield LA. Mathematical modelling of the environmental and ecological drivers of zoonotic disease with an application to Lassa fever: Imperial College London; 2022.

43. Lerch A, Ten Bosch QA, L’Azou Jackson M, et al. Projecting vaccine demand and impact for emerging zoonotic pathogens. BMC Med 2022; 20(1): 202.

44. Lo Iacono G, Cunningham AA, Fichet-Calvet E, et al. Using modelling to disentangle the relative contributions of zoonotic and anthroponotic transmission: the case of lassa fever. PLoS Negl Trop Dis 2015; 9(1): e3398.

45. World Health Organization. Geographic distribution of Lassa fever in West African affected countries, 1969-2018. 2018. https://cdn.who.int/media/images/default-source/health-topics/lassa-fever/lassa-fever-countries-2018png.tmb-1024v.png?sfvrsn=10af107d_7 (accessed 16/10/2023.

46. Robinson LA, Hammitt JK, Cecchini M, et al. Reference Case Guidelines for Benefit-Cost Analysis in Global Health and Development, 2019.

