## Supplementary appendix for "Health and economic impacts of Lassa vaccination campaigns in West Africa"

**Contents:**

A. LASV spillover model

B. LASV spillover estimates

C. LASV transmission model
   1. Stochastic branching process model
   2. Vaccination scenarios
   3. Simulating vaccination by pruning infections

D. Health-economic model
   1. Model structure and outcomes
   2. Outcomes averted due to vaccination
   3. Parameter inputs
   4. Simulations and sensitivity analysis

E. Supplementary results for Lassa fever
   1. Health and economic burden in the absence of vaccination
   2. Vaccine impact

F. Lassa-X spillover, geospatial spread, transmission and vaccination
   1. Lassa-X spillover and geospatial spread
   2. Quantifying Ebola-like transmission dynamics
   3. Compartmental model and simulation of Lassa-X outcomes

G. Supplementary results for Lassa-X
   1. Lassa-X burden in the absence of vaccination
   2. Vaccine impact

H. GATHER checklist

I. Appendix references

#### Appendix A. Lassa spillover model

The incidence of LASV spillover was estimated by extending a previously published geospatial risk model by Basinski et al, which synthesises environmental features, *M. natalensis* occurrence data and Lassa virus seroprevalence estimates from both rodents and humans to estimate rates of zoonotic LASV infection across West Africa.[1] This model consists of several steps. First, environmental features are used to estimate a spatial surface of LASV spillover risk, which is generated by combining two spatial risk layers: (i) a classification score between 0 and 1, obtained using a boosted classification tree, indicating the likelihood that a spatial pixel (modelled in 0.05°x0.05° pixels, but later aggregated to the district level for analyses using spatial data from the Database of Global Administrative Areas)[2] contains the primary rodent reservoir (*M. natalensis)*, and (ii) a classification score between 0 and 1, again obtained using a boosted classification tree, indicating the likelihood that LASV circulates within the local *M. natalensis* population, conditional on the rodent being present in the spatial pixel of interest. The product of these two layers describes *spillover risk*, the probability that a pixel contains *M. natalensis* and LASV simultaneously.

Second, we used this spatial layer of spillover risk to predict human seroprevalence across the different districts using a generalised linear model (GLM). This GLM regresses seroprevalence estimates from human serosurveys onto the layer of spillover risk,

$$y = \alpha + \beta x$$

Where $y$ is the predicted seroprevalence, $\alpha$ the intercept and $\beta$ the fitted coefficient to observed seroprevalence ($x$).

Third, following Attfield (2022), a Susceptible-Infected-Recovered (SIR) model is used to model transition of humans between susceptible (seronegative), LASV-infected (seropositive), and recovered (seropositive) compartments.[3] Unlike the model by Basinski et al., which assumes a constant population size at steady-state equilibrium, this model accounts for increasing human population size in West Africa using World Bank 2019 birth and death rates at the country level, thus accounting for potential impacts of a growing population size on the force of infection.[4-6] It is important to note that this model, in line with the original model proposed by Basinksi et al. (2021), ignores any human-to-human transmission events. Finally, the SIR model was fitted to spatial human seroprevalence estimates from 2015 to predict the number of new zoonotic infections for each year from 1960 to 2015, using a forward Euler model with 4-week time-steps.[3] The predicted number of zoonotic infections was then fitted forward to 2019, accounting for demographic turnover. The model was fitted twice, allowing for reversion to susceptibility or without reversion, but herein we conservatively use only the model without reversion. From this an incidence rate of LASV spillover was estimated using the force of infection and birth and death rates. While the assumption of no reversion may lead to an underestimate of the total number of infections, since infection-hospitalisation risk in our model is calibrated to hospital case surveillance data (see **Appendix D.3**), this assumption does not impact our estimates of the major drivers of Lassa fever burden (numbers of hospitalised cases, deaths and individuals with sequelae).

To account for uncertainty in spillover risk, we generated three seroprevalence layers from the fitted GLM by Basinski et al (2021) by considering the mean and standard error (SE) of the model fit. The three spatial layers were the mean seroprevalence ± 1SE. From each of these layers we modelled spillover incidence, yielding mean spillover incidence (notated $\bar{L}$), mean – 1SE spillover incidence ($L_l$), and mean + 1SE spillover incidence ($L_u$). The full range of sampled values is t-distributed with 93 degrees of freedom, which describes the range of error from the GLM of Basinski et al. (2021). We then include uncertainty aggregated at the district level by taking each centile from the 1st to 99th centiles of the t-distribution ($b$), and generating each spillover incidence centile as $\bar{L} + SE \times b$. This resulted in a final distribution of 99 estimates of LASV spillover incidence for each of the 183 districts included in the model.

#### Appendix B. LASV spillover estimates

Non-aggregated spillover incidence estimates (at the pixel level) are shown in **Figure 1**, while aggregated estimates showing spillover incidence at the district level are shown in **Figure B.1**.

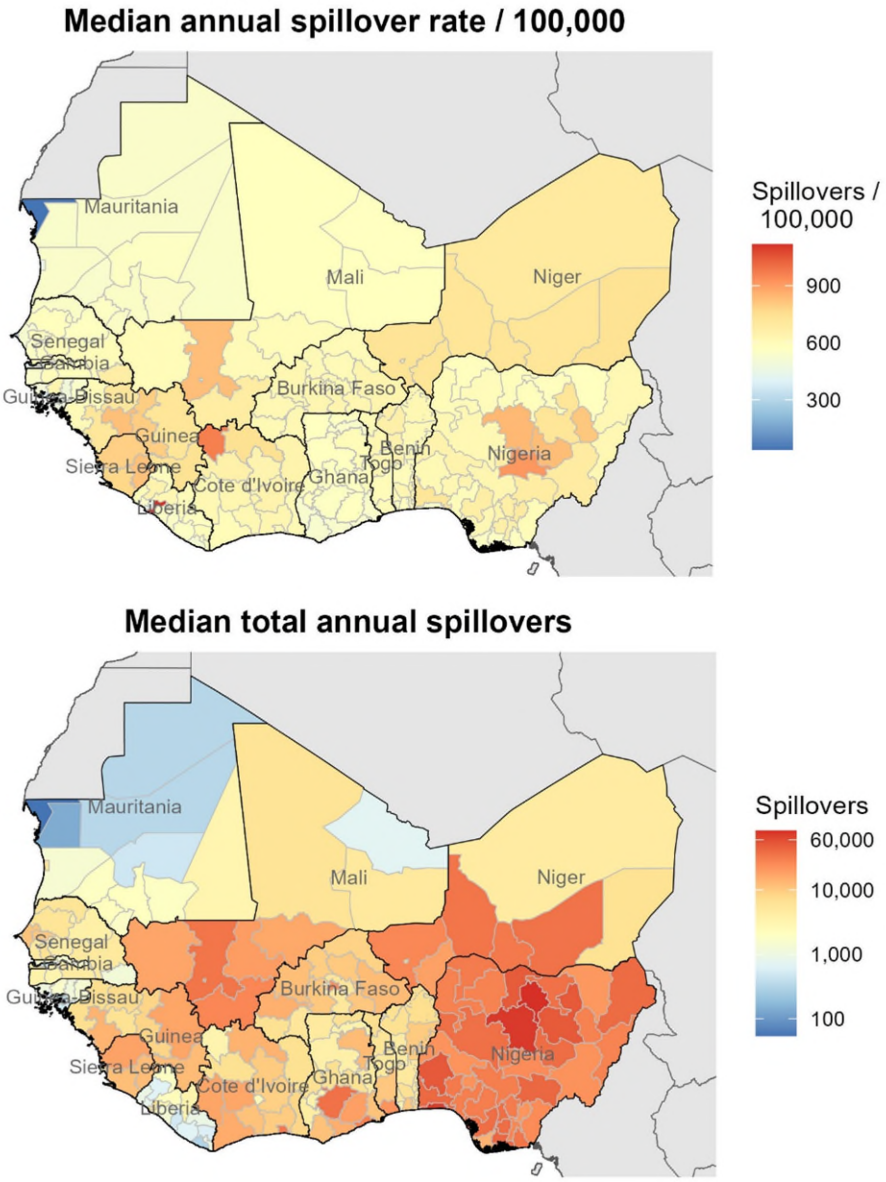


***Figure B.1. LASV spillover aggregated at the district level.*** *Top: the median annual incidence of zoonotic LASV infection as estimated by our risk map, aggregated at the level of sub-national "districts" (ADM1 regions). Bottom: the median total annual number of zoonotic LASV infections as estimated by our risk map, aggregated at the district level. LASV: Lassa virus.*

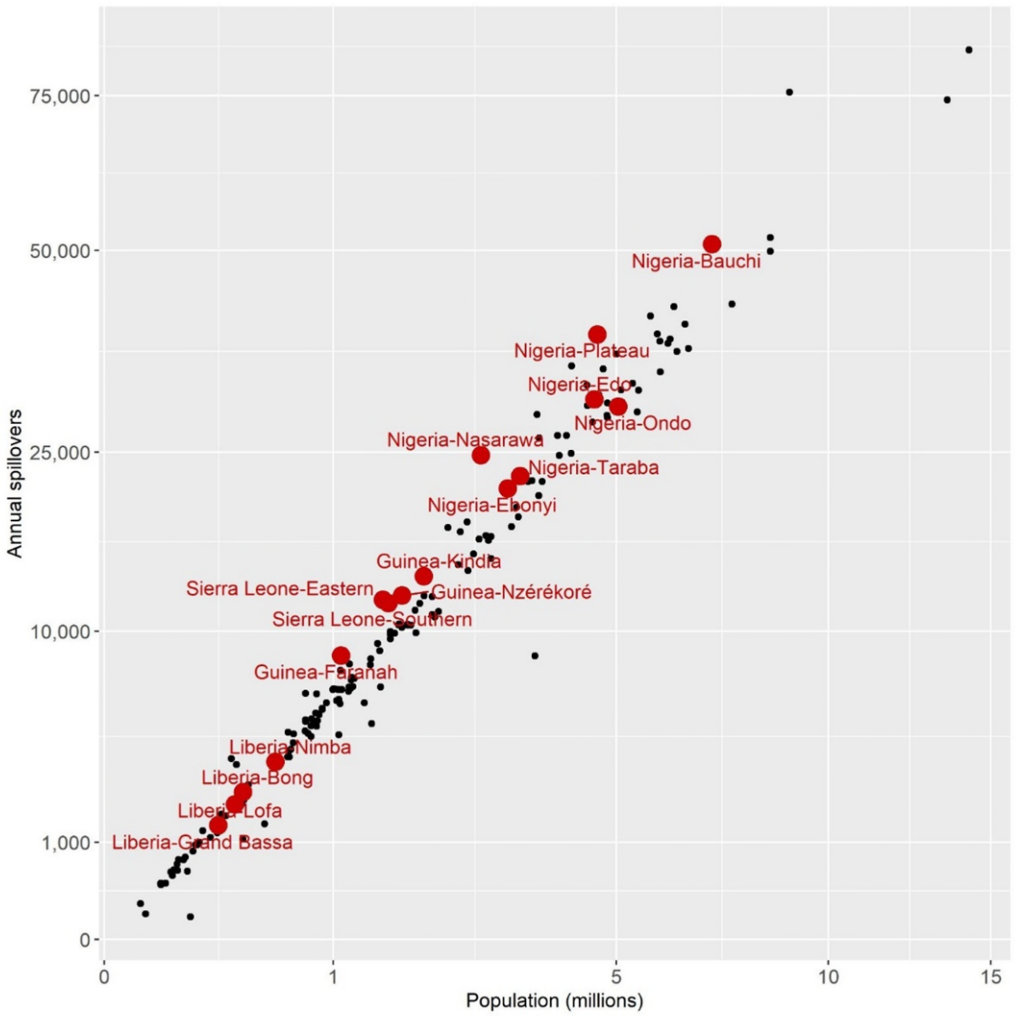


***Figure B.2. LASV spillover infections at the district level.*** *The annual number of LASV infections resulting from spillover in each district estimated by our risk map ($L_{d,y}$) against the population size of each district ($N_d$). Red dots represent districts classified as endemic. LASV: Lassa virus.*

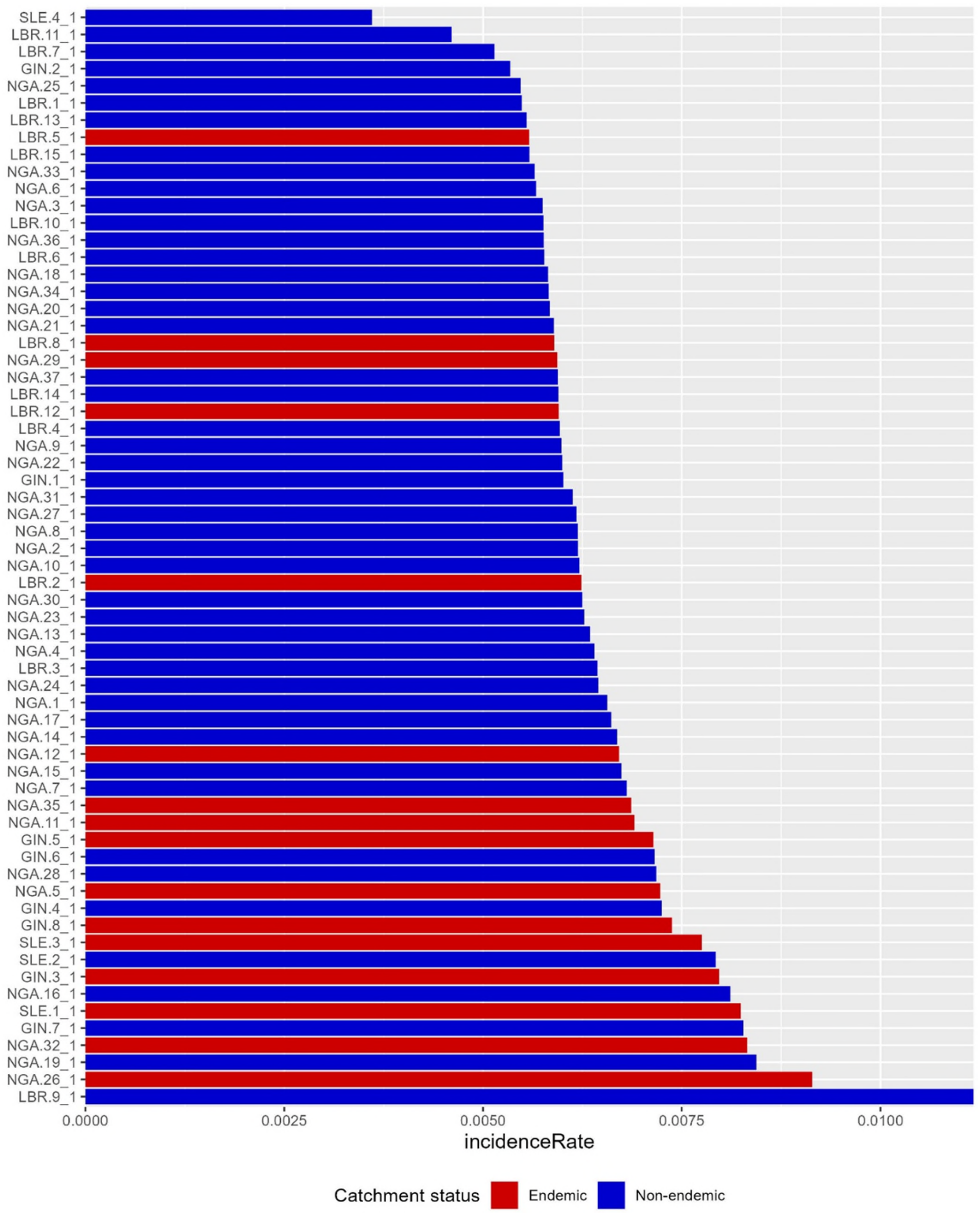


***Figure B.3. LASV spillover incidence at the district level in high-endemic countries.*** *The annual incidence rate $L_{d,y}/N_d$ of LASV spillover infection (x-axis) in each district (y-axis) as estimated by our risk map. Red bars represent districts classified by WHO as endemic, and blue bars represents districts classified as non-endemic. LASV: Lassa virus, WHO: World Health Organization.*

### Appendix C. LASV transmission model

#### *C.1. Stochastic branching process model*

A stochastic branching process model was used to simulate outbreak trees subsequent to LASV spillover from *Mastomys natalensis* to humans. The approach used to estimate the annual number of spillover infections in each district is described in **Appendix B**. To estimate the number of LASV infections resulting from human-to-human transmission, outbreak trees were simulated for each spillover infection using a stochastic branching process model based on four key parameters. This model is adapted from Lerch et al., who estimated distributions for model parameters by collating weekly LASV spillover case data from WHO outbreak reports, ProMED reports, country-level reports and a literature search.[7]

Let $L_{d,y}$ be the total number of spillover LASV infections occurring in humans in district $d$ and year $y$. The timing $\tau_{d,y,i}$ of each $i^{th}$ spillover infection $L_{d,y,i}$ in each epidemiological year is distributed seasonally, with each infection's timing drawn randomly from a Beta distribution,

$$\tau_{d,y,i} \sim \mathrm{B}(a = 9.53, b = 6.44) \times 365$$

To simulate the number of first-generation secondary infections resulting from each spillover infection, we use a Poisson process with mean $R_0$,

$$H_{d,y,i} \sim \mathrm{Pois}(R_0 = 0.063)$$

Accounting for subsequent generations of human-to-human transmission, the recurrence equation that represents the number of secondary infections $E_{d,y,i,n}$ in a given transmission generation $n$ is a classic Bienaymé-Galton-Watson branching process given by

$$E_{d,y,i,n} = \sum_{h=1}^{E_{d,y,i,n-1}} H_{d,y,i,n,h}$$

where $H_{d,y,i,n,h}$ represents the number of secondary infections generated at generation $n$ by the previous infection $h$. The sum total of human-to-human infections in each district and year of simulation across all generations of transmission is given by

$$E_{d,y} = \sum_{i} \sum_{n} E_{d,y,i,n}$$

For each infection, the incubation period $p_{d,y,i}$ and infectious period $q_{d,y,i}$ are randomly drawn from Gamma distributions

$$p_{d,y,i} \sim \Gamma(shape = 11.12, scale = 0.92)$$

$$q_{d,y,i} \sim \Gamma(shape = 1.86, scale = 6.07)$$

and the timing of transmission to each subsequent generation is distributed accordingly.

Finally, the total number of LASV infections $U$, including both spillover infection and human-to-human transmission, across all 183 districts and 10 years of simulation included in the model is

$$U = \sum_{d=1}^{183} \sum_{y=1}^{10} \left(L_{d,y} + E_{d,y}\right)$$

As described in the main text and further in **Appendix B**, we account for uncertainty in the geospatial LASV spillover risk map underlying our model,[1,3] resulting in a distribution of $\varepsilon = 99$ estimates of the number of LASV spillover infections $L_{d,\varepsilon}$ for each district. Within each district and simulation, the same spillover risk estimate is used for each of the ten years of simulation. We ran our branching process model for each simulation, resulting in a final mean estimate of LASV infection burden,

$$\bar{U} = \frac{\sum_{\varepsilon} U_{\varepsilon}}{\varepsilon}$$

Simulations were dispatched to computing resources at the University of Liverpool using HTCondor, a specialised workload management system for computer-intensive jobs (https://htcondor.org/, https://condor.liv.ac.uk). This branching process was implemented as an algorithm in R and is available at www.github.com/drmsmith/lassaVac/.

***Table C.1. LASV parameters.*** *Model parameters describing LASV infection and transmission in the stochastic branching process model. CI = confidence interval, LASV = Lassa virus.*

| Parameter | Mean [95% CI] | Distribution [parameters] | Notes |
|---|---|---|---|
| *Spillover timing ($\tau$)* | 0.60 [0.36, 0.815] | Beta [α=9.53, β=6.44] | The annual estimated number of spillovers was distributed seasonally by fitting a Beta distribution to weekly spillover case data.[7] |
| *Basic reproduction number ($R_0$)* | 0.063 | / | Point estimate from a branching process transmission model fit to nosocomial case data.[7] |
| *Infectious period ($q$)* | 11.31 days [1.22, 32.37] | Gamma [shape=1.86, scale=6.07] | Estimated mean and standard deviation derived by fitting a Gamma distribution to estimates from the literature.[7] |
| *Incubation period ($p$)* | 10.26 days [5.14, 17.11] | Gamma [shape=11.12, scale=0.92] | Estimated mean and standard deviation derived by fitting a Gamma distribution to estimates from the literature.[7,8] |

#### *C.2. Vaccination scenarios*

Six vaccination scenarios are considered, which vary in terms of assumptions regarding the geographic allocation of vaccine according to Lassa fever endemicity classification, the use of vaccine for reactive outbreak response versus preventive vaccination, population vaccine coverage targets and the corresponding number of vaccine doses required to meet those targets. These vaccination scenarios are summarised in **Table C.2**., where the number of doses administered accounts for assumed 10% wastage relative to vaccine dose targets.

*Endemicity classification*

Levels of Lassa fever endemicity for the 15 countries included in the analysis were classified by referencing an outbreak distribution map from the United States Centers for Disease Control and Prevention (CDC).[9] Countries labelled as high endemic (Guinea, Liberia, Nigeria, and Sierra Leone) are those identified as reporting large outbreaks and consistent infections. Medium endemic countries (Benin, Côte d'Ivoire, Ghana, and Togo) are those with fewer reported infections and/or occasional isolation or serological evidence of infection. Low endemic countries (Burkina Faso, Gambia, Guinea Bissau, Mali, Mauritania, Niger, and Senegal) include parts of or entire countries where no known evidence of Lassa infections is reported.[9] Subnational classification of Lassa fever endemicity in high endemic countries was referenced from a distribution map from WHO.[10] Districts within Guinea (Kindia, Faranah, Nzérékoré), Liberia (Bong, Grand Bassa, Lofa, Nimba), Nigeria (Bauchi, Ebonyi, Edo, Nasarawa, Ondo, Plateau, Taraba), and Sierra Leone (Bo, Kailahun) are labelled as endemic by the WHO. The endemic districts identified for Guinea, Liberia, and Nigeria are at the level of administrative units level 1, whereas the endemic districts for Sierra Leone are at the level of administrative units level 2.[10] As our model operates on level 1 administrative units, level 1 units of Sierra Leone were classified as endemic when any constituent level 2 units were classified as endemic. These endemicity classifications are visualised in **Figure 1** in the main text.

*Vaccine allocation and dosing*

Vaccination scenario 1 describes the use of reactive vaccination in response to local outbreaks. Vaccine dosing for this strategy was constrained to an annual stockpile of 1 million doses. This value was selected to split lower estimates from CEPI's stockpile plans for chikungunya (200,000 doses per year) and higher estimates from the early global cholera stockpiles (~2 million doses per year).[11,12]

Remaining scenarios 2 through 6 all include population-wide preventive mass vaccination campaigns in addition to reactive vaccination. These scenarios were designed to cover varying percentages of the population based on the district's endemicity level. Focal targets for these preventative campaigns were endemic districts within high endemic countries (Guinea, Liberia, Nigeria and Sierra Leone). In these districts, preventative campaigns aimed to cover 80% of the total population. This coverage target was based on previous campaigns against cholera in or near West Africa,[13,14] as well as WHO guidance.[15] Cholera was chosen as it shares similarities with Lassa fever in the estimated scale of their incidence rates in West Africa (estimates range from the hundreds of thousands to the low millions of cases per year)[16] and infection-fatality ratios (≤1%)[17] and because cholera vaccination campaigns have recently been conducted successfully in West Africa in response to outbreaks.[13,14] In non-endemic districts in high, medium, and low endemic countries, the preventative campaigns targeted 5% of the population, representing generally low anticipated rates of Lassa vaccine uptake in areas classified as non-endemic.

Scenarios 2, 3 and 4 include “unconstrained” preventive vaccination, where the number of doses reflects population coverage targets in different districts, regardless of total population size (**Table C.3**). Of these, the narrowest scenario (Scenario 2) aimed to only vaccinate populations in endemic districts of high endemic countries. The wider unconstrained campaigns expanded to include non-endemic districts of high endemic countries (Scenario 3) and non-endemic districts in the rest of West

Africa (Scenario 4). Scenarios 5 and 6 include “constrained” preventive vaccination, where the number of doses in endemic districts is limited to respect an assumed limited global vaccine stockpile. These supply-constrained scenarios were designed assuming a manufacturing constraint of roughly 20 million doses per year. For these scenarios, population coverage targets in non-endemic districts of high endemic countries (scenario 5) and non-endemic districts of all countries (scenario 6) remained the same (5%), requiring the endemic districts in high endemic countries to reduce their coverage to accommodate the supply constraint.

The allocation of vaccine doses is staggered over a three-year period for preventive vaccination campaigns, to make the strategies more realistic in terms of the number of vaccine doses required (**Table C.3**). For scenarios only targeting high endemic countries, all vaccine is allocated to Liberia, Guinea and Sierra Leone in the first year, while doses are allocated over three years to Nigeria to reflect a more realistic rollout of the very large number of doses required. For scenarios including preventive vaccination in all countries, vaccine is first allocated to high endemic countries in the first year, followed by medium endemic countries in the second year and low endemic ones in the third year. Populations are thus allocated preventive vaccine in years 1, 2 or 3, with booster doses administered in years 6, 7 or 8, respectively, where introduction patterns for the second dose are identical to the first dose.

***Table C.2. Summary of Lassa vaccination scenarios and dose allocation.*** *Scenario 1 includes outbreak response vaccination only, while scenarios 2 through 6 include preventive vaccination in addition to outbreak response vaccination. For outbreak response, a stockpile of 1,000,000 doses is made available annually across West Africa (distributed evenly to each district relative to its population size),[18] to be used reactively in response to local surges in Lassa fever cases (see* ***Appendix C.3****). This represents enough doses to vaccinate up to 0.2% of the population in each district each year. Scenarios 2 through 6 include additional doses allocated in the form of preventive vaccination. Preventive vaccination in scenarios 2 – 4 is unconstrained, i.e. the number of doses reflects desired coverage levels in targeted districts. Scenario 5 and 6 are constrained by an upper limit in the total number of doses to reflect an assumed limited global vaccine stockpile. The small vaccine pool reserved for reactive vaccination is available to all countries from year 1, while reactive vaccination is rolled out to different districts in different years (detailed further in* ***Table C.3****). To account for vaccine wastage, 90% of allocated doses are assumed to be delivered (i.e. the number of doses ultimately delivered in all scenarios is reduced by 10% relative to doses allocated).*

| **Scenario number** | **Scenario description** (vaccine coverage as % of population) | **Further details** (applied vaccine coverages) | **Preventive doses allocated** (in 3 years) | **Reactive doses allocated** (in 3 years) | **Total doses allocated** (in 3 years) | **Constrained** (doses limited) |
|---|---|---|---|---|---|---|
| **1** | Outbreak response only (0.2%) | Reactive vaccination only.<br>Same coverage in each district where outbreak identified.<br>Applied reactive coverage = 0.002*0.9 (i.e. 0.2% reduced by 10% wastage).<br>0.002 = reactive dose limit/ total population = 1,000,000/402,625,271. | 0 | 2,415,752 | 2,415,752 | Yes |
| **2** | Endemic districts (80%) | Preventive vaccination of populations in endemic districts followed by reactive vaccination (as in Scenario 1) if outbreak response triggered.<br>Applied preventive coverage = 0.8*0.9 in endemic districts and zero elsewhere. | 31,690,868 | 2,415,752 | 34,106,620 | No |
| **3** | Endemic districts (80%) + non-endemic districts of high endemic countries (5%) | Preventive vaccination of populations in endemic districts and non-endemic districts in high endemic countries followed by reactive vaccination (as in Scenario 1) if outbreak response triggered.<br>Applied preventive coverage = 0.8*0.9 in endemic districts, 0.05*0.9 in non-endemic districts in high endemic countries and zero elsewhere. | 41,337,902 | 2,415,752 | 43,753,654 | No |
| **4** | Endemic districts (80%) + non-endemic districts of all countries (5%) | Preventive vaccination of populations in endemic districts and non-endemic districts followed by reactive vaccination (as in Scenario 1) if outbreak response triggered.<br>Applied preventive coverage = 0.8*0.9 in endemic districts and 0.05*0.9 in non-endemic districts. | 49,841,452 | 2,415,752 | 52,257,204 | No |
| **5** | Endemic districts (55%) + non-endemic districts of high endemic countries (5%) | Preventive vaccination of populations in endemic districts and non-endemic districts in high endemic countries followed by reactive vaccination (as in Scenario 1) if outbreak response triggered.<br>Applied preventive coverage = 0.55*0.9 in endemic districts, 0.05*0.9 in non-endemic districts in high endemic countries and zero elsewhere. | 31,434,506 | 2,415,752 | 33,850,257 | Yes |
| **6** | Endemic districts (32.5%) + non-endemic districts of all countries (5%) | Preventive vaccination of populations in endemic districts and non-endemic districts followed by reactive vaccination (as in Scenario 1) if outbreak response triggered.<br>Applied preventive coverage = 0.325*0.9 in endemic districts and 0.05*0.9 in non-endemic districts. | 31,024,999 | 2,415,752 | 33,440,751 | Yes |

***Table C.3. Vaccine doses allocated by country and year.*** *Vaccine doses for preventive campaigns are rolled out to different countries in different years. Generally, vaccines are delivered to countries classified as high-, medium- and low-endemic in years 1, 2 and 3, respectively. However, Nigeria's coverage is divided evenly across the three years to reflect the country's campaign history for other infectious diseases and to reduce totals in the first year. A map showing which countries and districts are classified as "high", "medium" and "low" endemic is given in* ***Figure 1*** *in the main text. The vaccine doses listed here are the doses available to each country and year. Ultimately, 90% of allocated doses are delivered, representing 10% wastage.*

| | **Scenario** | **2** | **3** | **4** | **5** | **6** |
|---|---|---|---|---|---|---|
| **Year** | **Country** | | | | | |
| **1** | Liberia | 1,197,891 | 1,342,334 | 1,342,334 | 967,993 | 631,086 |
| | Guinea | 3,761,060 | 4,125,926 | 4,125,926 | 2,950,594 | 1,892,796 |
| | Sierra Leone | 2,412,597 | 2,593,359 | 2,593,359 | 1,839,423 | 1,160,880 |
| | Nigeria | 8,106,440 | 11,092,095 | 11,092,095 | 8,558,832 | 6,278,896 |
| | **Year 1 Total** | **15,477,987** | **19,153,713** | **19,153,713** | **14,316,842** | **9,963,658** |
| **2** | Benin | | | 625,285 | | 625,285 |
| | Côte d'Ivoire | | | 1,289,798 | | 1,289,798 |
| | Ghana | | | 1,604,533 | | 1,604,533 |
| | Togo | | | 416,782 | | 416,782 |
| | Nigeria | 8,106,440 | 11,092,095 | 11,092,095 | 8,558,832 | 6,278,896 |
| | **Year 2 Total** | **8,106,440** | **11,092,095** | **15,028,493** | **8,558,832** | **10,215,294** |
| **3** | Burkina Faso | | | 1,097,028 | | 1,097,028 |
| | Gambia | | | 115,302 | | 115,302 |
| | Guinea-Bissau | | | 90,570 | | 90,570 |
| | Mali | | | 1,110,207 | | 1,110,207 |
| | Mauritania | | | 215,427 | | 215,427 |
| | Niger | | | 1,148,218 | | 1,148,218 |
| | Nigeria | 8,106,440 | 11,092,095 | 11,092,095 | 8,558,832 | 6,278,896 |
| | Senegal | | | 790,399 | | 790,399 |
| | **Year 3 Total** | **8,106,440** | **11,092,095** | **15,659,247** | **8,558,832** | **10,846,048** |
| | **Grand Total** | **31,690,868** | **41,337,902** | **49,841,452** | **31,434,506** | **31,024,999** |

#### *C.3. Simulating vaccination by pruning infections*

*Summary*

Vaccination is applied in the model by retrospectively "pruning" zoonotic infections and ensuing person-to-person transmission chains (the baseline "unpruned" incidence data) to generate the number of infections averted due to vaccination. When vaccination prevents infection (including zoonotic infection) and therefore onward transmission, the probability of any infection being pruned is proportional to the share of the population that has already been vaccinated at the time of infection. For scenarios where reactive vaccination and preventive vaccination both occur, we implement preventive vaccination first. Since vaccine-induced immunity is assumed to last for five years and booster doses are assumed to extend immunity to ten years (the duration of our simulations), any vaccine-induced immunity acquired in previous years is carried forward and applied before additional vaccination.

*Details*

For a particular simulation $\varepsilon$, let $U_{d,y}$ represent the total number of "unpruned" LASV infections occurring at baseline in the absence of vaccination in each district $d$ and year $y$.

The number of infections averted by preventive vaccination $P_{v,d,y}$ for a given vaccine strategy $v$ depends on the relevant coverage of preventive vaccination $C^P_{v,d,y}$ (i.e. the proportion of the population vaccinated preventively at the start of year *y* after accounting for dose wastage) and the efficacy of vaccine against infection $VE_{infect}$.

In the first year of simulation, no immunity has accrued and the number of infections averted by preventive vaccination is calculated as

$$P_{v,d,1} = U_{d,1} \times C^P_{v,d,1} \times VE_{infect}$$

We next account for additional onward infections caused by human-to-human transmission. To estimate the number of these additional onward infections averted by preventive vaccination, $O_{v,d,1}$, for computational feasibility we use an approximate approach where the number of infections averted is multiplied by $R_0$, i.e. the average number of secondary infections those primary infections caused and which must also be pruned. The first generation $g$ of additional onward infections averted following preventive vaccination is thus approximated as

$$O^g_{v,d,1} = P_{v,d,1} \times R_0 \times \left(1 - \left(C^P_{v,d,1} \times VE_{infect}\right)\right)$$

where $1 - \left(C^P_{v,d,1} \times VE_{infect}\right)$ represents the proportion of the population who are susceptible (either not vaccinated or not protected by the vaccine) and hence ensures that only onward infections averted in unprotected individuals are counted.

To account for the next generation of additional human-to-human transmission $g + 1$, this process is repeated as

$$O^{g+1}_{v,d,1} = O^g_{v,d,1} \times R_0 \times \left(1 - \left(C^P_{v,d,1} \times VE_{infect}\right)\right)$$

until generation $G$, defined as the first generation at which $O^g_{v,d,1} \leq 1$, giving the total number of onward infections averted as

$$O_{v,d,1} = \sum_{g=1}^{G} O_{v,d,1}^{g}$$

We next account for reactive vaccination, where we use an algorithm (28-day rolling sum) to evaluate over all days $t$ to see if the number of infections remaining in each district (after pruning infections averted by preventive vaccination) exceeds a threshold of 50 infections in 28 days,

$$\sum_{t-27}^{t} U_{d,1,t} - P_{v,d,1,t} - O_{v,d,1,t} > 50$$

If this threshold is met or exceeded on day $T$, reactive vaccination is applied on day $T + 1$, adding additional immunity to the population in that district. The number of infections averted by reactive vaccination $R_{v,d,1}$ is calculated, as above, by multiplying the number of infections in that year by reactive vaccine coverage $C_{v,d,1}^{R}$ and $VE_{infect}$. However, reactive vaccination coverage is zero when $t \leq T$, so

$$R_{v,d,1} = \sum_{t=T+1}^{t=365} U_{d,1,t} \times C_{v,d,1,t}^{R} \times VE_{infect}$$

where we assume that reactive vaccination is only administered to individuals not previously vaccinated.

The number of additional onward human-to-human infections averted by vaccination is then re-calculated to include both preventive and reactive vaccination. The number in the first generation $j$ averted following both preventive and reactive vaccination is approximated as

$$Q_{v,d,1}^{j} = \sum_{t} \left(P_{v,d,1,t} + R_{v,d,1,t}\right) \times R_0 \times \left(1 - \left(\left(C_{v,d,1}^{P} + C_{v,d,1,t}^{R}\right) \times VE_{infect}\right)\right)$$

where $R_{v,d,1,t} = C_{v,d,1,t}^{R} = 0$ when $t < T + 1$.

The next generation of additional human-to-human transmission $j + 1$ is given by

$$Q_{v,d,1}^{j+1} = Q_{v,d,1}^{j} \times R_0 \times \sum_{t} \left(1 - \left(\left(C_{v,d,1}^{P} + C_{v,d,1,t}^{R}\right) \times VE_{infect}\right)\right)$$

where $C_{v,d,1,t}^{R} = 0$ when $t < T + 1$.

The total over all subsequent generations $\{j, j + 1, \ldots, J\}$ is calculated as above as

$$Q_{v,d,1} = \sum_{j=1}^{J} Q_{v,d,1}^{j}$$

where $J$ corresponds to the first generation at which $Q_{v,d,1}^{j} \leq 1$.

For subsequent years ($y > 1$), these steps are repeated but it is also necessary to account for the accruing of vaccine-induced immunity from previous years. Our model assumes stable incidence of LASV spillover over each year of simulation, so it follows that the same approximate number of individuals protected from infection in year $y$ should also be protected in year $y + 1$ in the absence of additional vaccination and assuming no waning of vaccine-induced immunity. (We assume that immunity lasts for five years and that booster doses are administered five years after each individual's first dose, so in our model immunity is effectively maintained among vaccinated individuals for all of simulation time.)

The degree of immune coverage remaining in the population in the second year $I_{v,d,2}$, i.e. the share of infections averted due to vaccination in the first year (reflecting effective vaccination and hence immunity), depends on the corresponding coverages of preventive and reactive vaccination in year 1 and is calculated as

$$I_{v,d,2} = C^{P}_{v,d,1} + C^{R}_{v,d,1}$$

For subsequent years $y > 2$, immune coverage continues to accumulate, such that the degree of immunity in year $y$ is calculated as

$$I_{v,d,y} = C^{P}_{v,d,y-1} + C^{R}_{v,d,y-1} + I_{v,d,y-1}$$

The total number of infections averted due to pre-existing immunity $S_{v,d,y}$ is thus calculated as

$$S_{v,d,y} = U_{d,y} \times I_{v,d,y} \times VE_{infect}$$

where

$$I_{v,d,1} = 0$$

After calculating the number of infections averted due to immunity, we repeat the steps above for any additional preventive and reactive vaccination administered in year $y$. As above, the number of infections averted by preventive vaccination is calculated as

$$P_{v,d,y} = U_{d,y} \times C^{P}_{v,d,y} \times VE_{infect}$$

However, the first generation $g$ of additional onward infections averted following preventive vaccination is now approximated as

$$O^{g}_{v,d,y} = \left(S_{v,d,y} + P_{v,d,y}\right) \times R_0 \times \left(1 - \left(\left(I_{v,d,y} + C^{P}_{v,d,y}\right) \times VE_{infect}\right)\right)$$

with

$$O^{g+1}_{v,d,y} = O^{g}_{v,d,y} \times R_0 \times \left(1 - \left(\left(I_{v,d,y} + C^{P}_{v,d,y}\right) \times VE_{infect}\right)\right)$$

and, as above,

$$O_{v,d,y} = \sum\nolimits_{g=1}^{G} O^{g}_{v,d,y}$$

In turn, the rolling daily sum trigger for reactive vaccination is also updated to account for shrinking outbreak sizes as a result of accruing immunity,

$$\sum\nolimits_{t-27}^{t} U_{d,y,t} - S_{v,d,y,t} - P_{v,d,y,t} - O_{v,d,y,t} > 50$$

Once triggered, the number of infections averted by reactive vaccination in year $y$ is calculated as above,

$$R_{v,d,y} = \sum_{t=T+1}^{t=365} U_{d,y,t} \times C^{R}_{v,d,y,t} \times VE_{infect}$$

However, the first generation $j$ of additional onward human-to-human infections averted following preventive and reactive vaccination now also accounts for immunity accrued from previous years and is approximated as

$$Q^{j}_{v,d,y} = \sum_{t} \left(S_{v,d,y,t} + P_{v,d,y,t} + R_{v,d,y,t}\right) \times R_0 \times \left(1 - \left(\left(I_{v,d,y} + C^{P}_{v,d,y} + C^{R}_{v,d,y,t}\right) \times VE_{infect}\right)\right)$$

where $R_{v,d,y,t} = C^{R}_{v,d,y,t} = 0$ when $t < T + 1$.

Again, the next generation of additional human-to-human transmission $j + 1$ is given by

$$Q^{j+1}_{v,d,y} = Q^{j}_{v,d,y} \times R_0 \times \sum_{t} \left(1 - \left(\left(I_{v,d,y} + C^{P}_{v,d,y} + C^{R}_{v,d,y,t}\right) \times VE_{infect}\right)\right)$$

where $C^{R}_{v,d,y,t} = 0$ when $t < T + 1$ and, as above,

$$Q_{v,d,y} = \sum\nolimits_{j=1}^{J} Q^{j}_{v,d,y}$$

Altogether, accounting for preventive vaccination, reactive vaccination and the accumulation of vaccine-induced immunity, the number of LASV infections averted $A_v$ by a given vaccination scenario $v$ across all ten years and 183 districts included in our simulations is calculated as

$$A_v = \sum_{d} \sum_{y} \left(S_{d,v,y} + P_{d,v,y} + R_{d,v,y} + Q_{d,v,y}\right)$$

This pruning process yields a final dataset containing the total number of unpruned infections, pruned infections and infections averted, which are carried forward into the health-economic model (described below) to estimate the health-economic burden of Lassa fever and impacts of vaccination. In addition, in our health-economic model we consider that vaccination can prevent disease without necessarily preventing infection, so it is necessary to count the number of infections occurring in vaccinated individuals $V_{v,d,y}$. This is found as the number of infections that are not pruned due to imperfect vaccine efficacy, incorporating infections in individuals vaccinated preventively,

$$P^{V}_{v,d,y} = U_{d,y} \times C^{P}_{v,d,y} \times (1 - VE_{infect})$$

infections in individuals vaccinated reactively,

$$R^{V}_{v,d,y} = \sum_{t=T+1}^{t=365} U_{d,y,t} \times C^{R}_{v,d,y,t} \times \left(1 - VE_{infect}\right)$$

and infections in individuals vaccinated in previous years who remain unprotected from infection in subsequent years,

$$S^{V}_{v,d,y} = U_{d,y} \times I_{v,d,y} \times \left(1 - VE_{infect}\right)$$

where

$$I_{v,d,1} = 0$$

Altogether, for vaccine scenario $v$, the total number of infections occurring among individuals vaccinated preventively or reactively over all years and districts is calculated as,

$$V_v = \sum_d \sum_y \left(P^V_{v,d,y} + R^V_{v,d,y} + S^V_{v,d,y}\right)$$

This infection pruning process was implemented as an algorithm in R and is available at www.github.com/drmsmith/lassaVac/.

#### Appendix D. Health-economic model

We developed a decision-analytic health-economic model to project the health and economic burden resulting from LASV infection based on outputs from our LASV infection model (**Figure D.1**). These outputs, used as inputs in the health-economic model, include: $U_{d,y}$, the total number of "unpruned" LASV infections at baseline in the absence of vaccination in each district $d$ and year $y$; $A_{v,d,y}$, the number of LASV infections averted by each vaccination strategy $v$; and $V_{v,d,y}$, the number of infections among vaccinated individuals. Additional parameters built into the model include probabilities of different clinical outcomes among LASV-infected individuals, durations of symptoms and hospitalisation, disability weights associated with different health states, and monetary costs. In **Appendix D.1**, we first describe the model and its outcomes. In **Appendix D.2**, we describe calculation of outcomes averted across considered vaccination scenarios. In **Appendix D.3**, we describe in detail the parameter values and distributions used as model inputs. Finally, in **Appendix D.4** we describe our simulation approach, the calculation and reporting of outcome distributions, and sensitivity analyses. All components of this model are implemented in R and are available at www.github.com/drmsmith/lassaVac/.

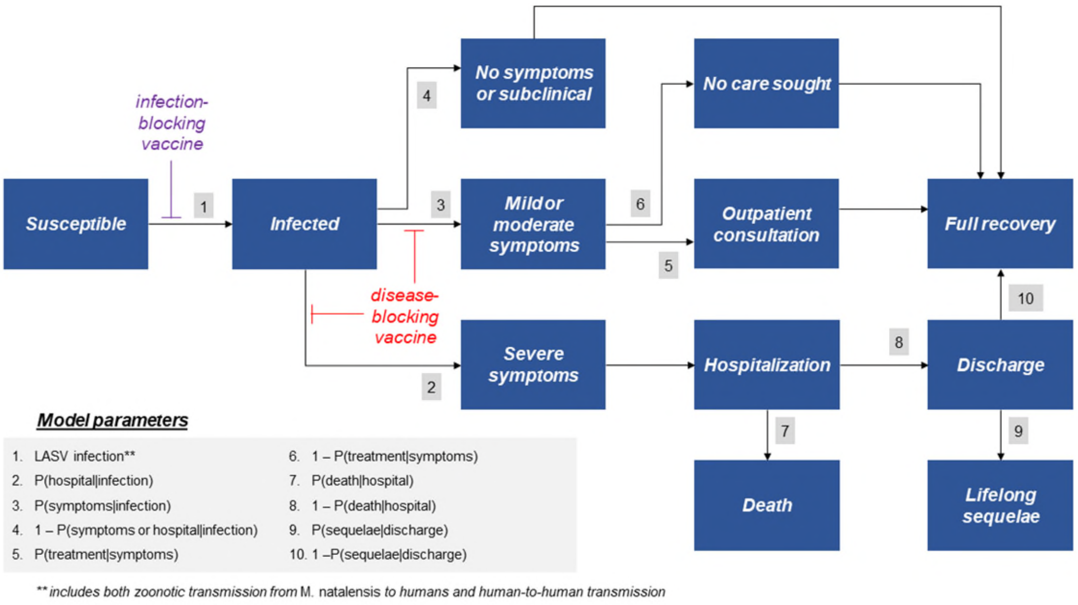


***Figure D.1. Schematic of the health-economic model.*** *Model parameters (transition probabilities) are described further in* ***Table D.1****. Transition probabilities equal 1 for arrows with no parameter listed. LASV: Lassa virus.*

#### *D.1. Model structure and outcomes*

This section describes the health-economic model used to estimate Lassa fever burden. More details on the parameters and data sources used as model inputs are provided in **Appendix D.3**.

LASV infection data were post-processed to assign ages to each infection using country-specific population pyramids assuming no association between age and infection risk in the absence of clear evidence that this risk differs by age,

$$U_{d,y} = \sum_{a=0}^{100} U_{d,y,a} \times \frac{N_{c,a}}{N_c}$$

where $N_{c,a}$ is the number of people of age a $a$ in country $c$, considering ages ranging from 0 to 99 and binning together all individuals ≥100 years old.

Among infected individuals, we consider three degrees of infection severity: asymptomatic/subclinical infection, mild/moderate disease and severe disease, where severe disease is defined as cases severe enough to result in hospitalisation. As fever is the primary symptom of unhospitalised mild/moderate symptomatic cases, the total number of cases in each district, year and age group is given by

$$N_{d,y,a}^{fever} = U_{d,y,a} \times P(symptoms|infection)$$

of whom a share seek outpatient treatment in the community,

$$N_{d,y,a}^{outpatient} = N_{d,y,a}^{fever} \times P(treatment|symptoms)$$

The number of Lassa fever hospitalisations is

$$N_{d,y,a}^{hospital} = U_{d,y,a} \times P(hospital|infection)$$

of whom a share die as a result of their infection,

$$N_{d,y,a}^{death} = N_{d,y,a}^{hospital} \times P(death|hospital)$$

Finally, among those discharged from hospital, a share go on to develop life-long sequelae in the form of sensorineural hearing loss,

$$N_{d,y,a}^{sequelae} = \left(N_{d,y,a}^{hospital} - N_{d,y,a}^{death}\right) \times P(sequelae|discharge)$$

We next measure impacts of Lassa fever on overall disease burden. The disability-adjusted life year (DALY) is a synthetic indicator for measuring health effects generically, ranging from 0 to 1 (0 equals no health effect and 1 equals one year of healthy life lost). Developed by the World Bank in the early 90s, it is one of the most common metrics for estimating health impacts.[19] The DALY incorporates a measure of disease burden, combining disability weights $dw$, which represent health disutility associated with disability on a scale of 0 to 1, as well as years of life lost prematurely due to disease. The number of DALYs associated with each health state, $DALY^{\omega}$, is scaled according to the duration of time associated with the health impact, $dur^{\omega}$.

The number of DALYs due to mild/moderate symptomatic Lassa fever is calculated as

$$DALY^{fever} = \sum_{d}\sum_{y}\sum_{a} N_{d,y,a}^{fever} \times \frac{dw^{fever}}{365} \times dur^{fever}$$

The number of DALYs due to severe hospitalised Lassa fever is the sum of DALYs among those who survive and those who die from severe Lassa fever, who have different hospital length of stay,

$$DALY^{severe} = \sum_{d}\sum_{y}\sum_{a}\Bigg(\left(N_{d,y,a}^{hospital} - N_{d,y,a}^{death}\right) \times \frac{dw^{severe}}{365} \times \left(dur_{pre-hospital}^{severe} + dur_{hospital\_survived}^{severe}\right) + N_{d,y,a}^{death} \times \frac{dw^{severe}}{365} \times \left(dur_{pre-hospital}^{severe} + dur_{hospital\_died}^{severe}\right)\Bigg)$$

DALYs due to disability (i.e. life-long sequelae) or death both apply over individuals' expected remaining lifespan, so years of life lived with disability $YLD$ and years of life lost $YLL$ are both calculated from the difference between each individual's age $a$ at the time of infection and their country-specific life expectancy at age $a$, $x_{c,a}$,

$$YLD_{c,a} = YLL_{c,a} = x_{c,a} - a$$

These measures are used to calculate the number of DALYs due to chronic sequelae,

$$DALY^{sequelae} = \sum_{d}\sum_{y}\sum_{a} N_{d,y,a}^{sequelae} \times dw^{sequelae} \times YLD_{c,a}$$

and the number of DALYs due to death,

$$DALY^{death} = \sum_{d}\sum_{y}\sum_{a} N_{d,y,a}^{death} \times YLL_{c,a}$$

giving the total number of DALYs due to Lassa fever,

$$DALY^{total} = DALY^{fever} + DALY^{hospital} + DALY^{sequelae} + DALY^{death}$$

We next consider monetary costs associated with Lassa fever treatment and hospitalisation. Among those consulting to outpatient care, treatment costs are calculated from country-specific unit costs of treatment $Unit_{c}^{treatment}$, stratified by costs paid out-of-pocket (OOP) versus those paid for or reimbursed by governments,

$$Cost_{gvt}^{outpatient} = \sum_{d}\sum_{y}\sum_{a} N_{d,y,a}^{outpatient} \times Unit_{c}^{treatment} \times P(gvt\ treatment|symptoms)$$

$$Cost_{OOP}^{outpatient} = \sum_{d}\sum_{y}\sum_{a} N_{d,y,a}^{outpatient} \times Unit_{c}^{treatment} \times \left(1 - P(gvt\ treatment|symptoms)\right)$$

Monetary costs due to Lassa fever hospitalisation are estimated slightly differently, assuming that each hospitalisation is associated with both OOP and government-reimbursed costs, based on country-specific estimates of total Lassa fever hospitalisation costs $Unit_{c}^{hospital}$ and the amount therein paid out of pocket $Unit_{c}^{hospital,OOP}$,

$$Cost_{gvt}^{hospital} = \sum_{d}\sum_{y}\sum_{a} N_{d,y,a}^{hospital} \times \left(Unit_{c}^{hospital} - Unit_{c}^{hospital,OOP}\right)$$

$$Cost_{OOP}^{hospital} = \sum_{d}\sum_{y}\sum_{a} N_{d,y,a}^{hospital} \times Unit_{c}^{hospital,OOP}$$

We also measure the number of instances of OOP Lassa fever hospitalisation costs resulting in catastrophic expenditures, $N^{catastrophic}$, or impoverishing OOP expenditures, $N^{impoverishing}$ as,

$$N_{d,y,a}^{catastrophic} = N_{d,y,a}^{hospital} \times P_c(catastrophic|hospital)$$

$$N_{d,y,a}^{impoverishing} = N_{d,y,a}^{hospital} \times P_c(impoverishing|hospital)$$

Probabilities of these outcomes, $P_c(catastrophic|hospital)$ and $P_c(impoverishing|hospital)$, are estimated from country-specific per-capita estimates of OOP expenditure per Lassa fever hospitalisation combined with country-specific estimated income distributions.

We next consider the monetary value of DALYs caused by Lassa fever, $MDALY$, which is calculated from country-specific estimates of the willingness-to-pay per DALY $m_c$, reflecting the opportunity costs of healthcare spending,

$$MDALY = \sum_{c} DALY_{c}^{total} \times m_c$$

We next consider productivity losses, i.e. reduced economic activity resulting from missed work due to Lassa fever. For each clinical outcome $\omega$, in order to estimate impacts of Lassa fever specifically on workers, we calculate $W$, the number of instances of that outcome occurring in the working population (16 to 65),

$$W_{d,y}^{\omega} = \sum_{a=16}^{65} N_{d,y,a}^{\omega} \times w_c$$

where $w_c$ is the country-specific share of the working-age population that is actively employed.

Productivity loss $PL$ is calculated by multiplying the duration of work missed by country-specific estimates of per-capita gross national income $GNI_c$, and depending on the duration of work missed due to each outcome. For those with mild/moderate symptomatic Lassa fever, productivity loss is equal to

$$PL^{fever} = \sum_{d}\sum_{y} W_{d,y}^{fever} \times dur^{fever} \times \frac{GNI_c}{365}$$

and for those hospitalised with severe Lassa fever, productivity loss is equal to

$$PL^{hospital} = \sum_{d}\sum_{y} \Big( \big(W_{d,y}^{hospital} - W_{d,y}^{death}\big) \times \big(dur_{pre-hospital}^{severe} + dur_{hospital\_survived}^{severe}\big) + W_{d,y}^{death} \times \big(dur_{pre-hospital}^{severe} + dur_{hospital\_died}^{severe}\big)\Big) \times \frac{GNI_c}{365}$$

Productivity loss due to Lassa fever mortality depends on each individual's number of years of work lost $YWL$ due to premature mortality, i.e. the number of years between that person's age $a$ at death and the retirement age $r$,

$$YWL_a = r - a$$

Productivity loss due to mortality is thus,

$$PL^{death} = \sum_{d}\sum_{y}\sum_{a} W_{d,y,a}^{death} \times YWL_a \times GNI_c$$

Together, we calculate societal costs $SC$, the total direct monetary cost borne by society as a result of Lassa fever, as the sum total of treatment costs and productivity losses,

$$SC = Cost_{gvt}^{outpatient} + Cost_{OOP}^{outpatient} + Cost_{gvt}^{hospital} + Cost_{OOP}^{hospital} + PL^{fever} + PL^{hospital} + PL^{death}$$

All monetary costs $M_y$ incurred in year $y$, including costs of care $Cost_y$, monetised DALYs $MDALY_y$ and productivity losses $PL_y$, are discounted using a standard discrete annual discounting term, giving total monetary costs as

$$M = \sum_{y} \frac{M_y}{(1 + dis)^{y-1}}$$

where costs in the first year of simulation $y = 1$ are not discounted. To correctly estimate the present monetary value of future productivity losses and future DALYs, i.e. those due to a health event in year $y$ but occurring in year $y + n$, we also apply a continuous-time discounting function[20] to future years lived with disability and future years of life lost

$$YLD_{c,a}^{dis} = YLL_{c,a}^{dis} = \frac{1 - e^{-dis(x_{c,a} - a)}}{dis}$$

and to future years of work lost

$$YWL_a^{dis} = \frac{1 - e^{-dis(r-a)}}{dis}$$

In sensitivity analysis, we calculate monetary costs without applying discounting.

Finally, we consider an additional measures of the value of life lost due to Lassa fever mortality, the value of statistical life $VSL$. This is based simply on country-specific estimates of the value per statistical life $vsl_c$,

$$VSL = \sum_{c} N_c^{death} \times vsl_c$$

More details on the data underlying these parameters and their calculation are given in **Appendix D.3**.

#### *D.2. Outcomes averted due to vaccination*

**Appendix D.1** describes the estimation of Lassa fever burden in the absence of vaccination for a particular simulation $\varepsilon$, i.e. using the baseline number of unpruned infections $U_{d,y}$ from the LASV transmission model to calculate downstream health outcomes and economic costs.

To calculate outcomes averted due to vaccination, $N^V$, we first age-distribute the number of infections averted due to vaccination and those occurring in vaccinated individuals,

$$A_{v,d,y} = \sum_{a=0}^{100} A_{v,d,y,a} \times \frac{N_{c,a}}{N_c}$$

$$V_{v,d,y} = \sum_{a=0}^{100} V_{v,d,y,a} \times \frac{N_{c,a}}{N_c}$$

assuming no relationship between age and vaccination.

From these, we estimate the number of mild/moderate symptomatic cases averted by vaccination, accounting both for vaccine efficacy in preventing against infection (through $A$) and vaccine efficacy in protecting vaccinated infected individuals from disease, $VE_{disease}$,

$$N_{v,d,y,a}^{V,fever} = \left(A_{v,d,y,a} + V_{v,d,y,a} \times VE_{disease}\right) \times P(symptoms|infection)$$

Similarly, the number of hospitalisations averted by vaccination is,

$$N_{v,d,y,a}^{V,hospital} = \left(A_{v,d,y,a} + V_{v,d,y,a} \times VE_{disease}\right) \times P(hospital|infection)$$

In the same way that all health-economic outcomes presented in **Appendix D.1** are downstream from either $N^{fever}$ or $N^{hospital}$, all health-economic outcomes averted due to vaccination are calculated downstream from $N^{V,fever}$ and $N^{V,hospital}$ , using the same assumptions and formulae as above.

#### *D.3. Parameter inputs*

This section describes the estimation of parameters used as inputs for the health-economic model. Data synthesised for use in this study were identified through literature review and inputs from subject-matter experts. Parameter distributions used as inputs for Monte Carlo simulation are provided below in **Table D.1**. Parameter calculation and estimation was conducted in R and is available at www.github.com/drmsmith/lassaVac/.

##### *Lassa fever symptoms, severity and care-seeking*

The probability of LASV infection resulting in mild/moderate symptomatic Lassa fever, $P(symptoms|infection)$, was estimated using prospective cohort data from Sierra Leone, in which 9 of 48 individuals with detected LASV seroconversion experienced temporally related fever.[21] These data come from four different villages and were meta-analyzed using the inverse variance method and a generalised linear mixed model. These data are limited by a small sample size and date from the 1980s. Future data from the ongoing ENABLE study may soon allow for better refined estimates.[22]

Due to poor diagnostic capacity in affected regions and low ascertainment of mild Lassa fever cases in community settings, data describing outpatient care-seeking for Lassa fever are unavailable. Where parameter estimates specific to Lassa fever are unavailable, we use malaria as a proxy as these diseases are known to have significant overlap in the presentation of mild cases. The probability that individuals with mild/moderate symptomatic Lassa fever seek outpatient care, $P(treatment|symptoms)$, and the probability that they seek government-reimbursed outpatient care specifically, $P(gvt\ treatment|symptoms)$, are thus taken from modelled estimates of treatment-seeking for symptomatic malaria in West Africa.[23]

The probability of LASV infection resulting in hospitalisation, $P(hospital|infection)$, was estimated by dividing the annual number of confirmed hospitalised Lassa fever cases in two states of Nigeria (Edo and Ondo) between 2018 and 2021 by the annual number of LASV infections estimated in those states by our infection model. These states were chosen as they are known to have both high Lassa fever incidence and robust Lassa fever surveillance, in particular subsequent to substantial improvement and expansion of testing capacity since 2018.[24,25] The number of hospitalisations was taken from a literature review comprising 38 records of cases and fatalities.[26] The probability of LASV hospitalisation resulting in death, $P(death|hospital)$, is estimated from these same data using the weighted mean of the hospital case-fatality rate (CFR) over these years and states. Our estimated mean 16.1% (95% CI: 6.5%-29.0%) is consistent with interim data from the prospective ENABLE study (16%) and a recent estimate (12%) from a prospective cohort study in Owo, Nigeria,[27,28] while considerable uncertainty may be consistent with true heterogeneity in this parameter, which is likely to vary across settings depending on local diagnostic capacity, treatment protocols and the availability of dialysis and other clinical resources. To estimate the risk of hospitalisation upon infection, we divided the number of hospitalisations by the number of infections in those same districts as predicted by our infection model, resulting in a mean infection-hospitalisation risk of 0.9%. These estimates were subsequently applied to predicted infections in all countries, including those with poor surveillance.

Sensorineural hearing loss is known to affect a substantial share of survivors of viral haemorrhagic fever, and the probability of Lassa fever patients developing hearing loss following hospital discharge is included as $P(sequelae|discharge)$. In preliminary data from the ENABLE study, 13 of 21 (62%) Lassa fever survivors reported sensorineural hearing loss during follow-up at four months, although audiometry results were available for only a subset of patients.[27] However, in a case-control study among Lassa fever survivors in Sierra Leone by Ficenec et al., only 8/47 (17%) reported hearing loss between 3 months and 3 years post-infection, with audiometry results again only available for a subset of patients.[29] Due to small sample sizes and considerable divergence in these two estimates, we used the estimate from the ENABLE study in our primary analysis and the one from Ficenec et al. in a sensitivity analysis.

For the duration of fever among individuals with mild/moderate symptomatic Lassa fever, $dur^{fever}$, we used recent estimates of the duration of fever among symptomatic malaria cases in Indonesia.[30] All other duration parameters pertaining to Lassa fever hospitalisation were taken from the LASCOPE study,[28] with their distributions estimated from quantiles using the method proposed by Wan et al.[31]

##### *Disability weights and DALY monetization*

We estimate DALYs by assigning disability weights to Lassa fever patients based on Lassa-associated clinical symptoms identified in a systematic review and health state disutility values from Global Burden of Disease estimates and other studies. For mild/moderate symptomatic Lassa fever, $dw^{fever}$, we used the disability weight associated with fever and aches from the Global Burden of Disease (GBD) study.[32] Estimates of disability weights associated with severe Lassa fever and resulting sequelae are unavailable, so we developed our own estimates from data in the literature, accounting for a high degree of uncertainty.

To estimate a disability weight for severe Lassa fever, $dw^{severe}$, we first reviewed the literature for evidence on clinical symptoms. Merson et al. (2021) report the most comprehensive systematic review of symptoms in hospitalised cases to date.[33] We assigned disability weights to each of these symptoms using GBD data where possible,[32] and supplemented with data from other studies where necessary.[34-36] We then simulated joint symptom profiles for 100,000 theoretical Lassa fever patients by sampling each symptom independently from the list of identified symptoms based on the proportion of patients presenting with that symptom at "baseline" and "post-baseline", as reported by Merson et al. Due to uncertainty in how disability weights combine in patients simultaneously experiencing multiple symptoms, for each patient we conservatively considered the symptom having the greatest disutility as being representative of that patient's overall disutility, with one estimate at baseline and one estimate post-baseline. Estimates for each patient were averaged across the two time-points to generate a non-parametric distribution of 100,000 disability weights associated with Lassa fever hospitalisation, from which 100 estimates were randomly sampled for inclusion in Monte Carlo simulations. We applied $dw^{severe}$ across the full duration of each individual's severe disease, including both the duration of hospitalisation and the duration of illness prior to hospitalisation.

Due to great uncertainty in the severity of Lassa fever-induced sensorineural hearing loss, to estimate an associated disability weight $dw^{sequelae}$ we used the severity distribution of all-cause hearing loss from GBD data from Nigeria, i.e. the proportion of those with hearing loss having mild, moderate, moderate-severe, severe, profound or complete hearing loss.[37] We then coupled each level of severity with its associated disability weight (for hearing loss with ringing), and simulated disutility in 100,000 theoretical patients based on these prevalence proportions. This resulted in a highly right-skewed disutility distribution due to the high prevalence of mild and moderate hearing loss relative to profound and complete hearing loss. We randomly sampled 100 estimates from this distribution for inclusion in Monte Carlo simulations.

DALYs were monetised using country-specific health opportunity costs estimated by Ochalek et al.[38] Specifically, we made use of the provided country-specific percentage of GDP per capita estimate that underlies the DALY-4 estimation method, multiplying the total per-individual DALY value by a specific proportion of the GDP per capita. We note that country-specific DALY monetization, reported in 2021 International dollars (Int$), ranged from Int$95/DALY in Guinea-Bissau to Int$851/DALY in Ghana, with lower average estimates across high-endemic countries (Int$=223/DALY) relative to medium- and low-endemic countries (Int$=403/DALY). This explains why a vaccine scenario targeting only high-endemic countries may avert more DALYs but less monetised DALY value than a scenario targeting high-, medium- and low-endemic countries (see vaccine scenarios 5 and 6 in **Table 2** in the main text).

##### *Treatment costs*

Due to limited data on Lassa fever patients in outpatient settings, for unit costs of outpatient treatment $Unit_c^{treatment}$ we used mean country-specific unit cost estimates of all-cause outpatient visits measured in 2017 International dollars.[39] Importantly, unlike commonly used WHO CHOICE estimates, these estimates account for ancillary services related to outpatient visits such as diagnostics and medications. We adjusted these estimates for inflation to Int$ 2021 using the US dollar inflation rate (i.e. using the quotient of World Bank GDP deflator values for 2021 over 2017).[40]

Unit costs for Lassa fever hospitalisation and associated OOP expenditures are based on data from Asogun et al. (2016), who reported the medical cost of Lassa fever treatment in a specialist teaching hospital in Irrua, Nigeria.[41] In this study, the average total treatment costs were ₦205,559, and the associated average total OOP expenditures were ₦86,803 (in 2016 Naira). We first took the local currency values of the treatment costs and OOP expenditures and converted these to Int$ value using the exchange rate (purchasing power parity (PPP) conversion factor ) at the time of costing (2016) and inflated them to Int$ 2021, as above. The resulting cost estimate for Lassa fever hospitalisation in Nigeria is $Unit_{NGA}^{hospital} = \text{Int\$2,193}$, of which $Unit_{NGA}^{hospital,OOP} = \text{Int\$926}$ is paid OOP.

We assumed the same total Lassa fever hospitalisation unit cost in all countries, but adjusted the cost paid OOP using World Bank data on country-specific proportions of healthcare expenditures.[40] For example, the proportion of per-capita healthcare expenditures paid OOP in Nigeria (2019) was 70.5%. However, in Asogun et al. the proportion of OOP expenditures for Lassa given total treatment costs was 42.2%. Based on this, we calculated an adjustment factor of 0.5986 by dividing the Nigerian study proportion by the World Bank proportion. Using this factor, we then translated Nigerian study-specific treatment costs to OOP expenditures in other settings. For example, the proportion of per-capita healthcare expenditures paid OOP in Benin (2019) was 47%. Multiplying the average total treatment costs from the Nigerian study (Int$2,193) by the product of the adjustment factor (0.5986) and the proportion of OOP expenditures (47%) resulted in a Benin-specific OOP expenditure estimate $Unit_{BEN}^{hospital,OOP} = \text{Int\$617}$.

##### *Risk of catastrophic / impoverishing healthcare expenditures*

Catastrophic healthcare expenditures are defined based on Sustainable Development Goal 3.8.2 (catastrophic health spending), which outlines that a "population with household expenditures on health greater than 10% of total household expenditure or income" are at risk of experiencing catastrophic health spending.[42] Estimation of the proportion of individuals in each country at risk of either catastrophic healthcare expenditure resulting from Lassa fever hospitalisation, $P_c(catastrophic|hospital)$, or impoverishing healthcare expenditures, $P_c(impoverishing|hospital)$, is based on estimated country-specific per-capita OOP expenditure for Lassa fever hospitalisation, $Unit_c^{hospital,OOP}$.

To estimate these parameters, first we extrapolated total annual income by assuming that estimated OOP expenditures represent ten percent of the annual income. Second, we estimated daily income by dividing the estimated total annual income by 365 days. Third, we used the World Bank Poverty and Inequality Platform (WB PIP) to estimate the number of individuals that live below a specific income threshold, where the income threshold was defined as the estimated daily income.[43] Fourth, given total population count, we estimated the proportion of individuals that fall below the defined income threshold. This proportion represents the number of individuals at risk of catastrophic healthcare expenditures. Fifth, we repeated this exercise, however, with a changed daily income threshold of 2.15 PPP Int$ (i.e., the international poverty line; in 2019 Int$ PPP), to estimate the number of individuals that live in poverty. To obtain the number of individuals at risk of impoverishing healthcare expenditures, we then subtracted the number of individuals below the poverty line from the number of individuals at risk of catastrophic healthcare expenditures. Due to low unit costs for outpatient care, which ranged from approximately Int$6 in Guinea to Int$21 in Nigeria, we assumed no contribution of mild disease to catastrophic or impoverishing healthcare expenditures.

##### *Productivity losses*

Productivity losses resulting from days of work missed due to Lassa fever were quantified using gross national income per capita. Country-specific estimates of per-capita gross national income in Int$ 2021 ($GNI_c$) underlying productivity loss estimation were sourced from the World Bank.[40] Data from the International Labour Organisation were used to define the share of the working-age population active in the workforce in each country, $w_c$.[44] Due to the lack of data on projected wage growth in the countries of interest, wages (proxied through GNI) were held constant over the time horizon of the analysis.

##### *Value of statistical life*

The conceptual idea underlying the value of statistical life (VSL) method of life valuation is to estimate an individual's willingness to pay for a reduction in the probability of a risk (i.e., the risk of dying), aggregated at the population level to represent the demand for collective risk-reduction.[45] To produce global VSL estimates, we applied the methodology developed in 2019 by a global expert group of benefit-cost researchers.[46] The reference point of this approach is the recently updated United States of America (USA) VSL estimate of $12.3 million (2022 US$).[47] Due to the limited availability of direct VSL estimates in low- and middle-income countries, we applied the value-transfer method outlined in the Reference Case Guidelines for Benefit-Cost Analysis in Global Health and Development.[46] We used the following equation to compute the VSL of all countries $c$ in our model:

$$VSL_c = VSL_{USA} \times (GNI_c \div GNI_{USA})_E$$

where $VSL_c$ is the unknown value for country $c$, $VSL_{USA}$ is the USA value ($12.3 million USD), and $GNI_c$ is the 2021 gross national income per capita (PPP) of country $c$, or the USA, as applicable. Finally, $E$ denotes the income elasticity underlying the VSL, i.e., the percentage change in VSL associated with a 1% change in real income. The value-transfer method adjusts VSL from the USA setting to lower-income settings based on the income ratio between the target and reference countries. The ratio is raised to the income elasticity for the value of reducing mortality risk, which is estimated at 1.2 for middle-income countries and 1.5 for low-income countries (other high-income countries have an income elasticity of 1.0).

##### *Discounting*

In our baseline estimates, we assume an annual discounting rate of $dis = 3\%$ for future monetary costs, which also necessitates discounting of future years of life lost (YLLs), future years of work lost

(YWLs) and future years of life lived with disability (YLD). In sensitivity analysis, we consider $dis = 0\%$, in which case undiscounted YLLs, YWLs and YLDs are used when calculating costs..

***Table D.1. Lassa fever parameters.*** *Model parameters describing Lassa fever burden in the health-economic model. CI = confidence interval, LASV = Lassa virus.*

| Parameter | Mean [95% CI] | Distribution [parameters] | Notes |
|---|---|---|---|
| ***Lassa fever (clinical probabilities)*** | | | |
| $P(symptoms\|infection)$ | 18.8% [10.0%, 32.3%] | inverse logit-transformed Normal [μ=-1.47, σ=0.37] | Among a prospective cohort of individuals living in four villages in Sierra Leone, the proportion who had fever temporally related to LASV seroconversion.[21] We meta-analyzed these primary data using the inverse variance method and a generalised linear mixed model. |
| $P(treatment\|symptoms)$ | | | Modelled estimates from West Africa of treatment-seeking rates for febrile malaria,[23] fit to Normal distributions. |
| *any treatment* | 59.8% [54.3%, 65.1%] | Normal [μ=0.598, σ=0.028] | |
| *government treatment* | 48.9% [43.5%, 54.5%] | Normal [μ=0.489, σ=0.028] | |
| $P(hospital\|infection)$ | 0.87% [0.58%, 1.12%] | Normal [μ=0.009, σ=0.001] | Estimated as the annual number of confirmed hospitalised Lassa fever cases in Edo and Ondo between 2018 and 2021,[26] divided by the estimated annual number of LASV infections in Edo and Ondo predicted by our model, fit to a Normal distribution. |
| $P(sequelae\|discharge)$ | 61.9% [17.0% in sensitivity analysis] | / | In preliminary data from ENABLE, 13/21 (61.9%) Lassa fever survivors experienced sensorineural hearing loss at 4 months post-hospitalisation.[27] For sensitivity analysis, we considered a case-control study in which 8/47 (17.0%) survivors reported hearing loss over 3 months to 3 years post-hospitalisation.[29] |
| $P(death\|hospital)$ | 16.1% [6.5%, 29.0%] | Beta [α=6.24, β=32.51] | Hospital case-fatality rate estimated as weighted means from surveillance data of hospitalised cases in Edo and Ondo between 2018 and 2021,[26] fit to a Beta distribution. |
| ***Durations of symptoms and hospital stays*** | | | |
| $dur^{fever}$ | 3.53 days [3.29, 3.77] | Normal [μ=3.53, σ=0.12] | Estimated fever duration among 261 symptomatic malaria cases,[30] fit to a Normal distribution. |
| $dur^{severe}_{pre-hospital}$ | 9.33 days [8.95, 9.72] | Normal [μ=9.33, σ=0.20] | Estimates of durations of illness prior to and during Lassa fever hospitalisation among a prospective cohort of 510 patients in Owo, Nigeria.[28] Means and standard errors were estimated from reported quartiles,[31] and fit to Normal distributions. |
| $dur^{severe}_{hospital\_died}$ | 3.33 days [2.40, 4.26] | Normal [μ=3.33, σ=0.47] | |
| $dur^{severe}_{hospital\_survived}$ | 12.00 days [11.66, 12.34] | Normal [μ=12.00, σ=0.17] | |
| ***Disability weights*** | | | |
| $dw^{fever}$ | 0.051 [0.030, 0.072] | Normal [μ=0.051, σ=0.011] | Global Burden of Disease estimate for fever ("has a fever and aches, and feels weak, which causes some difficulty with daily activities),[32] fit to a Normal distribution. |
| $dw^{severe}$ | 0.335 [0.220, 0.458] | Non-parametric | Distribution generated by randomly sampling from Lassa fever symptoms according to their prevalence across early ("baseline") and late ("post-baseline") hospitalisation,[33] and selecting the symptom with the highest disutility from available estimates.[32,34-36] |
| $dw^{sequelae}$ | 0.062 [0.010, 0.302] | Non-parametric | Among Nigerians having all-cause hearing loss, the proportion experiencing different severities multiplied by corresponding disability weights associated with that level of hearing loss (with ringing),[37] which were randomly sampled from Normal distributions. |

##### *D.4. Simulations and sensitivity analysis*

To account for uncertainty in parameter inputs, our final outcomes are estimated using probabilistic sensitivity analysis (PSA), where over $\rho = 100$ Monte Carlo simulations input parameters are randomly drawn from their probability distributions and fed through the model. Distributions of all input parameters varied in the PSA are provided in **Table D.1** and include clinical probabilities, durations of symptoms and hospital stays, and disability weights associated with Lassa fever. The final parameter set used as model inputs is visualised in **Figure D.2 panel A**.

For each vaccination scenario, final outcomes are calculated across all $\varepsilon = 99$ estimates of LASV spillover in all $d = 183$ districts, $y = 10$ years and $\rho = 100$ Monte Carlo parameter sets. Final distributions are reported as the mean and 95% uncertainty interval [2.5% quantile, 97.5% quantile] across all simulations.

To identify the sources of uncertainty in our model outcomes, we conducted a univariate sensitivity analysis, where each input parameter varied in Monte Carlo simulations was varied individually using its minimum or maximum value, while holding all other parameters constant at the distribution mean. Median LASV infection estimates from $\varepsilon = 50$ were used as model inputs for this analysis and 62% of Lassa patients discharged from hospital were assumed to develop hearing loss. Final outcomes are calculated for each parameter varied, and reported as the difference in that outcome relative to a "baseline" simulation using the mean distribution value for all input parameters. Results from the univariate sensitivity analysis for two key model outcomes (total cumulative DALYs and total cumulative societal costs) are presented in **Figure D.2 panels B and C**.

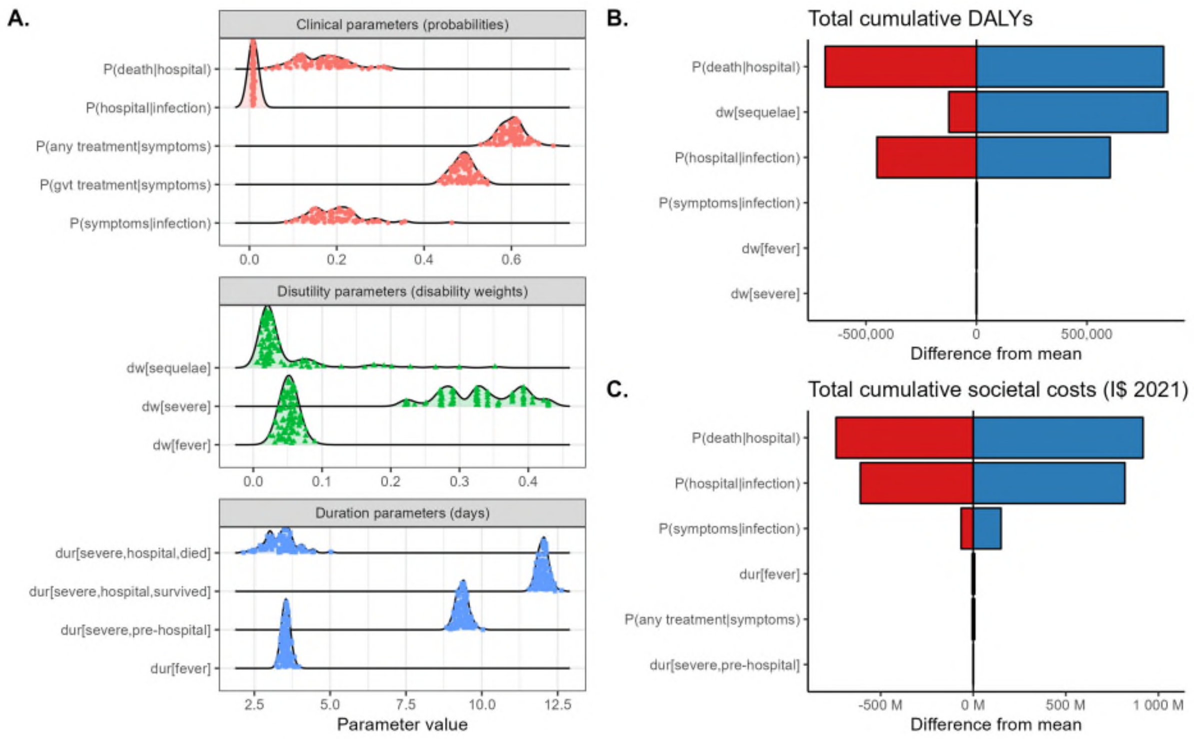


***Figure D.2. Input parameter uncertainty and impact on outcomes.*** *(**A**) Parameter distributions used in Monte Carlo simulations. (**B**) Tornado plot depicting the mean difference in the total cumulative DALYs due to Lassa fever when using the maximum (blue) or minimum (red) value of corresponding parameters (y-axis). (**C**) Tornado plot depicting the mean difference in the total cumulative societal costs due to Lassa fever when using the maximum (blue) or minimum (red) value of corresponding parameters (y-axis).*

#### Appendix E. Supplementary results for Lassa fever

##### *E.1 Health and economic burden in the absence of vaccination*

Most LASV infections (59.0%) occurred in high endemic countries, but there were approximately four-fold more infections in "non-endemic" than "endemic" districts of high endemic countries (**Table E.1**). The vast majority of DALYs were due to mortality, with DALYs due to chronic sequelae making only a small contribution to total DALYs, and those due to acute infection in the community or severe infection in hospital making negligible contribution (**Figure E.1**). Consequently, monetised DALYs were reduced by approximately 9% in the sensitivity analysis considering a lower risk of sensorineural hearing loss after hospital discharge; conversely, undiscounted estimates of monetised DALYs and societal costs were approximately 113% and 56% greater, respectively, than discounted estimates (**Table E.5**).

**Table E.1. Cumulative human LASV infections over ten years in the absence of vaccination.** Infection totals are aggregated according to endemicity classifications. LASV = Lassa virus, UI = uncertainty interval.

| | LASV infections | |
|---|---|---|
| | **mean (95% UI)** | **% of total** |
| **High endemic countries** | | |
| All districts | 16.1M (12.6M-19.7M) | 59.0% |
| Endemic districts | 3.1M (2.5M-3.7M) | 11.3% |
| Non-endemic districts | 13.0M (10.1M-16.0M) | 47.8% |
| **Medium endemic countries** | | |
| All districts | 5.1M (3.9M-6.3M) | 18.6% |
| **Low endemic countries** | | |
| All districts | 6.1M (4.6M-7.7M) | 22.4% |

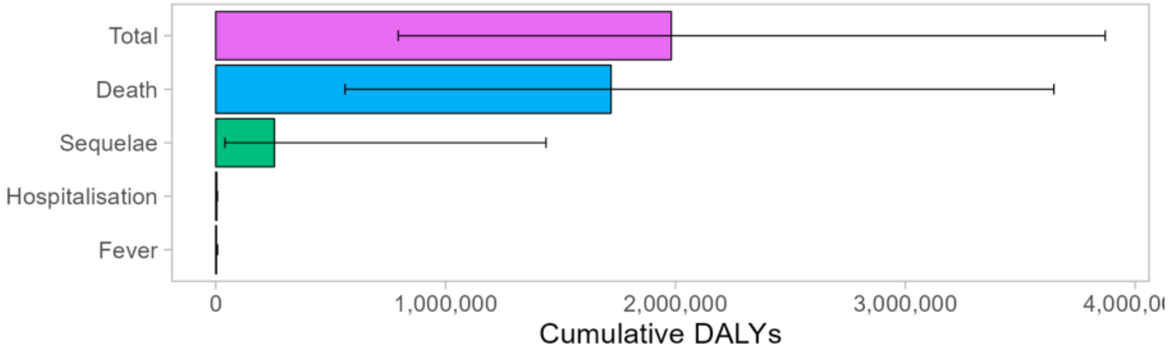


***Figure E.1. Breakdown of Lassa fever DALYs in the absence of vaccination.*** *Summary estimates of DALYs due to Lassa fever over ten years, stratified across mild/moderate symptomatic disease in the community (fever), severe disease (hospitalisation), chronic sensorineural hearing loss (sequelae) and death. Bars represent means and error bars represent 95% uncertainty intervals, for the baseline scenario assuming a probability of sequelae of 62% among patients discharged from hospital. DALY = disability-adjusted life year.*

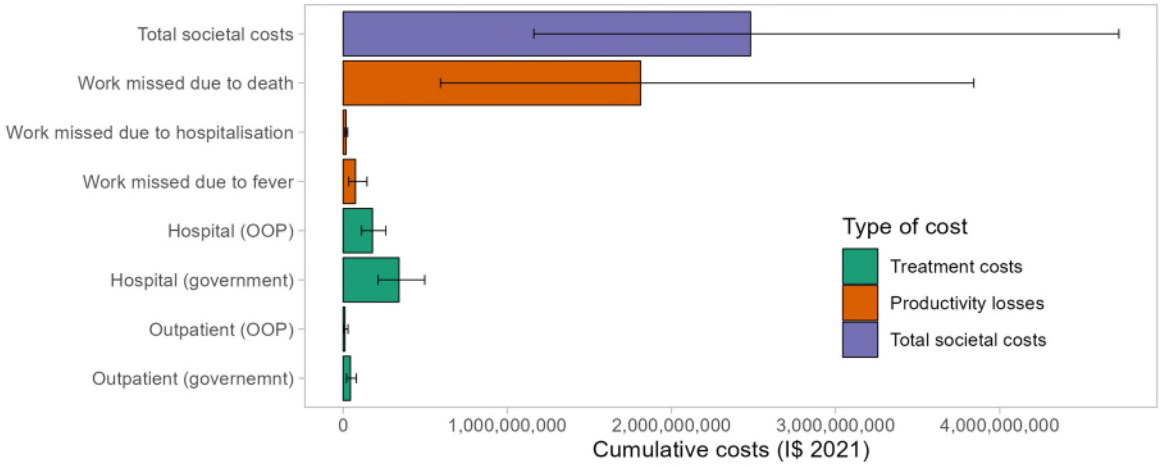


***Figure E.2. Breakdown of societal costs due to Lassa fever in the absence of vaccination.*** *Summary estimates of the total societal costs due to Lassa fever over ten years (purple), stratified across productivity losses (orange) and treatment costs (green). Bars represent means and error bars represent 95% uncertainty intervals, here assuming a discount rate of 0%. OOP = out-of-pocket, I$ = International dollar.*

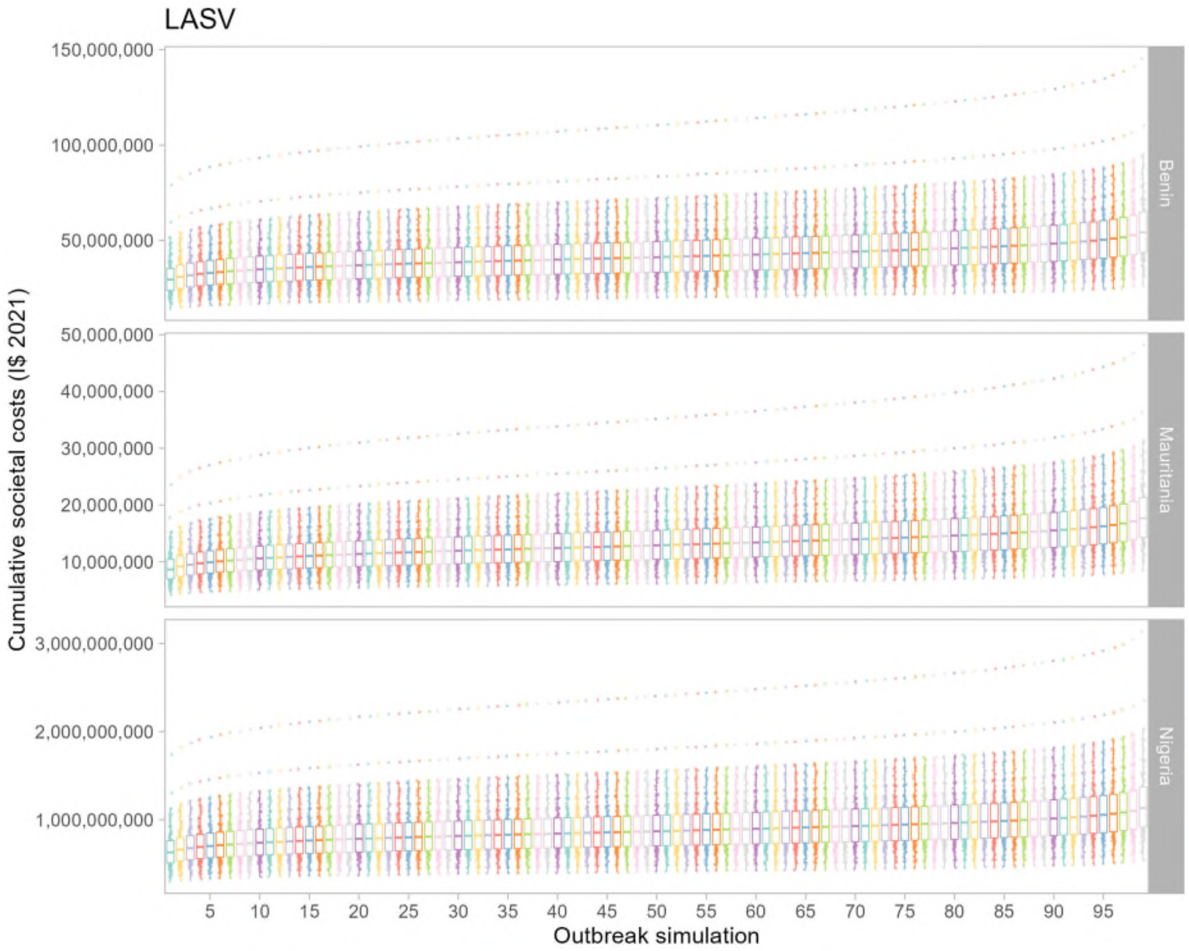


***Figure E.3. Raw Lassa fever simulation output data in absence of vaccination.*** *Raw data overlaid with boxplots depicting estimates of the cumulative societal costs incurred by Lassa fever over ten years across all simulations. The y-axis depicts* $SC_{\varepsilon,\rho,c}$*, an estimate of the cumulative societal costs from LASV outbreak simulation* $\varepsilon$ *(x-axis), Monte Carlo simulation* $\rho$ *(points) and country* $c$ *(panels). Results are shown for three selected countries having high estimated burden of LASV (Nigeria), moderate burden (Benin) or low burden (Mauritania). These estimates correspond to baseline parameter assumptions of a 3% annual discounting rate for future monetary costs and a probability of sequelae of 62% among patients discharged from hospital. Upper and lower boxplot whiskers extend no further than values 150% larger or smaller, respectively, than the interquartile range. I$ = International dollar, LASV = Lassa virus.*

***Table E.2. Cumulative health burden of Lassa fever (per 100,000 population) by country in the absence of vaccination.*** *All figures represent means (95% uncertainty intervals) over ten years across 99 runs of the infection model and 100 runs of the health-economic model, for the baseline scenario assuming a probability of sequelae of 62% among patients discharged from hospital. DALY = disability-adjusted life year, K = thousand.*

| | Infections | Mild/moderate cases | Hospitalisations | Deaths | Sequelae | DALYs |
|---|---|---|---|---|---|---|
| *Benin* | 6.6K (5.1K-8.2K) | 1.3K (647.5-2.4K) | 57.5 (36.0-83.8) | 9.5 (3.1-20.2) | 29.7 (18.1-43.1) | 505.2 (202.5-986.4) |
| *Burkina Faso* | 6.7K (5.1K-8.4K) | 1.3K (651.6-2.4K) | 58.1 (36.2-85.3) | 9.6 (3.1-20.5) | 30.0 (18.3-43.9) | 509.6 (203.2-1.0K) |
| *Côte d'Ivoire* | 6.8K (5.1K-8.6K) | 1.4K (665.6-2.5K) | 59.5 (36.9-87.5) | 9.9 (3.2-21.0) | 30.7 (18.6-45.0) | 500.2 (198.9-984.6) |
| *Ghana* | 6.0K (4.5K-7.6K) | 1.2K (585.3-2.2K) | 52.3 (32.4-77.0) | 8.7 (2.8-18.5) | 27.0 (16.4-39.6) | 454.2 (180.4-894.4) |
| *Guinea* | 7.5K (6.0K-8.9K) | 1.5K (744.2-2.7K) | 64.9 (41.8-92.8) | 10.8 (3.6-22.5) | 33.5 (21.1-47.6) | 569.2 (234.2-1.1K) |
| *Gambia* | 5.0K (3.6K-6.4K) | 990.9 (478.2-1.8K) | 43.3 (26.4-64.3) | 7.2 (2.3-15.4) | 22.4 (13.3-33.2) | 385.6 (151.2-765.9) |
| *Guinea-Bissau* | 4.9K (3.6K-6.3K) | 968.0 (467.5-1.8K) | 42.3 (25.9-62.8) | 7.0 (2.3-15.0) | 21.8 (13.0-32.4) | 358.1 (140.5-710.7) |
| *Liberia* | 6.3K (4.7K-7.9K) | 1.2K (610.4-2.3K) | 54.5 (33.9-79.9) | 9.0 (2.9-19.2) | 28.1 (17.1-41.1) | 467.7 (186.6-918.8) |
| *Mali* | 7.0K (5.3K-8.8K) | 1.4K (684.4-2.5K) | 60.9 (38.1-89.0) | 10.1 (3.3-21.5) | 31.5 (19.2-45.8) | 537.0 (214.6-1.1K) |
| *Mauritania* | 5.4K (4.0K-7.0K) | 1.1K (523.4-2.0K) | 47.4 (28.9-70.4) | 7.9 (2.5-16.8) | 24.5 (14.6-36.3) | 430.0 (168.6-853.7) |
| *Niger* | 7.8K (5.8K-9.9K) | 1.6K (754.9-2.8K) | 67.8 (41.7-100.1) | 11.2 (3.6-24.0) | 35.0 (21.1-51.5) | 622.4 (246.0-1.2K) |
| *Nigeria* | 6.9K (5.3K-8.5K) | 1.4K (676.0-2.5K) | 59.8 (37.7-86.8) | 9.9 (3.3-21.0) | 30.9 (19.0-44.7) | 477.3 (192.2-928.1) |
| *Senegal* | 5.3K (3.9K-6.9K) | 1.1K (513.1-2.0K) | 46.4 (28.4-69.0) | 7.7 (2.5-16.5) | 24.0 (14.3-35.6) | 437.7 (171.8-869.5) |
| *Sierra Leone* | 7.6K (6.3K-8.9K) | 1.5K (759.7-2.7K) | 65.9 (42.8-93.4) | 10.9 (3.6-22.8) | 34.0 (21.6-47.9) | 568.3 (236.5-1.1K) |
| *Togo* | 6.5K (4.9K-8.2K) | 1.3K (637.2-2.4K) | 56.8 (35.4-83.3) | 9.4 (3.1-20.0) | 29.3 (17.9-42.9) | 485.5 (193.7-953.1) |
| **Total** | **6.8K (5.2K-8.4K)** | **1.3K (663.1-2.4K)** | **58.9 (36.9-85.8)** | **9.8 (3.2-20.7)** | **30.4 (18.6-44.1)** | **492.3 (197.1-961.2)** |

***Table E.3. Cumulative economic burden of Lassa fever by country over ten years in the absence of vaccination.*** *All figures represent means (95% uncertainty intervals) across 100 runs of the health-economic model for each of 99 runs of the infection model, for the baseline scenario assuming a probability of sequelae of 62% among patients discharged from hospital. Monetary costs are reported in International dollars (2021) and future costs are discounted at 3%/year. Catastrophic and impoverishing expenditures are reported as the number of individuals facing those costs. DALY = disability-adjusted life year, VSL = value of statistical life, K = thousand, M = million, B = billion.*

| | Treatment costs, government-reimbursed ($ 2021) | Treatment costs, out-of-pocket ($ 2021) | Catastrophic expenditures (N) | Impoverishing expenditures (N) | Productivity losses ($ 2021) | Monetised DALYs ($ 2021) | VSL ($ 2021) |
|---|---|---|---|---|---|---|---|
| *Benin* | 10.5M (6.5M-15.5M) | 4.0M (2.5M-6.0M) | 7.0K (4.4K-10.2K) | 5.6K (3.5K-8.1K) | 29.0M (10.5M-59.9M) | 11.1M (4.5M-21.7M) | 405.3M (132.7M-859.2M) |
| *Burkina Faso* | 21.4M (13.1M-32.0M) | 5.6M (3.3M-8.7M) | 11.3K (7.0K-16.5K) | 7.4K (4.6K-10.8K) | 22.8M (8.2M-47.2M) | 14.3M (5.7M-28.0M) | 140.5M (45.8M-299.2M) |
| *Côte d'Ivoire* | 25.3M (15.4M-37.9M) | 7.2M (4.3M-11.2M) | 14.7K (9.1K-21.6K) | 12.9K (8.0K-19.0K) | 101.8M (36.7M-211.3M) | 29.5M (11.7M-57.9M) | 1.4B (463.1M-3.0B) |
| *Ghana* | 28.6M (17.3M-43.1M) | 7.8M (4.6M-12.4M) | 16.2K (10.0K-23.8K) | 11.9K (7.4K-17.6K) | 129.8M (47.0M-269.2M) | 57.2M (22.8M-112.5M) | 1.6B (522.2M-3.4B) |
| *Guinea* | 10.1M (6.4M-14.6M) | 5.4M (3.5M-7.9M) | 7.8K (5.0K-11.1K) | 6.7K (4.3K-9.6K) | 18.8M (6.8M-38.3M) | 10.1M (4.2M-19.3M) | 277.9M (91.9M-582.3M) |
| *Gambia* | 1.7M (1.0M-2.6M) | 285.7K (168.4K-444.7K) | 864.7 (528.2-1.3K) | 694.1 (424.0-1.0K) | 2.4M (844.1K-4.9M) | 2.2M (851.7K-4.3M) | 10.6M (3.4M-22.8M) |
| *Guinea-Bissau* | 966.9K (581.7K-1.5M) | 592.1K (356.9K-896.0K) | 764.0 (467.4-1.1K) | 596.8 (365.1-886.2) | 1.6M (560.1K-3.3M) | 289.9K (114.0K-575.1K) | 7.0M (2.3M-15.1M) |
| *Liberia* | 3.4M (2.1M-5.1M) | 1.6M (959.6K-2.4M) | 2.4K (1.5K-3.5K) | 1.7K (1.1K-2.5K) | 2.9M (1.0M-5.9M) | 2.3M (925.1K-4.5M) | 13.7M (4.5M-29.2M) |
| *Mali* | 22.9M (14.1M-34.1M) | 5.3M (3.2M-8.3M) | 12.4K (7.7K-18.1K) | 10.4K (6.5K-15.2K) | 33.0M (11.9M-68.4M) | 6.2M (2.5M-12.2M) | 144.1M (47.0M-306.3M) |
| *Mauritania* | 3.2M (1.9M-4.8M) | 1.1M (668.0K-1.8M) | 2.0K (1.2K-2.9K) | 1.8K (1.1K-2.7K) | 9.4M (3.4M-19.6M) | 4.7M (1.8M-9.3M) | 192.4M (62.2M-412.3M) |
| *Niger* | 22.7M (13.9M-33.9M) | 8.5M (5.2M-12.9M) | 15.4K (9.5K-22.8K) | 7.5K (4.6K-11.1K) | 8.7M (3.1M-18.1M) | 12.5M (5.0M-24.7M) | 76.9M (25.0M-164.1M) |
| *Nigeria* | 165.5M (100.6M-248.4M) | 108.4M (66.4M-164.8M) | 125.4K (79.0K-182.0K) | 86.7K (54.6K-125.8K) | 646.7M (234.5M-1.3B) | 110.2M (44.5M-214.1M) | 10.5B (3.4B-22.2B) |
| *Senegal* | 10.5M (6.3M-15.9M) | 4.5M (2.7M-6.9M) | 7.2K (4.4K-10.6K) | 6.5K (4.0K-9.6K) | 24.9M (9.0M-52.2M) | 20.5M (8.1M-40.7M) | 415.3M (134.3M-889.9M) |
| *Sierra Leone* | 6.0M (3.8M-8.7M) | 2.9M (1.8M-4.2M) | 4.3K (2.8K-6.1K) | 3.2K (2.1K-4.5K) | 7.6M (2.8M-15.4M) | 1.8M (732.6K-3.3M) | 32.6M (10.8M-68.0M) |

| | | | | | | | |
|---|---|---|---|---|---|---|---|
| *Togo* | 5.9M (3.6M-8.7M) | 3.7M (2.3M-5.5M) | 4.7K (2.9K-6.9K) | 3.4K (2.1K-4.9K) | 11.6M (4.2M-24.0M) | 5.0M (2.0M-9.7M) | 54.6M (17.8M-116.2M) |
| **Total** | **338.9M (206.6M-506.3M)** | **166.9M (116.0M-289.3M)** | **232.3K (145.6K-338.7K)** | **167.0K (104.7K-243.6K)** | **1.1B (380.5M-2.2B)** | **287.7M (115.4M-562.9M)** | **15.3B (5.0B-32.4B)** |

***Table E.4. Cumulative economic burden of Lassa fever (per 100,000 population) by country in the absence of vaccination.*** *All figures represent means (95% uncertainty intervals) over ten years across 99 runs of the infection model and 100 runs of the health-economic model, for the baseline scenario assuming a probability of sequelae of 62% among patients discharged from hospital. Monetary costs are reported in International dollars (2021) and future costs are discounted at 3%/year. Catastrophic and impoverishing expenditures are reported as the number of individuals facing those costs. DALY = disability-adjusted life year, VSL = value of statistical life, K = thousand, M = million.*

| | Treatment costs, government-reimbursed ($ 2021) | Treatment costs, out-of-pocket ($ 2021) | Catastrophic expenditures (N) | Impoverishing expenditures (N) | Productivity losses ($ 2021) | Monetised DALYs ($ 2021) | VSL ($ 2021) |
|---|---|---|---|---|---|---|---|
| *Benin* | 83.9K (52.0K-123.6K) | 32.3K (19.9K-48.2K) | 56.0 (35.1-81.7) | 44.6 (27.9-65.0) | 232.1K (84.0K-478.8K) | 88.9K (35.7K-173.3K) | 3.2M (1.1M-6.9M) |
| *Burkina Faso* | 97.6K (59.6K-145.7K) | 25.4K (15.2K-39.8K) | 51.4 (32.0-75.4) | 33.7 (21.0-49.4) | 103.9K (37.5K-215.3K) | 65.0K (26.0K-127.6K) | 640.6K (208.9K-1.4M) |
| *Cote d'Ivoire* | 98.1K (59.6K-147.0K) | 27.8K (16.6K-43.4K) | 57.0 (35.3-83.7) | 50.2 (31.1-73.7) | 394.7K (142.3K-819.1K) | 114.2K (45.5K-224.5K) | 5.5M (1.8M-11.7M) |
| *Ghana* | 89.2K (53.9K-134.3K) | 24.4K (14.3K-38.8K) | 50.4 (31.2-74.1) | 37.2 (23.0-54.7) | 404.6K (146.4K-839.0K) | 178.2K (71.0K-350.7K) | 5.0M (1.6M-10.6M) |
| *Guinea* | 84.3K (53.7K-121.6K) | 45.2K (28.8K-65.6K) | 64.8 (41.7-92.6) | 55.8 (35.9-79.8) | 156.6K (57.0K-318.9K) | 84.2K (34.7K-161.2K) | 2.3M (766.2K-4.9M) |
| *Gambia* | 75.1K (45.5K-112.6K) | 12.4K (7.3K-19.3K) | 37.5 (22.9-55.7) | 30.1 (18.4-44.7) | 102.0K (36.6K-213.6K) | 94.0K (36.9K-186.6K) | 461.2K (149.3K-988.2K) |
| *Guinea-Bissau* | 53.4K (32.1K-80.4K) | 32.7K (19.7K-49.5K) | 42.2 (25.8-62.6) | 32.9 (20.2-48.9) | 86.2K (30.9K-180.4K) | 16.0K (6.3K-31.7K) | 388.2K (125.5K-832.3K) |
| *Liberia* | 78.5K (47.8K-117.3K) | 36.1K (21.9K-55.0K) | 54.3 (33.8-79.6) | 39.2 (24.4-57.5) | 65.1K (23.5K-134.9K) | 52.7K (21.1K-103.4K) | 312.7K (101.9K-665.8K) |
| *Mali* | 103.3K (63.4K-153.5K) | 24.0K (14.4K-37.3K) | 55.8 (34.8-81.5) | 46.8 (29.2-68.3) | 148.8K (53.6K-308.0K) | 28.1K (11.3K-55.0K) | 649.1K (211.9K-1.4M) |
| *Mauritania* | 73.4K (43.9K-111.0K) | 26.3K (15.5K-40.8K) | 45.6 (27.9-67.8) | 42.5 (26.0-63.2) | 217.7K (78.2K-455.5K) | 108.8K (42.8K-216.0K) | 4.5M (1.4M-9.6M) |
| *Niger* | 99.0K (60.4K-147.5K) | 37.1K (22.5K-56.0K) | 67.1 (41.3-99.2) | 32.8 (20.2-48.5) | 37.8K (13.6K-78.7K) | 54.6K (21.6K-107.5K) | 335.0K (108.8K-714.5K) |
| *Nigeria* | 79.0K (48.0K-118.6K) | 51.7K (31.7K-78.7K) | 59.8 (37.7-86.8) | 41.4 (26.1-60.0) | 308.6K (111.9K-635.4K) | 52.6K (21.2K-102.2K) | 5.0M (1.6M-10.6M) |
| *Senegal* | 66.7K (40.1K-100.5K) | 28.4K (17.0K-43.5K) | 45.3 (27.7-67.3) | 41.0 (25.1-60.9) | 157.8K (56.6K-330.0K) | 129.5K (50.9K-257.2K) | 2.6M (849.6K-5.6M) |
| *Sierra Leone* | 90.8K (58.0K-130.6K) | 43.4K (27.7K-63.1K) | 65.4 (42.5-92.7) | 48.2 (31.3-68.3) | 114.7K (41.9K-232.4K) | 26.5K (11.0K-50.3K) | 491.4K (163.3K-1.0M) |
| *Togo* | 70.4K (43.3K-104.4K) | 44.5K (27.4K-66.3K) | 56.4 (35.2-82.7) | 40.5 (25.2-59.3) | 139.1K (50.3K-288.0K) | 59.4K (23.8K-116.5K) | 655.2K (213.7K-1.4M) |
| **Total** | **84.2K (51.3K-125.8K)** | **41.5K (25.3K-63.1K)** | **57.7 (36.2-84.1)** | **41.5 (26.0-60.5)** | **261.0K (94.5K-538.5K)** | **71.5K (28.7K-139.8K)** | **3.8M (1.2M-8.0M)** |

***Table E.5. Sensitivity of economic outcomes to discounting and the probability of sequelae.*** *All figures represent means (95% uncertainty intervals) summed across all countries over ten years across 99 runs of the infection model and 100 runs of the health-economic model, varying the discounting rate and the probability of sequelae among patients discharged from hospital. Costs are reported in International dollars (2021). DALY = disability-adjusted life year, M = million, B = billion.*

| ***Discounting rate*** | ***Probability of sequelae*** | **Monetised DALYs ($ 2021)** | **Societal costs ($ 2021)** |
|---|---|---|---|
| *0%* | *17%* | 556.6M (200.9M-1.1B) | 2.5B (1.2B-4.7B) |
| *0%* | *62%* | 613.8M (245.3M-1.2B) | 2.5B (1.2B-4.7B) |
| *3%* | *17%* | 261.0M (94.5M-537.4M) | 1.6B (805.1M-2.8B) |
| *3%* | *62%* | 287.7M (115.4M-562.9M) | 1.6B (805.1M-2.8B) |

##### *E.2. Lassa fever vaccine impact*

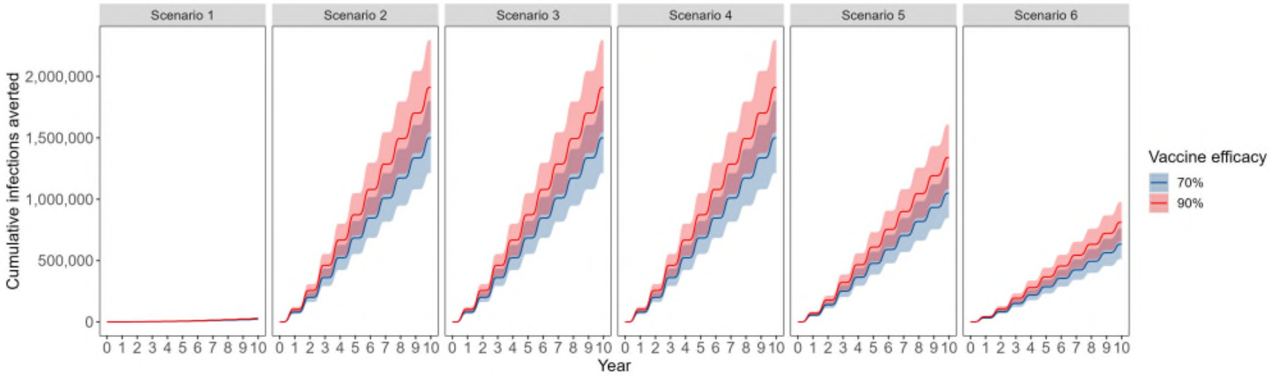


***Figure E.4. Vaccine impact in endemic districts.*** *For each vaccination scenario described in* ***Table C.2****, the mean cumulative number of infections averted due to vaccination (lines), summed across the 16 districts classified as endemic. Shading around each line indicates 95% uncertainty intervals. Vaccine efficacy refers to efficacy against infection (*$VE_{infect}$*).*

***Table E.6. Ten-year health burden of Lassa fever averted due to vaccine with 70% efficacy.*** *Columns represent the vaccination scenarios considered and rows represent the outcomes averted over ten years from the initiation of vaccine rollout. All figures represent means (95% uncertainty intervals) across 99 runs of the infection model and 100 runs of the health-economic model simulations, for the baseline scenario assuming a probability of sequelae of 62% among patients discharged from hospital. LASV = Lassa virus, DALY = disability-adjusted life-year, VE = vaccine efficacy, K = thousand, M = million.*

| ***Outcome averted due to vaccination*** | **Scenario 1** Outbreak response only | **Scenario 2** Endemic districts (80%) | **Scenario 3** Endemic districts (80%) + non-endemic districts of high-endemic countries (5%) | **Scenario 4** Endemic districts (80%) + non-endemic districts of all countries (5%) | **Scenario 5** Endemic districts (55%) + non-endemic districts of high-endemic countries (5%) | **Scenario 6** Endemic districts (32.5%) + non-endemic districts of all countries (5%) |
|---|---|---|---|---|---|---|
| **Vaccine 70% effective against disease** ($VE_{infect} = 0\%, VE_{disease} = 70\%$) | | | | | | |
| *LASV infections* | 0.0 (0.0-0.0) | 0.0 (0.0-0.0) | 0.0 (0.0-0.0) | 0.0 (0.0-0.0) | 0.0 (0.0-0.0) | 0.0 (0.0-0.0) |
| *Mild/moderate cases* | 37.6K (18.5K-68.3K) | 322.8K (160.7K-580.8K) | 396.8K (197.2K-715.4K) | 456.0K (226.4K-822.7K) | 307.7K (152.9K-554.9K) | 286.7K (141.7K-519.1K) |
| *Hospitalisations* | 1.6K (1.0K-2.4K) | 14.1K (9.0K-20.3K) | 17.3K (11.1K-25.0K) | 19.9K (12.7K-28.8K) | 13.4K (8.6K-19.4K) | 12.5K (7.9K-18.1K) |
| *Deaths* | 272.0 (89.0-576.8) | 2.3K (771.7-4.9K) | 2.9K (947.3-6.0K) | 3.3K (1.1K-7.0K) | 2.2K (734.0-4.7K) | 2.1K (680.7-4.4K) |
| *Sequelae* | 847.5 (518.2-1.2K) | 7.3K (4.6K-10.4K) | 9.0K (5.6K-12.8K) | 10.3K (6.4K-14.8K) | 6.9K (4.3K-9.9K) | 6.5K (4.0K-9.3K) |
| *DALYs* | 13.7K (5.5K-26.8K) | 115.4K (47.1K-222.2K) | 141.4K (57.6K-273.2K) | 164.1K (66.7K-317.7K) | 109.6K (44.6K-211.9K) | 103.8K (41.9K-201.4K) |
| **Vaccine 70% effective against infection and disease** ($VE_{infect} = 70\%, VE_{disease} = 70\%$) | | | | | | |
| *LASV infections* | 200.2K (153.5K-250.0K) | 1.7M (1.3M-2.0M) | 2.1M (1.6M-2.5M) | 2.4M (1.9M-2.9M) | 1.6M (1.3M-2.0M) | 1.5M (1.2M-1.9M) |
| *Mild/moderate cases* | 51.2K (25.2K-93.1K) | 431.2K (214.7K-776.0K) | 532.2K (264.4K-959.4K) | 612.7K (304.1K-1.1M) | 415.4K (206.4K-749.1K) | 389.3K (192.4K-704.9K) |
| *Hospitalisations* | 2.2K (1.4K-3.3K) | 18.8K (12.0K-27.1K) | 23.2K (14.8K-33.5K) | 26.8K (17.0K-38.6K) | 18.1K (11.6K-26.2K) | 17.0K (10.8K-24.6K) |
| *Deaths* | 370.6 (121.3-785.9) | 3.1K (1.0K-6.6K) | 3.9K (1.3K-8.1K) | 4.4K (1.5K-9.4K) | 3.0K (990.8-6.3K) | 2.8K (924.5-6.0K) |
| *Sequelae* | 1.2K (706.1-1.7K) | 9.7K (6.1K-13.9K) | 12.0K (7.5K-17.2K) | 13.8K (8.6K-19.8K) | 9.4K (5.8K-13.4K) | 8.8K (5.4K-12.7K) |
| *DALYs* | 18.7K (7.5K-36.5K) | 154.2K (63.0K-296.8K) | 189.6K (77.3K-366.3K) | 220.5K (89.6K-427.0K) | 148.0K (60.2K-286.1K) | 140.9K (56.9K-273.6K) |

***Table E.7. Ten-year health burden of Lassa fever averted due to vaccine with 90% efficacy.*** *Columns represent the vaccination scenarios considered and rows represent the outcomes averted over ten years from the initiation of vaccine rollout. All figures represent means (95% uncertainty intervals) across 99 runs of the infection model and 100 runs of the health-economic model simulations, for the baseline scenario assuming a probability of sequelae of 62% among patients discharged from hospital. LASV = Lassa virus, DALY = disability-adjusted life-year, VE = vaccine efficacy, K = thousand, M = million.*

| ***Outcome averted due to vaccination*** | **Scenario 1** Outbreak response only | **Scenario 2** Endemic districts (80%) | **Scenario 3** Endemic districts (80%) + non-endemic districts of high-endemic countries (5%) | **Scenario 4** Endemic districts (80%) + non-endemic districts of all countries (5%) | **Scenario 5** Endemic districts (55%) + non-endemic districts of high-endemic countries (5%) | **Scenario 6** Endemic districts (32.5%) + non-endemic districts of all countries (5%) |
|---|---|---|---|---|---|---|
| **Vaccine 90% effective against disease** ($VE_{infect} = 0\%, VE_{disease} = 90\%$) | | | | | | |
| *LASV infections* | 0.0 (0.0-0.0) | 0.0 (0.0-0.0) | 0.0 (0.0-0.0) | 0.0 (0.0-0.0) | 0.0 (0.0-0.0) | 0.0 (0.0-0.0) |
| *Mild/moderate cases* | 48.3K (23.8K-87.8K) | 415.0K (206.6K-746.8K) | 510.2K (253.5K-919.8K) | 586.3K (291.1K-1.1M) | 395.6K (196.6K-713.4K) | 368.6K (182.2K-667.4K) |
| *Hospitalisations* | 2.1K (1.3K-3.1K) | 18.1K (11.6K-26.1K) | 22.3K (14.2K-32.1K) | 25.6K (16.3K-37.0K) | 17.3K (11.0K-24.9K) | 16.1K (10.2K-23.3K) |
| *Deaths* | 349.7 (114.5-741.6) | 3.0K (992.2-6.3K) | 3.7K (1.2K-7.8K) | 4.2K (1.4K-8.9K) | 2.9K (943.7-6.0K) | 2.7K (875.3-5.6K) |
| *Sequelae* | 1.1K (666.3-1.6K) | 9.4K (5.9K-13.4K) | 11.5K (7.2K-16.5K) | 13.2K (8.2K-19.0K) | 8.9K (5.6K-12.8K) | 8.3K (5.1K-12.0K) |
| *DALYs* | 17.6K (7.1K-34.5K) | 148.3K (60.6K-285.6K) | 181.8K (74.1K-351.2K) | 211.0K (85.7K-408.5K) | 140.9K (57.4K-272.4K) | 133.4K (53.9K-259.0K) |
| **Vaccine 90% effective against infection and disease** ($VE_{infect} = 90\%, VE_{disease} = 90\%$) | | | | | | |
| *LASV infections* | 257.4K (197.4K-321.4K) | 2.1M (1.7M-2.6M) | 2.6M (2.1M-3.2M) | 3.1M (2.4M-3.7M) | 2.1M (1.6M-2.5M) | 2.0M (1.5M-2.4M) |
| *Mild/moderate cases* | 56.2K (27.6K-102.1K) | 468.0K (233.0K-842.1K) | 578.6K (287.5K-1.0M) | 666.9K (331.0K-1.2M) | 453.2K (225.2K-817.3K) | 426.1K (210.6K-771.6K) |
| *Hospitalisations* | 2.5K (1.5K-3.6K) | 20.4K (13.1K-29.4K) | 25.3K (16.1K-36.4K) | 29.1K (18.5K-42.1K) | 19.8K (12.6K-28.5K) | 18.6K (11.8K-27.0K) |
| *Deaths* | 406.6 (133.0-862.1) | 3.4K (1.1K-7.1K) | 4.2K (1.4K-8.8K) | 4.8K (1.6K-10.2K) | 3.3K (1.1K-6.9K) | 3.1K (1.0K-6.5K) |
| *Sequelae* | 1.3K (774.5-1.8K) | 10.6K (6.6K-15.1K) | 13.1K (8.1K-18.7K) | 15.0K (9.3K-21.6K) | 10.2K (6.4K-14.7K) | 9.6K (5.9K-13.9K) |
| *DALYs* | 20.5K (8.2K-40.1K) | 167.3K (68.3K-322.1K) | 206.1K (84.0K-398.3K) | 240.1K (97.5K-464.9K) | 161.4K (65.7K-312.1K) | 154.2K (62.3K-299.5K) |

***Table E.8. Ten-year economic burden of Lassa fever averted due to vaccine with 70% efficacy.*** *Columns represent the vaccination scenarios considered and rows represent the outcomes averted over ten years from the initiation of vaccine rollout. All figures represent means (95% uncertainty intervals) across 99 runs of the infection model and 100 runs of the health-economic model simulations, for the baseline scenario assuming a probability of sequelae of 62% among patients discharged from hospital. Monetary costs are reported in International dollars (2021) and future costs are discounted at 3%/year. Catastrophic and impoverishing expenditures are reported as the number of individuals facing those costs. DALY = disability-adjusted life-year, VSL = value of statistical life, VE = vaccine efficacy, K = thousand, M = million, B = billion.*

| ***Outcome averted due to vaccination*** | **Scenario 1** Outbreak response only | **Scenario 2** Endemic districts (80%) | **Scenario 3** Endemic districts (80%) + non-endemic districts of high-endemic countries (5%) | **Scenario 4** Endemic districts (80%) + non-endemic districts of all countries (5%) | **Scenario 5** Endemic districts (55%) + non-endemic districts of high-endemic countries (5%) | **Scenario 6** Endemic districts (32.5%) + non-endemic districts of all countries (5%) |
|---|---|---|---|---|---|---|
| **Vaccine 70% effective against disease** ($VE_{infect} = 0\%, VE_{disease} = 70\%$) | | | | | | |
| *Treatment costs, government-reimbursed ($ 2021)* | 2.2M (1.4M-3.4M) | 18.6M (11.5M-27.5M) | 22.8M (14.1M-33.8M) | 26.8M (16.5M-39.8M) | 17.7M (10.9M-26.3M) | 17.1M (10.5M-25.5M) |
| *Treatment costs, out-of-pocket ($ 2021)* | 1.1M (674.4K-1.7M) | 11.2M (7.0M-16.7M) | 13.9M (8.6M-20.8M) | 15.1M (9.4M-22.8M) | 10.7M (6.7M-16.1M) | 9.2M (5.7M-13.9M) |
| *Catastrophic expenditures (N)* | 1.6K (1.0K-2.3K) | 14.1K (9.0K-20.2K) | 17.3K (11.0K-24.9K) | 19.8K (12.6K-28.5K) | 13.4K (8.5K-19.3K) | 12.4K (7.8K-17.9K) |
| *Impoverishing expenditures (N)* | 1.2K (725.0-1.7K) | 10.1K (6.5K-14.5K) | 12.4K (7.9K-17.8K) | 14.2K (9.0K-20.5K) | 9.6K (6.1K-13.8K) | 8.9K (5.6K-12.9K) |
| *Productivity losses ($ 2021)* | 7.0M (2.5M-14.4M) | 60.5M (22.0M-123.7M) | 76.2M (27.7M-155.9M) | 86.3M (31.3M-176.9M) | 59.4M (21.6M-121.7M) | 54.5M (19.8M-112.1M) |
| *Monetised DALYs ($ 2021)* | 1.9M (763.1K-3.7M) | 12.9M (5.3M-24.9M) | 15.8M (6.4M-30.4M) | 20.1M (8.2M-39.0M) | 12.3M (5.0M-23.8M) | 13.6M (5.5M-26.4M) |
| *VSL ($ 2021)* | 105.7M (34.6M-224.1M) | 948.9M (313.2M-2.0B) | 1.2B (396.6M-2.5B) | 1.3B (436.8M-2.8B) | 939.7M (309.4M-2.0B) | 826.3M (271.1M-1.7B) |
| **Vaccine 70% effective against infection and disease** ($VE_{infect} = 70\%, VE_{disease} = 70\%$) | | | | | | |
| *Treatment costs, government-reimbursed ($ 2021)* | 3.1M (1.9M-4.6M) | 24.8M (15.4M-36.7M) | 30.6M (18.9M-45.4M) | 36.0M (22.2M-53.5M) | 23.9M (14.7M-35.5M) | 23.2M (14.3M-34.6M) |
| *Treatment costs, out-of-pocket ($ 2021)* | 1.5M (919.0K-2.3M) | 14.9M (9.3M-22.3M) | 18.6M (11.6M-28.0M) | 20.3M (12.6M-30.6M) | 14.5M (9.0M-21.8M) | 12.5M (7.7M-18.8M) |
| *Catastrophic expenditures (N)* | 2.2K (1.4K-3.2K) | 18.8K (12.0K-27.0K) | 23.2K (14.8K-33.4K) | 26.5K (16.9K-38.3K) | 18.1K (11.5K-26.1K) | 16.8K (10.6K-24.3K) |
| *Impoverishing expenditures (N)* | 1.6K (987.8-2.3K) | 13.5K (8.6K-19.4K) | 16.6K (10.6K-23.9K) | 19.1K (12.1K-27.6K) | 12.9K (8.2K-18.6K) | 12.1K (7.6K-17.5K) |
| *Productivity losses ($ 2021)* | 9.5M (3.4M-19.6M) | 80.9M (29.4M-165.3M) | 102.2M (37.2M-209.2M) | 115.9M (42.1M-237.8M) | 80.3M (29.2M-164.4M) | 74.0M (26.8M-152.3M) |
| *Monetised DALYs ($ 2021)* | 2.6M (1.0M-5.1M) | 17.3M (7.1M-33.3M) | 21.1M (8.6M-40.8M) | 27.1M (11.0M-52.5M) | 16.6M (6.8M-32.2M) | 18.4M (7.4M-35.9M) |
| *VSL ($ 2021)* | 144.1M (47.2M-305.4M) | 1.3B (418.5M-2.7B) | 1.6B (532.1M-3.4B) | 1.8B (586.7M-3.8B) | 1.3B (417.8M-2.7B) | 1.1B (368.1M-2.4B) |

***Table E.9. Ten-year economic burden of Lassa fever averted due to vaccine with 90% efficacy.*** *Columns represent the vaccination scenarios considered and rows represent the outcomes averted over ten years from the initiation of vaccine rollout. All figures represent means (95% uncertainty intervals) across 99 runs of the infection model and 100 runs of the health-economic model simulations, for the baseline scenario assuming a probability of sequelae of 62% among patients discharged from hospital. Monetary costs are reported in International dollars (2021) and future costs are discounted at 3%/year. Catastrophic and impoverishing expenditures are reported as the number of individuals facing those costs. DALY = disability-adjusted life-year, VSL = value of statistical life, VE = vaccine efficacy, K = thousand, M = million, B = billion.*

| ***Outcome averted due to vaccination*** | **Scenario 1** Outbreak response only | **Scenario 2** Endemic districts (80%) | **Scenario 3** Endemic districts (80%) + non-endemic districts of high-endemic countries (5%) | **Scenario 4** Endemic districts (80%) + non-endemic districts of all countries (5%) | **Scenario 5** Endemic districts (55%) + non-endemic districts of high-endemic countries (5%) | **Scenario 6** Endemic districts (32.5%) + non-endemic districts of all countries (5%) |
|---|---|---|---|---|---|---|
| **Vaccine 90% effective against disease** ($VE_{infect} = 0\%, VE_{disease} = 90\%$) | | | | | | |
| *Treatment costs, government-reimbursed ($ 2021)* | 2.9M (1.8M-4.3M) | 23.9M (14.8M-35.4M) | 29.3M (18.1M-43.5M) | 34.5M (21.3M-51.1M) | 22.8M (14.0M-33.8M) | 22.0M (13.5M-32.8M) |
| *Treatment costs, out-of-pocket ($ 2021)* | 1.4M (867.1K-2.2M) | 14.4M (8.9M-21.5M) | 17.8M (11.1M-26.8M) | 19.5M (12.1M-29.3M) | 13.8M (8.6M-20.8M) | 11.8M (7.3M-17.8M) |
| *Catastrophic expenditures (N)* | 2.1K (1.3K-3.0K) | 18.1K (11.6K-26.0K) | 22.2K (14.2K-32.0K) | 25.4K (16.2K-36.7K) | 17.2K (11.0K-24.8K) | 15.9K (10.1K-23.0K) |
| *Impoverishing expenditures (N)* | 1.5K (932.1-2.2K) | 13.0K (8.3K-18.7K) | 15.9K (10.1K-22.9K) | 18.3K (11.6K-26.4K) | 12.3K (7.8K-17.7K) | 11.4K (7.2K-16.6K) |
| *Productivity losses ($ 2021)* | 8.9M (3.2M-18.5M) | 77.8M (28.3M-159.0M) | 97.9M (35.6M-200.4M) | 110.9M (40.3M-227.4M) | 76.4M (27.8M-156.5M) | 70.0M (25.4M-144.1M) |
| *Monetised DALYs ($ 2021)* | 2.4M (981.1K-4.8M) | 16.6M (6.8M-32.0M) | 20.3M (8.3M-39.1M) | 25.8M (10.5M-50.1M) | 15.8M (6.4M-30.6M) | 17.4M (7.0M-33.9M) |
| *VSL ($ 2021)* | 136.0M (44.5M-288.2M) | 1.2B (402.7M-2.6B) | 1.5B (509.9M-3.3B) | 1.7B (561.5M-3.6B) | 1.2B (397.8M-2.5B) | 1.1B (348.5M-2.2B) |
| **Vaccine 90% effective against infection and disease** ($VE_{infect} = 90\%, VE_{disease} = 90\%$) | | | | | | |
| *Treatment costs, government-reimbursed ($ 2021)* | 3.4M (2.0M-5.0M) | 27.0M (16.7M-39.9M) | 33.3M (20.5M-49.3M) | 39.2M (24.2M-58.2M) | 26.1M (16.1M-38.7M) | 25.4M (15.6M-37.9M) |
| *Treatment costs, out-of-pocket ($ 2021)* | 1.7M (1.0M-2.5M) | 16.2M (10.1M-24.2M) | 20.2M (12.6M-30.4M) | 22.1M (13.7M-33.3M) | 15.8M (9.8M-23.8M) | 13.6M (8.4M-20.6M) |
| *Catastrophic expenditures (N)* | 2.4K (1.5K-3.5K) | 20.4K (13.0K-29.3K) | 25.2K (16.1K-36.3K) | 28.9K (18.4K-41.7K) | 19.7K (12.6K-28.4K) | 18.4K (11.6K-26.6K) |
| *Impoverishing expenditures (N)* | 1.7K (1.1K-2.5K) | 14.6K (9.4K-21.0K) | 18.0K (11.5K-26.0K) | 20.8K (13.2K-30.0K) | 14.1K (9.0K-20.3K) | 13.2K (8.4K-19.1K) |
| *Productivity losses ($ 2021)* | 10.4M (3.8M-21.4M) | 87.8M (31.9M-179.4M) | 111.1M (40.4M-227.6M) | 126.2M (45.8M-258.9M) | 87.6M (31.8M-179.5M) | 81.0M (29.4M-166.7M) |
| *Monetised DALYs ($ 2021)* | 2.8M (1.1M-5.6M) | 18.8M (7.7M-36.1M) | 23.0M (9.4M-44.4M) | 29.5M (12.0M-57.2M) | 18.1M (7.4M-35.1M) | 20.2M (8.1M-39.3M) |
| *VSL ($ 2021)* | 158.0M (51.8M-335.0M) | 1.4B (454.1M-2.9B) | 1.8B (578.7M-3.7B) | 1.9B (638.5M-4.1B) | 1.4B (456.0M-2.9B) | 1.2B (402.9M-2.6B) |

***Table E.10. Ten-year Lassa fever burden and economic costs averted per 100,000 population due to vaccine with 70% efficacy.*** *Columns represent the vaccination scenarios considered and rows represent the outcomes averted over ten years from the initiation of vaccine rollout. All figures represent means (95% uncertainty intervals) across 99 runs of the infection model and 100 runs of the health-economic model simulations, for the baseline scenario assuming a probability of sequelae of 62% among patients discharged from hospital. Monetary costs are reported in International dollars (2021) and future monetary costs are discounted at 3%/year. LASV = Lassa virus, DALY = disability-adjusted life-year, VE = vaccine efficacy, K = thousand, M = million.*

| ***Outcome averted due to vaccination*** | **Scenario 1** Outbreak response only | **Scenario 2** Endemic districts (80%) | **Scenario 3** Endemic districts (80%) + non-endemic districts of high-endemic countries (5%) | **Scenario 4** Endemic districts (80%) + non-endemic districts of all countries (5%) | **Scenario 5** Endemic districts (55%) + non-endemic districts of high-endemic countries (5%) | **Scenario 6** Endemic districts (32.5%) + non-endemic districts of all countries (5%) |
|---|---|---|---|---|---|---|
| **Vaccine 70% effective against disease** ($VE_{infect} = 0\%, VE_{disease} = 70\%$) | | | | | | |
| *LASV infections (N)* | 0.0 (0.0-0.0) | 0.0 (0.0-0.0) | 0.0 (0.0-0.0) | 0.0 (0.0-0.0) | 0.0 (0.0-0.0) | 0.0 (0.0-0.0) |
| *Hospitalisations (N)* | 0.4 (0.3-0.6) | 3.5 (2.2-5.0) | 4.3 (2.7-6.2) | 4.9 (3.1-7.1) | 3.3 (2.1-4.8) | 3.1 (2.0-4.5) |
| *Deaths (N)* | 0.1 (0.0-0.1) | 0.6 (0.2-1.2) | 0.7 (0.2-1.5) | 0.8 (0.3-1.7) | 0.6 (0.2-1.2) | 0.5 (0.2-1.1) |
| *DALYs (N)* | 3.4 (1.4-6.7) | 28.7 (11.7-55.2) | 35.1 (14.3-67.8) | 40.8 (16.6-78.9) | 27.2 (11.1-52.6) | 25.8 (10.4-50.0) |
| *Impoverishing expenditures (N)* | 0.3 (0.2-0.4) | 2.5 (1.6-3.6) | 3.1 (2.0-4.4) | 3.5 (2.2-5.1) | 2.4 (1.5-3.4) | 2.2 (1.4-3.2) |
| *Societal costs (I$ 2021)* | 2.6K (1.3K-4.7K) | 22.4K (11.8K-40.4K) | 28.0K (14.7K-50.6K) | 31.8K (16.7K-57.6K) | 21.8K (11.4K-39.5K) | 20.1K (10.4K-36.4K) |
| *Monetised DALYs (I$ 2021)* | 472.8 (189.5-925.0) | 3.2K (1.3K-6.2K) | 3.9K (1.6K-7.6K) | 5.0K (2.0K-9.7K) | 3.1K (1.2K-5.9K) | 3.4K (1.4K-6.6K) |
| *VSL (I$ 2021)* | 26.3K (8.6K-55.7K) | 235.7K (77.8K-495.1K) | 298.8K (98.5K-629.3K) | 329.6K (108.5K-694.8K) | 233.4K (76.8K-492.0K) | 205.2K (67.3K-433.5K) |
| **Vaccine 70% effective against infection and disease** ($VE_{infect} = 70\%, VE_{disease} = 70\%$) | | | | | | |
| *LASV infections (N)* | 49.7 (38.1-62.1) | 416.5 (332.5-505.4) | 514.6 (408.2-627.2) | 592.8 (467.0-726.2) | 402.4 (318.2-491.7) | 377.8 (294.4-466.5) |
| *Hospitalisations (N)* | 0.6 (0.3-0.8) | 4.7 (3.0-6.7) | 5.8 (3.7-8.3) | 6.6 (4.2-9.6) | 4.5 (2.9-6.5) | 4.2 (2.7-6.1) |
| *Deaths (N)* | 0.1 (0.0-0.2) | 0.8 (0.3-1.6) | 1.0 (0.3-2.0) | 1.1 (0.4-2.3) | 0.7 (0.2-1.6) | 0.7 (0.2-1.5) |
| *DALYs (N)* | 4.6 (1.9-9.1) | 38.3 (15.6-73.7) | 47.1 (19.2-91.0) | 54.8 (22.3-106.1) | 36.7 (15.0-71.0) | 35.0 (14.1-68.0) |
| *Impoverishing expenditures (N)* | 0.4 (0.2-0.6) | 3.4 (2.1-4.8) | 4.1 (2.6-5.9) | 4.7 (3.0-6.8) | 3.2 (2.0-4.6) | 3.0 (1.9-4.3) |
| *Societal costs (I$ 2021)* | 3.5K (1.8K-6.4K) | 30.0K (15.8K-53.9K) | 37.6K (19.7K-67.9K) | 42.8K (22.4K-77.4K) | 29.5K (15.4K-53.3K) | 27.2K (14.2K-49.5K) |
| *Monetised DALYs (I$ 2021)* | 644.2 (258.2-1.3K) | 4.3K (1.8K-8.3K) | 5.3K (2.1K-10.1K) | 6.7K (2.7K-13.0K) | 4.1K (1.7K-8.0K) | 4.6K (1.8K-8.9K) |
| *VSL (I$ 2021)* | 35.8K (11.7K-75.9K) | 314.9K (103.9K-661.6K) | 401.0K (132.1K-844.6K) | 442.8K (145.7K-933.5K) | 315.2K (103.8K-664.5K) | 278.8K (91.4K-588.9K) |

***Table E.11. Ten-year Lassa fever burden and economic costs averted per 100,000 population due to vaccine with 90% efficacy.*** *Columns represent the vaccination scenarios considered and rows represent the outcomes averted over ten years from the initiation of vaccine rollout. All figures represent means (95% uncertainty intervals) across 99 runs of the infection model and 100 runs of the health-economic model simulations, for the baseline scenario assuming a probability of sequelae of 62% among patients discharged from hospital. Monetary costs are reported in International dollars (2021) and future costs are discounted at 3%/year. LASV = Lassa virus, DALY = disability-adjusted life-year, VE = vaccine efficacy, K = thousand, M = million.*

| ***Outcome averted due to vaccination*** | **Scenario 1** Outbreak response only | **Scenario 2** Endemic districts (80%) | **Scenario 3** Endemic districts (80%) + non-endemic districts of high-endemic countries (5%) | **Scenario 4** Endemic districts (80%) + non-endemic districts of all countries (5%) | **Scenario 5** Endemic districts (55%) + non-endemic districts of high-endemic countries (5%) | **Scenario 6** Endemic districts (32.5%) + non-endemic districts of all countries (5%) |
|---|---|---|---|---|---|---|
| **Vaccine 90% effective against disease** ($VE_{infect} = 0\%, VE_{disease} = 90\%$) | | | | | | |
| *LASV infections (N)* | 0.0 (0.0-0.0) | 0.0 (0.0-0.0) | 0.0 (0.0-0.0) | 0.0 (0.0-0.0) | 0.0 (0.0-0.0) | 0.0 (0.0-0.0) |
| *Hospitalisations (N)* | 0.5 (0.3-0.8) | 4.5 (2.9-6.5) | 5.5 (3.5-8.0) | 6.4 (4.0-9.2) | 4.3 (2.7-6.2) | 4.0 (2.5-5.8) |
| *Deaths (N)* | 0.1 (0.0-0.2) | 0.7 (0.2-1.6) | 0.9 (0.3-1.9) | 1.1 (0.3-2.2) | 0.7 (0.2-1.5) | 0.7 (0.2-1.4) |
| *DALYs (N)* | 4.4 (1.8-8.6) | 36.8 (15.0-70.9) | 45.1 (18.4-87.2) | 52.4 (21.3-101.5) | 35.0 (14.2-67.7) | 33.1 (13.4-64.3) |
| *Impoverishing expenditures (N)* | 0.4 (0.2-0.5) | 3.2 (2.1-4.6) | 3.9 (2.5-5.7) | 4.5 (2.9-6.5) | 3.1 (1.9-4.4) | 2.8 (1.8-4.1) |
| *Societal costs (I$ 2021)* | 3.3K (1.7K-6.0K) | 28.8K (15.2K-51.9K) | 36.0K (18.9K-65.1K) | 40.9K (21.5K-74.1K) | 28.1K (14.7K-50.7K) | 25.8K (13.4K-46.8K) |
| *Monetised DALYs (I$ 2021)* | 607.9 (243.7-1.2K) | 4.1K (1.7K-8.0K) | 5.0K (2.1K-9.7K) | 6.4K (2.6K-12.4K) | 3.9K (1.6K-7.6K) | 4.3K (1.7K-8.4K) |
| *VSL (I$ 2021)* | 33.8K (11.1K-71.6K) | 303.0K (100.0K-636.6K) | 384.2K (126.6K-809.2K) | 423.7K (139.5K-893.3K) | 300.1K (98.8K-632.5K) | 263.9K (86.6K-557.4K) |
| **Vaccine 90% effective against infection and disease** ($VE_{infect} = 90\%, VE_{disease} = 90\%$) | | | | | | |
| *LASV infections (N)* | 63.9 (49.0-79.8) | 531.2 (424.1-644.5) | 657.2 (521.4-801.0) | 757.7 (596.9-928.4) | 515.3 (407.5-629.5) | 484.9 (377.9-598.7) |
| *Hospitalisations (N)* | 0.6 (0.4-0.9) | 5.1 (3.2-7.3) | 6.3 (4.0-9.0) | 7.2 (4.6-10.4) | 4.9 (3.1-7.1) | 4.6 (2.9-6.7) |
| *Deaths (N)* | 0.1 (0.0-0.2) | 0.8 (0.3-1.8) | 1.0 (0.3-2.2) | 1.2 (0.4-2.5) | 0.8 (0.3-1.7) | 0.8 (0.3-1.6) |
| *DALYs (N)* | 5.1 (2.0-9.9) | 41.5 (17.0-80.0) | 51.2 (20.9-98.9) | 59.6 (24.2-115.5) | 40.1 (16.3-77.5) | 38.3 (15.5-74.4) |
| *Impoverishing expenditures (N)* | 0.4 (0.3-0.6) | 3.6 (2.3-5.2) | 4.5 (2.9-6.4) | 5.2 (3.3-7.4) | 3.5 (2.2-5.0) | 3.3 (2.1-4.8) |
| *Societal costs (I$ 2021)* | 3.8K (2.0K-7.0K) | 32.5K (17.2K-58.5K) | 40.9K (21.4K-73.9K) | 46.6K (24.4K-84.3K) | 32.2K (16.8K-58.2K) | 29.8K (15.5K-54.2K) |
| *Monetised DALYs (I$ 2021)* | 706.6 (283.3-1.4K) | 4.7K (1.9K-9.0K) | 5.7K (2.3K-11.0K) | 7.3K (3.0K-14.2K) | 4.5K (1.8K-8.7K) | 5.0K (2.0K-9.7K) |
| *VSL (I$ 2021)* | 39.3K (12.9K-83.2K) | 341.7K (112.8K-718.0K) | 436.1K (143.7K-918.7K) | 482.0K (158.6K-1.0M) | 344.0K (113.3K-725.1K) | 305.1K (100.1K-644.5K) |

***Table E.12. Threshold vaccine cost (TVC) across Lassa vaccination scenarios, reported in International dollars ($ 2021).*** *Columns represent the vaccination scenarios considered and rows represent the economic costs considered, conditional on vaccine efficacy. All figures represent means (95% uncertainty intervals) across 99 runs of the infection model and 100 runs of the health-economic model simulations, for the baseline scenario assuming a probability of sequelae of 62% among patients discharged from hospital. Healthcare costs combine outpatient treatment costs and hospital treatment costs. Societal costs combine healthcare costs and productivity losses. Future monetary costs are discounted at 3%/year.*

| ***Economic costs considered*** | ***Vaccine efficacy*** | **Scenario 1** Outbreak response only | **Scenario 2** Endemic districts (80%) | **Scenario 3** Endemic districts (80%) + non-endemic districts of high-endemic countries (5%) | **Scenario 4** Endemic districts (80%) + non-endemic districts of all countries (5%) | **Scenario 5** Endemic districts (55%) + non-endemic districts of high-endemic countries (5%) | **Scenario 6** Endemic districts (32.5%) + non-endemic districts of all countries (5%) |
|---|---|---|---|---|---|---|---|
| **Healthcare costs + monetised DALYs** | 0% infection / 70% disease | 0.51 (0.30-0.80) | 0.63 (0.39-0.96) | 0.61 (0.38-0.95) | 0.62 (0.38-0.96) | 0.60 (0.37-0.93) | 0.60 (0.36-0.94) |
| | 0% infection / 90% disease | 0.65 (0.39-1.03) | 0.81 (0.50-1.24) | 0.79 (0.48-1.22) | 0.79 (0.48-1.23) | 0.78 (0.48-1.20) | 0.77 (0.47-1.21) |
| | 70% infection / 70% disease | 0.69 (0.41-1.10) | 0.84 (0.52-1.29) | 0.82 (0.50-1.27) | 0.83 (0.51-1.29) | 0.81 (0.50-1.26) | 0.81 (0.49-1.28) |
| | 90% infection / 90% disease | 0.76 (0.45-1.20) | 0.91 (0.56-1.40) | 0.89 (0.55-1.38) | 0.90 (0.55-1.40) | 0.89 (0.55-1.37) | 0.89 (0.54-1.40) |
| **Societal costs + monetised DALYs** | 0% infection / 70% disease | 1.18 (0.59-2.17) | 1.52 (0.78-2.75) | 1.50 (0.77-2.73) | 1.47 (0.75-2.68) | 1.48 (0.76-2.70) | 1.42 (0.72-2.59) |
| | 0% infection / 90% disease | 1.52 (0.76-2.79) | 1.95 (1.01-3.53) | 1.93 (0.99-3.51) | 1.89 (0.97-3.45) | 1.91 (0.98-3.47) | 1.83 (0.93-3.34) |
| | 70% infection / 70% disease | 1.61 (0.81-2.95) | 2.03 (1.05-3.67) | 2.02 (1.04-3.66) | 1.98 (1.01-3.60) | 2.00 (1.03-3.65) | 1.93 (0.98-3.52) |
| | 90% infection / 90% disease | 1.76 (0.89-3.24) | 2.20 (1.13-3.98) | 2.19 (1.13-3.99) | 2.16 (1.10-3.92) | 2.19 (1.12-3.98) | 2.11 (1.07-3.86) |
| **Value of statistical life + healthcare costs** | 0% infection / 70% disease | 10.54 (3.61-22.06) | 14.38 (4.96-29.81) | 14.49 (4.99-30.13) | 13.59 (4.68-28.30) | 14.34 (4.93-29.85) | 12.84 (4.41-26.78) |
| | 0% infection / 90% disease | 13.55 (4.65-28.36) | 18.48 (6.37-38.33) | 18.63 (6.41-38.73) | 17.48 (6.02-36.38) | 18.44 (6.34-38.38) | 16.50 (5.67-34.43) |
| | 70% infection / 70% disease | 14.36 (4.92-30.06) | 19.21 (6.62-39.83) | 19.45 (6.69-40.43) | 18.27 (6.29-38.02) | 19.37 (6.66-40.32) | 17.43 (5.99-36.37) |
| | 90% infection / 90% disease | 15.75 (5.40-32.97) | 20.85 (7.19-43.23) | 21.15 (7.28-43.97) | 19.88 (6.85-41.39) | 21.13 (7.27-44.00) | 19.08 (6.55-39.81) |

### Appendix F. Lassa-X spillover, geospatial spread, transmission and vaccination

We use a five-step approach to model the initial emergence and subsequent geospatial spread of Lassa-X across West Africa, and to estimate the health-economic impacts of reactive "100 Days Mission" vaccination campaigns (detailed below). First, we assume that Lassa-X would emerge following a single spillover event. Using our LASV spillover risk map, we define the probability of Lassa-X emergence in each district as the proportion of all LASV spillover events occurring in that district. Second, to simulate subsequent between-district spread, we use a gravity model fit to Ebola case data that accounts for population size, distance, international border crossings, and presence/absence of infection.[48] Third, to quantify the inherent transmissibility of Lassa-X, we use published Ebola case data to generate a pool of longitudinal estimates for the effective reproduction number ($R_t$) at the district level, representing plausible scenarios of Lassa-X outbreaks in districts of West Africa and behavioural responses to them.[49] Fourth, to simulate within-district Lassa-X transmission, we use a compartmental Susceptible-Exposed-Infectious-Recovered (SEIR) model with vaccination. For each simulation, population size-adjusted $R_t$ curves are randomly drawn from the pool of estimates generated in step 3 and used to produce time-varying transmission rate estimates for the SEIR model. Then, vaccines are randomly allocated in each district beginning either 100 days or 160 days from the initial detection of Lassa-X, and at rates corresponding to either 2.5%, 20% or 40% of each district's population per year. Finally, as with Lassa fever, we input Lassa-X infection estimates into our health-economic model to estimate the burden of Lassa-X outbreaks and, in turn, the burden averted due to vaccination.

##### *F.1. Lassa-X spillover and geospatial spread*

Spillover risk for Lassa-X is assumed to be directly proportional to LASV spillover risk as estimated in our geospatial risk map. Specifically, the probability $p_d$ of a Lassa-X spillover event occurring in any district $d$ is calculated as the estimated number of Lassa virus spillovers $L_d$ occurring in that district in the median simulation ($\varepsilon = 50$) divided by the total number of spillovers across all districts,

$$p_d = \frac{L_d}{\sum_{i=1}^{183} L_i}$$

Spillover probability in each district is visualised in **Figure 3** in the main text. Probabilities of spillover are on average highest in districts of Nigeria, ranging from $p = 0.6\%$ in Bayelsa to $p = 3.3\%$ in Kano. By contrast, in Mauritania probabilities of spillover range from just $p = 0.005\%$ in Inchiri to $p = 0.2\%$ in Nouakchott. We assume that Lassa-X spillover occurs only once, but that the virus subsequently spreads stochastically throughout West Africa, with between-district transmission dynamics of Lassa-X being similar to those of Ebola virus during the 2013/16 West Africa epidemic.

In an analysis of the district-level spatial spread of Ebola during the 2013/16 epidemic, Kramer *et al.* compared a range of candidate models and found generalised gravity models were best able to reproduce the geospatial spread of the virus.[48] In their analysis, and in agreement with similar work from Dudas *et al.*,[50] a gravity model including the distance between districts, their population sizes and the crossing of international borders was the best fitting model. Specifically, a model penalizing transmission from "core countries" (Guinea, Liberia and Sierra Leone) to "non-core countries" produced the best fit. However, the definition of core countries or geographic regions impacted by an outbreak can only be defined retrospectively. To prospectively simulate the spread of Lassa-X, we therefore use the following model,

$$\begin{cases} \beta_3 \dfrac{1}{1+e^{\beta_0+\beta_1 \frac{\delta_{ij}}{(N_i N_j)^{\beta_2}}}}, & \text{if crossing border} \\ \dfrac{1}{1+e^{\beta_0+\beta_1 \frac{\delta_{ij}}{(N_i N_j)^{\beta_2}}}}, & \text{if not crossing border} \end{cases}$$

where $\delta_{ij}$ is the Euclidean distance between any two districts $i$ and $j$ (latitude and longitude estimates for each administrative area were extracted from the online geographical database GeoNames),[51] the gravity term is the product of the district population sizes $N_i N_j$ (using rasterised estimates from the LASV spillover model) modulated by an exponent $\beta_2$, and $\beta_3$ describes the relative likelihood of spread to districts across an international border. Using this model and parameters estimated from Kramer et al. ($\beta_0 = 5.166$, $\beta_1 = 157.1$, $\beta_2 = 0.189$, $\beta_3 = 0.507$), a matrix is produced describing the daily probabilities of transmission between all pairwise combinations of districts included in the model.

For each of $\phi = 100$ Lassa-X outbreak simulations, an initial spillover location $d_\phi$ is drawn randomly based on $p_d$, and daily binomial draws from the gravity matrix are used to determine the subsequent spread of Lassa-X to all other districts, generating probabilistic trajectories of between-district spread that are distinct in each simulation. This stochastic process continues for two years, after which point it is assumed that the cross-district spread of Lassa-X is contained (representing a similar timeline to the containment of Ebola from the 2013/16 outbreak).

The proportion of simulations in which each district experienced a Lassa-X outbreak is shown in **Figure F.1**. Upon the establishment of Lassa-X in each district, its transmission dynamics are then simulated independently using Ebola-like transmission dynamics.

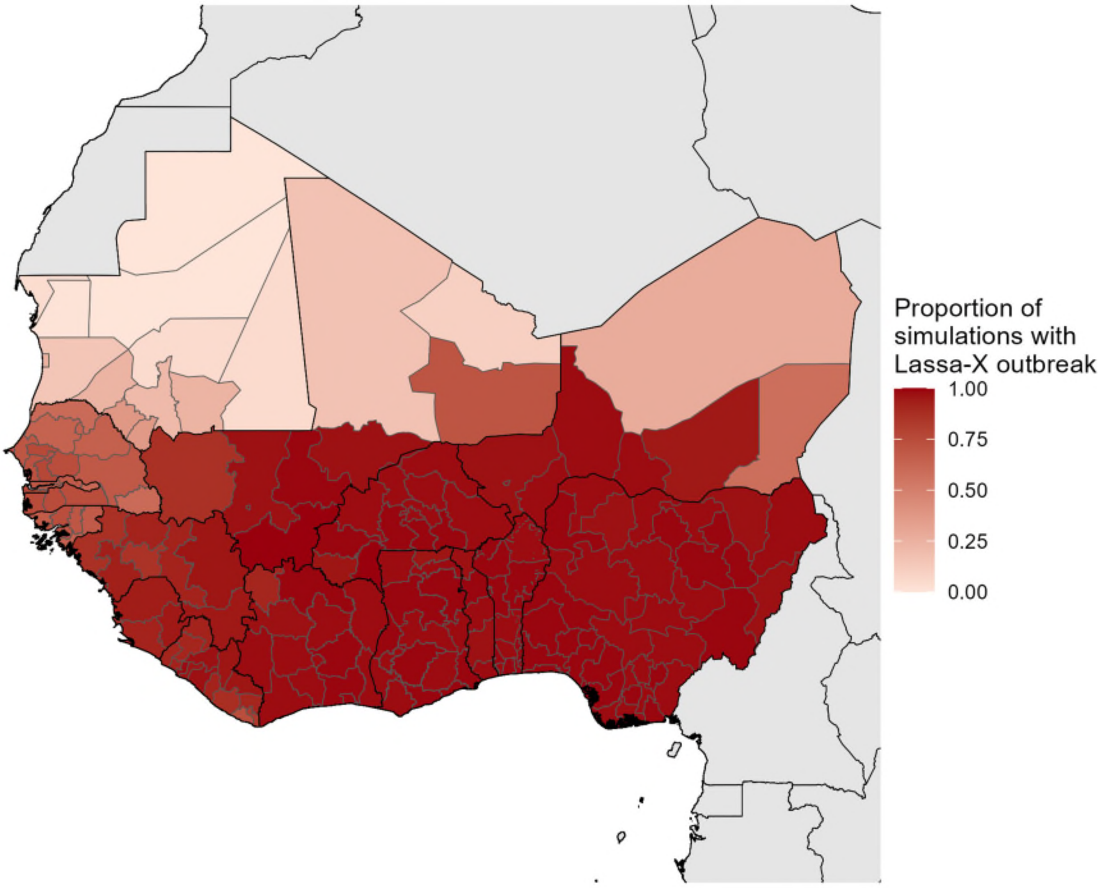


***Figure F.1. Geospatial spread of Lassa-X.*** *Map of West Africa showing the percentage of simulations in which each district included in the model experienced a Lassa-X outbreak. Values range from a minimum of 0% of simulations in Adrar (Mauritania) and Tiris Zemmour (Mauritania) to a maximum of 100% of simulations in Bamako (Mali), Sikasso (Mali) and Sud-Ouest (Burkina Faso). The following countries experienced outbreaks in every district in at least 95% of simulations: Benin, Burkina Faso, Ghana, Nigeria and Togo.*

#### F.2. Quantifying Ebola-like transmission dynamics

District-level transmission dynamics of Lassa-X are assumed to be similar to district-level transmission dynamics of Ebola virus during the West African outbreak of 2013/16. Publicly available Ebola infection data were sourced from Garske et al. and include daily counts of all confirmed and suspected infections by district of residence.[49] These outbreaks are visualised at the district level in **Figure F.2**.

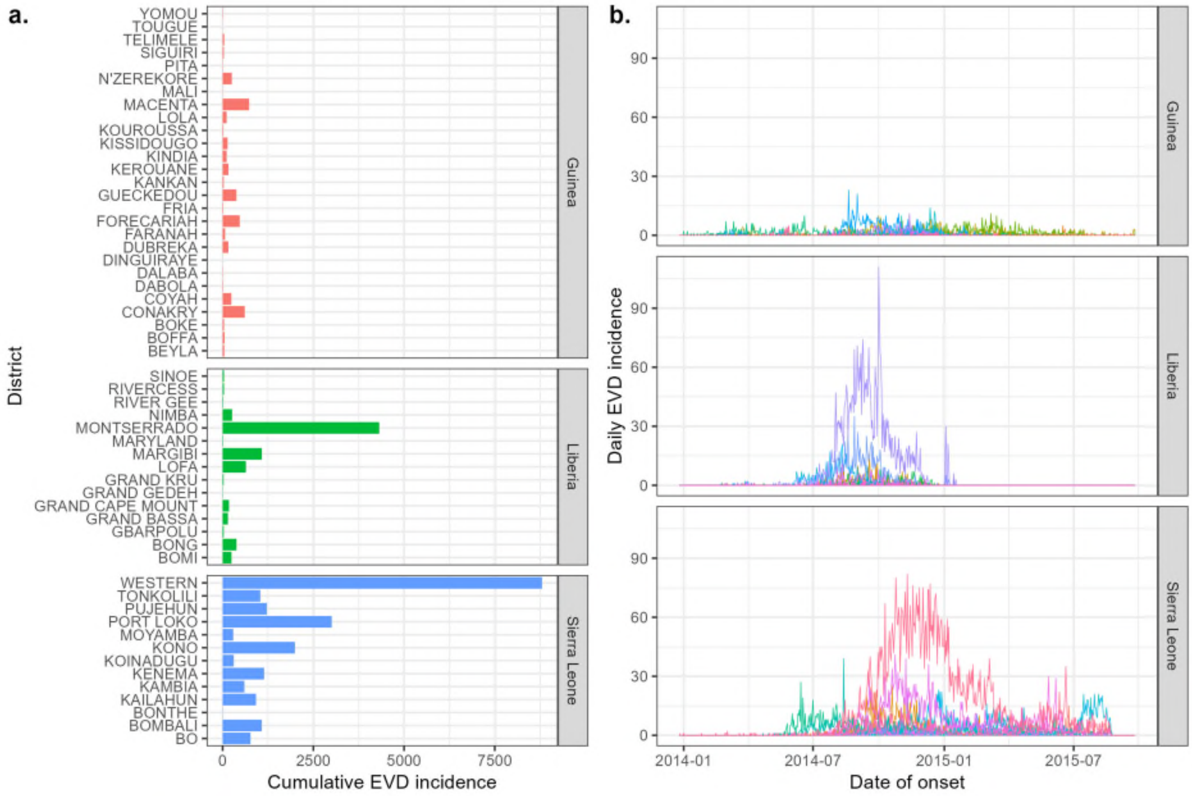


***Figure F.2. District-level Ebola case data.*** *Ebola virus disease (EVD) incidence from WHO case reports from the 2013/16 West African outbreak in Guinea, Liberia and Sierra Leone, as per data from Garske et al. (**a**) Cumulative infection incidence by district. (b) Daily infection incidence by district, where each coloured line represents a distinct district.*

Ebola infection data are fit using the R package EpiEstim to estimate $R_t$ (the time-varying reproduction number) for each district.[52] For simplicity, we assume that only the first infection in each district was imported, and that all others resulted from local transmission. However, for each instance in which there is a lag between any two infections greater than the 95th percentile of the serial interval distribution, outbreaks are severed and considered as distinct outbreaks, with the first infection after the lag assumed to be the first (imported) infection of a novel subsequent outbreak. The serial interval is assumed to follow a Gamma distribution, as estimated by the WHO Ebola Response Team.[53] From the final data point of each $R_t$ curve, we extrapolate linearly to $R_t = 0$ over 50 days to smooth extinctions out over time. Outbreaks with fewer than 10 infections were removed due to difficulty estimating $R_t$ for small outbreaks, resulting in the inclusion of a total of 54 outbreaks. Examples of $R_t$ curves generated for ten of these outbreaks using this method are visualised in **Figure F.3.**

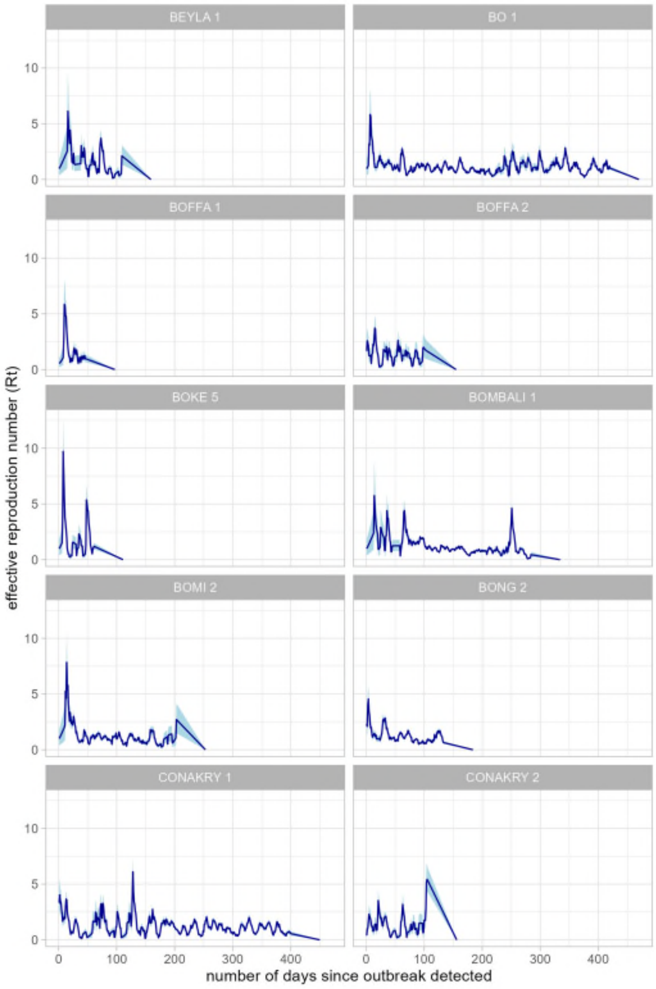


***Figure F.3. Ebola-like transmission dynamics.*** *Selection of ten $R_t$ curves estimated from Ebola infection data from Garske et al. using the R package EpiEstim. Dark blue lines represent means and light blue bands represent 95% confidence intervals, and each panel represents a distinct outbreak. Note that these 10 outbreaks originate from 8 districts, because districts with gaps between reported cases exceeding the 95th percentile of the serial interval distribution are considered to have multiple distinct outbreaks, each with a distinct $R_t$ curve.*

We estimate, for each day $t$ of each outbreak $k$, a distinct gamma distribution $\Gamma_{k,t}$ describing uncertainty in the estimated daily value of $R_t$. To recreate longitudinal $R_t$ curves that account for this uncertainty, we randomly draw $n = 1{,}000$ quantiles from within the interquartile range $q_n \sim U[0.25, 0.75]$ and, for each outbreak, draw values from $\Gamma_{k,t}$ corresponding to quantile $q_n$ over each day of the outbreak. This process results in a final pool of 54,000 distinct $R_t$ curves.

##### F.3. Compartmental model and simulation of Lassa-X outcomes

*Compartmental model*

Dynamics of Lassa-X infection and transmission are described for each district using a modified SEIR (S = Susceptible, E = Exposed, I = Infectious, R = Recovered) model accounting for potential vaccination. For each district we assume a stable population size over time,

$$N_d = S_d + S_d^V + E_d + E_d^V + I_d + I_d^V + R_d + R_d^V$$

where superscript $V$ denotes vaccinated individuals and $N_d$ corresponds to rasterised estimates used in the LASV spillover model. For any district, this model is described using a system of deterministic ordinary differential equations (ODEs),

$$\frac{dS}{dt} = -\beta(t)S(t)\frac{I(t) + I^V(t)}{N(t)} - (1-\omega)\mu_v(t)\frac{S(t)}{S(t)+R(t)}$$
$$\frac{dE}{dt} = \beta(t)S(t)\frac{I(t) + I^V(t)}{N(t)} - \alpha E(t)$$
$$\frac{dI}{dt} = \alpha E(t) - \gamma I(t)$$
$$\frac{dR}{dt} = \gamma I(t) - (1-\omega)\mu_v(t)\frac{R(t)}{S(t)+R(t)}$$
$$\frac{dS^V}{dt} = (1-\omega)\mu_v(t)\frac{S(t)}{S(t)+R(t)} - \left(1 - VE_{infect}\right)\beta(t)S^V(t)\frac{I(t) + I^V(t)}{N(t)}$$
$$\frac{dE^V}{dt} = \left(1 - VE_{infect}\right)\beta(t)S^V(t)\frac{I(t) + I^V(t)}{N(t)} - \alpha E^V(t)$$
$$\frac{dI^V}{dt} = \alpha E^V(t) - \gamma I^V(t)$$
$$\frac{dR^V}{dt} = (1-\omega)\mu_v(t)\frac{R(t)}{S(t)+R(t)} + \gamma I^V(t)$$

where Lassa-X infection is characterised by a non-infectious incubation period of $\alpha^{-1}$ days, an infectious period of $\gamma^{-1}$ days, and transmission rate $\beta$. The latter is assumed to vary each day based on time-varying estimates of Ebola virus transmission dynamics (see **Appendix F.2**), where daily $R_t$ values are translated into continuous time using a linear interpolating function and applied as

$$\beta(t) = R_t \times \gamma$$

Other epidemiological characteristics of Lassa-X are assumed identical to LASV, so mean estimates from **Table C.1** are used for the other viral parameters ($\alpha^{-1} = 10.3$ days, $\gamma^{-1} = 11.3$ days).

A total of $(1-\omega) \times \mu_v$ individuals are vaccinated per day, where $\mu_v$ represents the daily number of doses delivered and depends on the vaccination scenario $v$ and simulation time $t$ (see below), and $\omega$ represents the share of wasted doses (fixed at 10%, as for LASV). Vaccines are distributed randomly among susceptible and recovered individuals, assuming that infected individuals are not vaccinated due to symptoms potentially contra-indicating vaccination. We also assume no re-vaccination among vaccinated individuals nor any immune waning. Finally, vaccine efficacy against infection is modelled as a reduction in acquisition risk by a factor $VE_{infect}$ among vaccinated individuals exposed to infection.

*Vaccination scenarios*

Vaccination scenarios considered for Lassa-X are distinct from those considered for LASV, and represent high levels of vaccine investment in line with the 100 Days Mission, as described in the main text. These scenarios reflect three considered rates of annual vaccine uptake (2.5%, 20% or 40% of each district's population per 365 day period) and two delays to vaccination initiation (100 or 160 days from initial outbreak detection in the district where initial spillover occurred). We further consider three levels of vaccine efficacy against infection, $VE_{infect}$ (0%, 70% or 90%).

*Simulation initialisation*

For each Lassa-X outbreak simulation $\phi$, the gravity model described above determines the districts in which Lassa-X transmission occurs and the timing of Lassa-X emergence in each district. Lassa-X transmission is first simulated in the district $d_L$ where the initial spillover event occurred, with simulation time beginning at $t_{d_L} = 0$, which is assumed to correspond with the first detection of a hospitalised case of Lassa-X in that district (i.e. with the presence of virus becoming known to public health authorities). In all other districts $d$ where Lassa-X becomes established in a given outbreak simulation, each outbreak begins at time $t_d = 0$, while the timing relative to the overall outbreak (i.e. the number of days elapsed since initial detection of Lassa-X in $d_L$) is given by $T_d$.

To represent improvement in case ascertainment with increasing $T$, the average outbreak size in a given district upon the initial detection of the outbreak in that district ($X_T$) is assumed to decrease exponentially over time, from an average of $X_0 = 50$ infections upon detection of the index outbreak in $d_L$ to an average of $X_{365} = 5$ infections one year on. For each district in each simulation, this parameter is drawn randomly from a Poisson distribution,

$$X_T \sim Pois(\lambda_T)$$

where the rate parameter $\lambda_T$ is defined as

$$\lambda_T = X_0 e^{-rT}$$

and where the daily rate of decline $r$ is given by

$$r = -\frac{log\left(\frac{X_{365}}{X_0}\right)}{365}$$

The number of infected individuals upon outbreak detection is evenly distributed among exposed and infectious compartments. For outbreaks detected in a district after vaccination has already begun (e.g. when $T_d > 100$ in the scenario assuming rollout of vaccination within 100 days of initial outbreak detection), the number of individuals already vaccinated at time $T_d$ is calculated as the number of days elapsed since vaccination began in vaccine scenario $v$, $D_v$, times the number of individuals vaccinated daily in that district $\mu_{d,v}$. Finally, we assume that no individuals already vaccinated upon outbreak detection have been previously exposed to Lassa-X.

This results in the following initial conditions:

$$S_d(T_d) = N_d - X_T - (1-\omega)\mu_{d,v} D_v$$
$$E_d(T_d) = I_d(T_d) = \frac{X_T}{2}$$
$$S_d^V(T_d) = (1-\omega)\mu_{d,v} D_v$$
$$R_d(T_d) = E_d^V(T_d) = I_d^V(T_d) = R_d^V(T_d) = 0$$

For each simulation in each district, it is also necessary to apply one of the 54,000 district-level $R_t$ estimates derived from Ebola case data (described above). However, in the Ebola case data from Garske et al. underlying our $R_t$ estimates, the cumulative incidence of Ebola infection at the district level is strongly correlated with district population size (**Figure F.4**). For this reason, we binned both the districts corresponding to the 54 outbreaks included in our bank of $R_t$ curves and the 183 districts included in our Lassa infection model into three equal-sized groups according to relative population sizes (small, medium, large). Thus, when simulating Lassa-X transmission in districts with small, medium or large population sizes, we randomly drew $R_t$ curves only from districts also having small, medium or large population sizes, respectively.

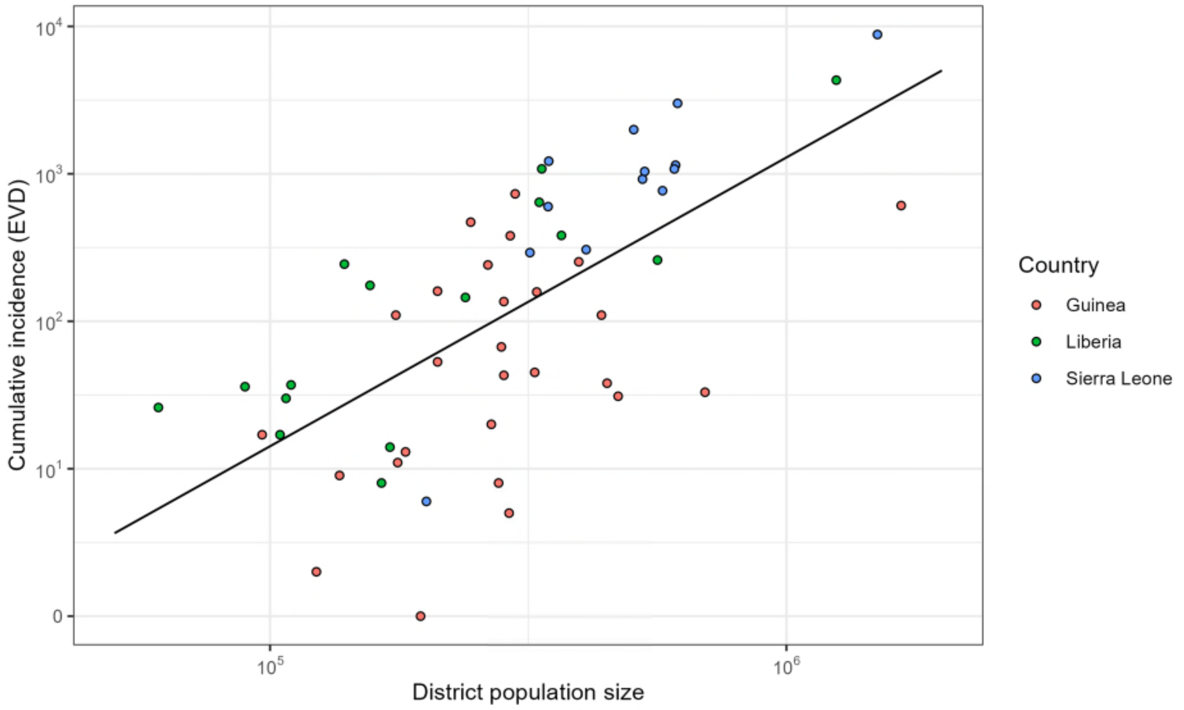


***Figure F.4. More populous districts experience larger outbreaks.*** *Correlation between each district's population size and cumulative Ebola incidence, as per WHO case report data from the 2013/16 West African outbreak reported in Garske et al. EVD = Ebola virus disease.*

#### *Numerical integration*

Based on these initialisation conditions, Lassa-X outbreak simulation was carried out through numerical ODE integration, using the R package deSolve, both without vaccination and with each of the Lassa-X vaccination scenarios considered.[54]

For a given Lassa-X outbreak simulation $\phi$, we calculate the cumulative incidence of Lassa-X infection in each district in the absence of vaccination as

$$incX_d = \int_{t_{d=0}}^{t_{max}} \beta_d(t) S_d(t) \frac{I_d(t)}{N_d(t)} dt$$

where $t_{max}$ corresponds to the last time-point in the $R_t$ curve sampled for that simulation, and hence the end of Lassa-X transmission in that district.

For that same outbreak in that district, for each vaccination strategy $v$ we then calculate the cumulative incidence of Lassa-X infection among unvaccinated individuals,

$$incS_{v,d} = \int_{t_{d=0}}^{t_{max}} \beta_d(t) S_{v,d}(t) \frac{I_{v,d}(t) + I_{v,d}^V(t)}{N_d(t)} dt$$

and among vaccinated individuals,

$$incV_{v,d} = \int_{t_{d=0}}^{t_{max}} \left(1 - VE_{infect}\right) \beta_d(t) S_{v,d}^V(t) \frac{I_{v,d}(t) + I_{v,d}^V(t)}{N_d(t)} dt$$

##### *Health-economic outcomes*

Health-economic outcomes for Lassa-X, and outcomes averted due to vaccination against Lassa-X, are calculated using the same health-economic model used for LASV (**Appendix D**). For a given Lassa-X outbreak simulation $\phi$, and using the same notation as for LASV, from cumulative Lassa-X incidence estimates we report the total number of Lassa-X infections in the absence of vaccination,

$$U_d = incX_d$$

the total number of Lassa-X infections averted due to vaccination,

$$A_{v,d} = incX_d - \left(incS_{v,d} + incV_{v,d}\right)$$

and the total number of Lassa-X infections occurring in vaccinated individuals,

$$V_{v,d} = incV_{v,d}$$

The same methods and input parameters are used for Lassa-X as for LASV (see **Table D.1**), and the same outcomes are reported. However, for Lassa-X we consider alternative values for the probability of severe disease and hospitalisation $P(hospital|infection)$. In our base case analysis, to consider a "worst case scenario" for Lassa-X, in each Monte Carlo simulation we increase this parameter by a factor of ten. We also run simulations using the same value of this parameter as for LASV, and simulations where we reduce it by a factor of ten, representing an alternative scenario where Lassa-X transmits much more readily from person-to-person than LASV, but causes much milder disease.

#### Appendix G. Supplementary results for Lassa-X

##### *G.1. Lassa-X burden in the absence of vaccination*

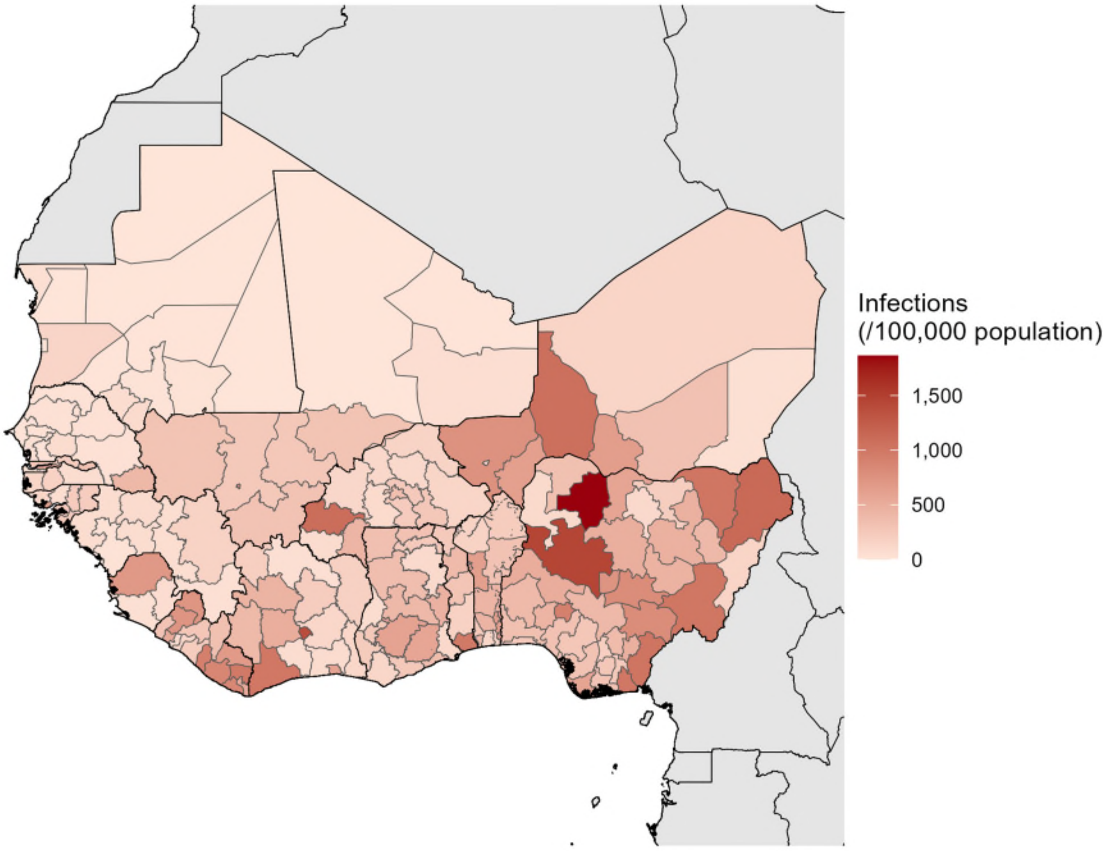


***Figure G.1. Cumulative Lassa-X infection incidence in the absence of vaccination.*** *A map of West Africa showing the mean cumulative incidence of Lassa-X infection per 100,000 individuals at the district level across 100 outbreak simulations.*

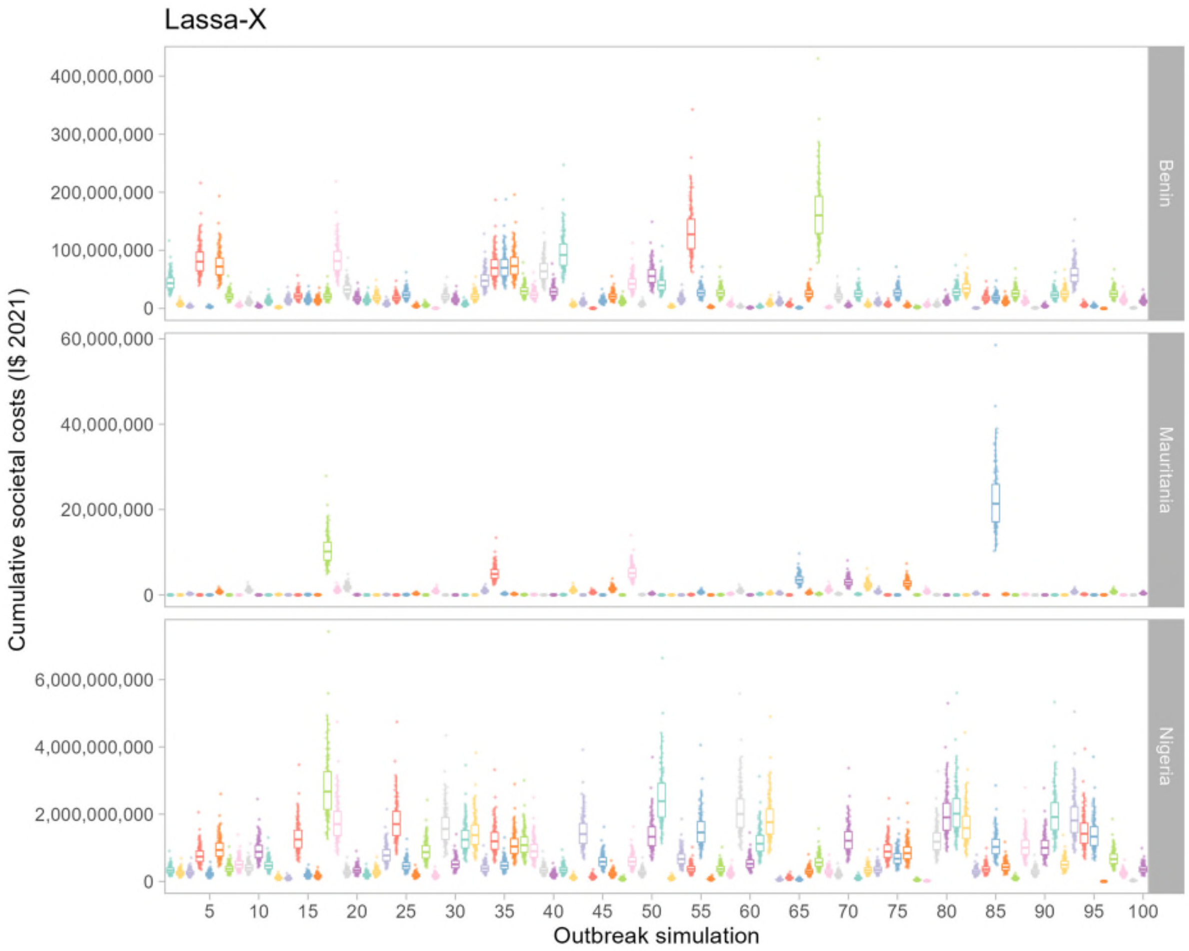


***Figure G.2. Raw Lassa-X simulation output data in the absence of vaccination.*** *Raw data points overlaid with boxplots depicting estimates of the cumulative societal costs incurred by Lassa-X across all simulations. The y-axis depicts* $SC_{\varepsilon,\rho,c}$*, an estimate of the cumulative societal costs from Lassa-X outbreak simulation* $\varepsilon$ *(x-axis), Monte Carlo simulation* $\rho$ *(points) and country* $c$ *(panels). Results are shown for three selected countries having high estimated burden of Lassa-X (Nigeria), moderate burden (Benin) or low burden (Mauritania). These estimates correspond to baseline parameter assumptions of a 3% annual discounting rate for future monetary costs, a probability of sequelae of 62% among patients discharged from hospital, and a ten-fold greater risk of hospitalisation relative to LASV. Upper and lower boxplot whiskers extend no further than values 150% larger or smaller, respectively, than the interquartile range. I$ = International dollar.*

***Table G.1. Cumulative health burden of Lassa-X by country in the absence of vaccination.*** *All figures represent means (95% uncertainty intervals) over approximately two years across 100 runs of the transmission model and 100 runs of the health-economic model, for the baseline scenario assuming a probability of sequelae of 62% among patients discharged from hospital, and a probability of hospitalisation ten times greater than for LASV. DALY = disability-adjusted life year, K = thousand, M = million.*

| | Infections | Mild/moderate cases | Hospitalisations | Deaths | Sequelae | DALYs |
|---|---|---|---|---|---|---|
| *Benin* | 40.8K (762.7-170.3K) | 8.1K (145.2-38.3K) | 3.6K (66.2-16.0K) | 589.4 (9.5-2.9K) | 1.8K (34.6-8.2K) | 31.2K (537.1-156.4K) |
| *Burkina Faso* | 65.1K (2.6K-520.0K) | 13.0K (422.1-87.2K) | 5.7K (204.8-40.7K) | 940.0 (26.3-6.1K) | 2.9K (103.4-21.0K) | 49.6K (1.4K-322.9K) |
| *Côte d'Ivoire* | 104.5K (1.8K-936.6K) | 20.8K (276.7-185.3K) | 9.1K (138.4-80.2K) | 1.5K (17.7-13.3K) | 4.7K (70.3-41.0K) | 76.4K (907.8-686.5K) |
| *Ghana* | 113.9K (1.9K-805.3K) | 22.7K (400.0-161.3K) | 9.9K (168.6-70.0K) | 1.6K (27.2-11.2K) | 5.1K (87.8-35.7K) | 86.0K (1.5K-597.6K) |
| *Guinea* | 8.9K (249.8-58.1K) | 1.8K (41.1-10.7K) | 770.8 (21.0-4.9K) | 127.8 (2.5-772.0) | 398.1 (10.7-2.5K) | 6.7K (140.7-39.7K) |
| *Gambia* | 3.6K (0.0-33.3K) | 709.7 (0.0-6.4K) | 310.0 (0.0-2.7K) | 51.4 (0.0-451.7) | 160.1 (0.0-1.4K) | 2.8K (0.0-24.7K) |
| *Guinea-Bissau* | 3.1K (0.0-15.9K) | 612.3 (0.0-3.7K) | 267.5 (0.0-1.5K) | 44.3 (0.0-288.2) | 138.1 (0.0-787.9) | 2.3K (0.0-14.7K) |
| *Liberia* | 14.7K (184.2-79.6K) | 2.9K (34.8-15.7K) | 1.3K (16.0-6.6K) | 212.4 (2.2-1.2K) | 661.8 (8.4-3.3K) | 11.0K (115.9-61.0K) |
| *Mali* | 72.9K (1.4K-537.3K) | 14.5K (246.8-109.8K) | 6.3K (114.2-48.1K) | 1.1K (15.8-7.8K) | 3.3K (59.2-25.3K) | 55.9K (876.6-415.7K) |
| *Mauritania* | 1.2K (0.0-8.7K) | 248.4 (0.0-1.9K) | 108.5 (0.0-814.3) | 18.0 (0.0-149.4) | 56.0 (0.0-425.9) | 983.3 (0.0-8.1K) |
| *Niger* | 142.1K (1.4K-1.1M) | 28.3K (213.5-251.2K) | 12.4K (106.6-105.7K) | 2.1K (12.7-19.0K) | 6.4K (54.6-54.7K) | 113.5K (723.5-1.1M) |
| *Nigeria* | 1.1M (41.2K-3.2M) | 214.1K (7.9K-756.0K) | 93.5K (3.7K-303.7K) | 15.5K (503.3-60.0K) | 48.3K (1.9K-157.9K) | 744.9K (25.6K-2.8M) |
| *Senegal* | 7.7K (0.0-47.5K) | 1.5K (0.0-9.5K) | 668.4 (0.0-4.1K) | 110.8 (0.0-698.9) | 345.2 (0.0-2.1K) | 6.3K (0.0-40.1K) |
| *Sierra Leone* | 18.3K (42.8-38.8K) | 3.7K (7.0-9.1K) | 1.6K (3.5-3.6K) | 264.5 (0.4-726.6) | 823.9 (1.8-1.9K) | 13.7K (23.2-37.7K) |
| *Togo* | 47.9K (608.8-247.2K) | 9.5K (97.5-54.5K) | 4.2K (48.0-22.8K) | 691.4 (6.1-4.4K) | 2.2K (24.7-11.9K) | 35.6K (322.5-223.0K) |
| **Total** | **1.7M (230.1K-4.2M)** | **342.7K (39.6K-936.3K)** | **149.7K (19.7K-374.4K)** | **24.8K (2.4K-76.0K)** | **77.3K (9.9K-194.3K)** | **1.2M (132.5K-3.7M)** |

***Table G.2. Cumulative incidence of Lassa-X health burden (per 100,000 population) by country in the absence of vaccination.*** *All figures represent means (95% uncertainty intervals) over approximately two years across 100 runs of the transmission model and 100 runs of the health-economic model, for the baseline scenario assuming a probability of sequelae of 62% among patients discharged from hospital, and a probability of hospitalisation ten times greater than for LASV. DALY = disability-adjusted life year, K = thousand.*

| | Infections | Mild/moderate cases | Hospitalisations | Deaths | Sequelae | DALYs |
|---|---|---|---|---|---|---|
| *Benin* | 326.5 (6.1-1.4K) | 65.1 (1.2-305.9) | 28.4 (0.5-127.7) | 4.7 (0.1-23.4) | 14.7 (0.3-65.9) | 249.7 (4.3-1.3K) |
| *Burkina Faso* | 296.8 (11.6-2.4K) | 59.2 (1.9-397.3) | 25.8 (0.9-185.6) | 4.3 (0.1-27.9) | 13.3 (0.5-95.7) | 226.2 (6.6-1.5K) |
| *Côte d'Ivoire* | 405.2 (7.0-3.6K) | 80.8 (1.1-718.3) | 35.3 (0.5-310.7) | 5.9 (0.1-51.7) | 18.2 (0.3-158.8) | 296.3 (3.5-2.7K) |
| *Ghana* | 355.0 (5.8-2.5K) | 70.8 (1.2-502.8) | 30.9 (0.5-218.0) | 5.1 (0.1-34.9) | 16.0 (0.3-111.3) | 268.0 (4.7-1.9K) |
| *Guinea* | 73.8 (2.1-484.1) | 14.7 (0.3-89.1) | 6.4 (0.2-40.5) | 1.1 (0.0-6.4) | 3.3 (0.1-20.8) | 56.3 (1.2-330.8) |
| *Gambia* | 154.3 (0.0-1.4K) | 30.8 (0.0-277.2) | 13.4 (0.0-119.3) | 2.2 (0.0-19.6) | 6.9 (0.0-62.3) | 119.6 (0.0-1.1K) |
| *Guinea-Bissau* | 169.5 (0.0-880.0) | 33.8 (0.0-203.4) | 14.8 (0.0-83.9) | 2.4 (0.0-15.9) | 7.6 (0.0-43.5) | 124.9 (0.0-810.5) |
| *Liberia* | 335.4 (4.2-1.8K) | 66.9 (0.8-358.8) | 29.2 (0.4-150.8) | 4.8 (0.0-26.9) | 15.1 (0.2-76.4) | 250.6 (2.6-1.4K) |
| *Mali* | 328.3 (6.4-2.4K) | 65.5 (1.1-494.6) | 28.6 (0.5-216.8) | 4.7 (0.1-35.3) | 14.8 (0.3-114.1) | 251.7 (3.9-1.9K) |
| *Mauritania* | 28.9 (0.0-201.8) | 5.8 (0.0-45.1) | 2.5 (0.0-18.9) | 0.4 (0.0-3.5) | 1.3 (0.0-9.9) | 22.8 (0.0-188.2) |
| *Niger* | 618.8 (6.0-4.8K) | 123.4 (0.9-1.1K) | 53.9 (0.5-460.2) | 8.9 (0.1-82.6) | 27.8 (0.2-238.3) | 494.2 (3.2-4.6K) |
| *Nigeria* | 512.5 (19.6-1.5K) | 102.2 (3.8-360.8) | 44.6 (1.7-144.9) | 7.4 (0.2-28.6) | 23.1 (0.9-75.4) | 355.5 (12.2-1.4K) |
| *Senegal* | 48.5 (0.0-300.5) | 9.7 (0.0-60.2) | 4.2 (0.0-26.2) | 0.7 (0.0-4.4) | 2.2 (0.0-13.4) | 39.8 (0.0-253.9) |
| *Sierra Leone* | 276.2 (0.6-585.2) | 55.1 (0.1-136.6) | 24.1 (0.1-54.8) | 4.0 (0.0-11.0) | 12.4 (0.0-28.4) | 207.2 (0.3-568.7) |
| *Togo* | 574.5 (7.3-3.0K) | 114.6 (1.2-653.7) | 50.0 (0.6-273.2) | 8.3 (0.1-52.7) | 25.8 (0.3-143.1) | 427.0 (3.9-2.7K) |
| **Total** | **426.8 (57.1-1.1K)** | **85.1 (9.8-232.5)** | **37.2 (4.9-93.0)** | **6.2 (0.6-18.9)** | **19.2 (2.5-48.3)** | **307.2 (32.9-909.1)** |

***Table G.3. Cumulative economic burden of Lassa-X by country in the absence of vaccination.*** *All figures represent means (95% uncertainty intervals) over approximately two years across 100 runs of the transmission model and 100 runs of the health-economic model, for the baseline scenario assuming a probability of sequelae of 62% among patients discharged from hospital, and a probability of hospitalisation ten times greater than for LASV. Monetary costs are reported in International dollars (2021) and future costs are discounted at 3%/year. Catastrophic and impoverishing expenditures are reported as the number of individuals facing those costs. DALY = disability-adjusted life year, VSL = value of statistical life, K = thousand, M = million, B = billion.*

| | Treatment costs, government-reimbursed ($ 2021) | Costs of care, out-of-pocket ($ 2021) | Catastrophic expenditures (N) | Impoverishing expenditures (N) | Productivity losses ($ 2021) | Monetised DALYs ($ 2021) | VSL ($ 2021) |
|---|---|---|---|---|---|---|---|
| *Benin* | 5.6M (104.9K-25.3M) | 2.2M (41.2K-9.9M) | 3.5K (64.6-15.6K) | 2.8K (51.4-12.4K) | 15.4M (251.9K-76.4M) | 6.2M (107.4K-31.3M) | 200.6M (3.2M-995.2M) |
| *Burkina Faso* | 10.0M (360.4K-71.5M) | 2.6M (94.2K-18.7M) | 5.0K (181.2-36.0K) | 3.3K (118.6-23.6K) | 10.9M (311.2K-70.8M) | 7.2M (209.7K-46.9M) | 62.5M (1.7M-406.7M) |
| *Côte d'Ivoire* | 15.7M (237.9K-138.2M) | 4.5M (68.2K-39.5M) | 8.7K (132.5-76.7K) | 7.7K (116.7-67.6K) | 64.9M (764.0K-569.8M) | 19.8M (235.8K-178.1M) | 843.4M (9.9M-7.4B) |
| *Ghana* | 17.3M (293.3K-121.6M) | 4.8M (81.2K-33.7M) | 9.5K (162.3-67.3K) | 7.0K (119.7-49.7K) | 82.3M (1.4M-561.6M) | 38.4M (678.3K-266.5M) | 948.5M (15.7M-6.5B) |
| *Guinea* | 1.1M (29.9K-6.9M) | 600.5K (16.3K-3.8M) | 769.9 (21.0-4.9K) | 663.1 (18.1-4.2K) | 2.0M (39.2K-12.0M) | 1.1M (23.6K-6.7M) | 27.5M (528.2K-166.2M) |
| *Gambia* | 588.2K (0.0-5.2M) | 95.1K (0.0-842.5K) | 268.6 (0.0-2.4K) | 215.6 (0.0-1.9K) | 785.0K (0.0-6.9M) | 764.2K (0.0-6.8M) | 3.3M (0.0-29.0M) |
| *Guinea-Bissau* | 360.5K (0.0-2.0M) | 229.5K (0.0-1.3M) | 266.8 (0.0-1.5K) | 208.4 (0.0-1.2K) | 585.9K (0.0-3.8M) | 114.9K (0.0-745.8K) | 2.5M (0.0-16.0M) |
| *Liberia* | 1.9M (23.9K-9.9M) | 920.3K (11.5K-4.8M) | 1.3K (15.9-6.6K) | 923.1 (11.5-4.8K) | 1.6M (17.2K-9.1M) | 1.4M (14.9K-7.8M) | 7.4M (75.3K-40.8M) |
| *Mali* | 11.4M (205.0K-86.4M) | 2.6M (47.3K-20.0M) | 5.8K (104.5-44.1K) | 4.9K (87.6-37.0K) | 16.7M (251.1K-125.7M) | 3.3M (52.1K-24.7M) | 67.7M (1.0M-503.8M) |
| *Mauritania* | 175.6K (0.0-1.3M) | 64.6K (0.0-484.2K) | 104.4 (0.0-783.9) | 97.4 (0.0-731.0) | 535.0K (0.0-4.4M) | 282.9K (0.0-2.3M) | 10.2M (0.0-84.9M) |
| *Niger* | 19.7M (170.0K-168.7M) | 7.5M (64.6K-64.2M) | 12.3K (105.6-104.7K) | 6.0K (51.6-51.2K) | 7.4M (47.3K-68.4M) | 11.3M (72.1K-105.2M) | 61.2M (380.1K-565.6M) |
| *Nigeria* | 120.7M (4.7M-392.9M) | 87.1M (3.4M-283.5M) | 93.5K (3.7K-303.7K) | 64.7K (2.5K-210.0K) | 517.7M (17.0M-2.0B) | 93.3M (3.2M-356.1M) | 7.8B (253.5M-30.2B) |
| *Senegal* | 1.0M (0.0-6.4M) | 449.3K (0.0-2.8M) | 652.7 (0.0-4.0K) | 590.5 (0.0-3.7K) | 2.4M (0.0-15.4M) | 2.1M (0.0-13.5M) | 37.8M (0.0-238.6M) |
| *Sierra Leone* | 2.4M (5.2K-5.4M) | 1.2M (2.5K-2.6M) | 1.6K (3.5-3.6K) | 1.2K (2.6-2.7K) | 3.0M (5.1K-8.2M) | 728.2K (1.2K-2.0M) | 11.9M (19.9K-32.7M) |
| *Togo* | 5.6M (64.0K-30.3M) | 3.6M (41.8K-19.9M) | 4.1K (47.7-22.6K) | 3.0K (34.2-16.2K) | 11.0M (98.7K-69.0M) | 4.9M (45.0K-31.0M) | 48.1M (423.5K-305.6M) |
| **Total** | **213.5M (29.7M-518.7M)** | **118.5M (12.2M-317.3M)** | **147.4K (18.5K-372.5K)** | **103.1K (13.6K-254.3K)** | **737.2M (56.4M-2.4B)** | **191.1M (18.4M-575.2M)** | **10.1B (625.9M-34.1B)** |

***Table G.4. Sensitivity of economic outcomes to discounting, the probability of sequelae, and the severity of Lassa-X relative to Lassa fever.*** *All figures represent means (95% uncertainty intervals) summed across all countries over approximately two years across 100 runs of the transmission model and 100 runs of the health-economic model, varying the discounting rate, the probability of sequelae among patients discharged from hospital, and the probability of hospitalisation per infection. Costs are reported in International dollars (2021). DALY = disability-adjusted life year, K = thousand, M = million, B = billion.*

| Discounting rate | Probability of sequelae | Severity relative to Lassa fever | Monetised DALYs ($ 2021) | Societal costs ($ 2021) |
|---|---|---|---|---|
| 0% | 17% | 0.1 | 3.3M (316.7K-9.9M) | 23.9M (2.3M-69.9M) |
| 0% | 17% | 1 | 32.3M (3.1M-97.2M) | 161.0M (15.0M-480.0M) |
| 0% | 17% | 10 | 322.4M (31.0M-970.0M) | 1.5B (143.7M-4.6B) |
| 0% | 62% | 0.1 | 3.6M (361.0K-10.8M) | 23.9M (2.3M-69.9M) |
| 0% | 62% | 1 | 35.6M (3.5M-106.7M) | 161.0M (15.0M-480.0M) |
| 0% | 62% | 10 | 355.6M (34.7M-1.1B) | 1.5B (143.7M-4.6B) |
| 3% | 17% | 0.1 | 1.8M (171.1K-5.4M) | 19.3M (1.9M-55.3M) |
| 3% | 17% | 1 | 17.4M (1.6M-52.4M) | 114.7M (11.3M-330.7M) |
| 3% | 17% | 10 | 173.3M (16.4M-522.6M) | 1.1B (105.2M-3.1B) |
| 3% | 62% | 0.1 | 2.0M (193.2K-5.9M) | 19.3M (1.9M-55.3M) |
| 3% | 62% | 1 | 19.2M (1.9M-57.7M) | 114.7M (11.3M-330.7M) |
| 3% | 62% | 10 | 191.1M (18.4M-575.2M) | 1.1B (105.2M-3.1B) |

##### *G.2. Lassa-X vaccine impact*

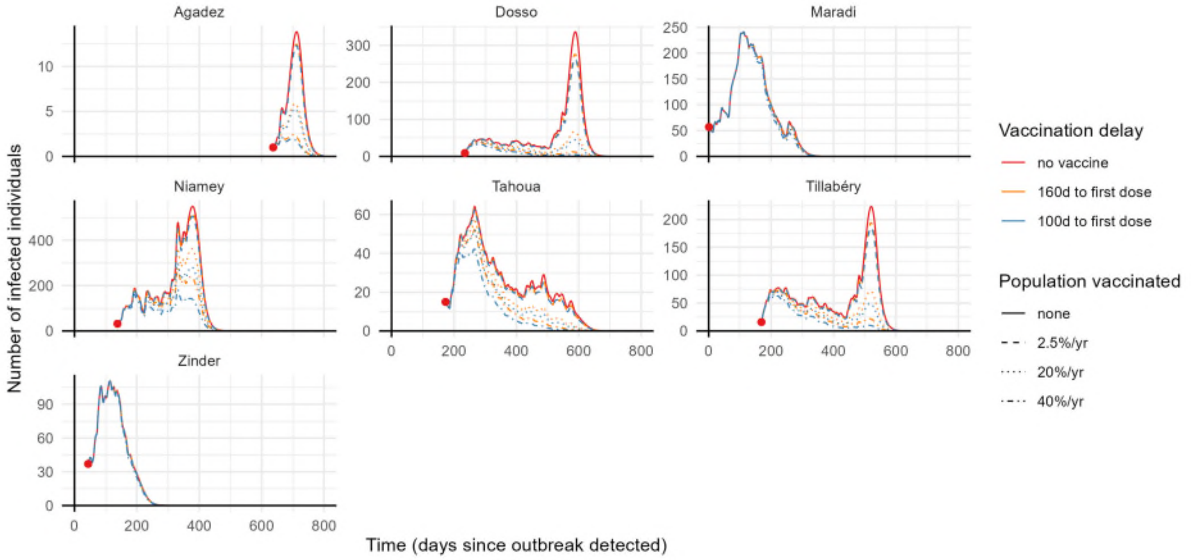


***Figure G.3. Example of how different vaccination strategies impact Lassa-X infection dynamics.*** *This plot shows the number of active Lassa-X infections over time in the districts of Niger that experienced a Lassa-X outbreak in one randomly selected outbreak simulation. Lines show how a vaccine with 70% efficacy against infection influences infection dynamics, where line colour represents the delay to vaccine rollout and line dashing represents the rate of vaccination (the proportion of each district's population vaccinated over a 1-year period). Red dots indicate the timing of the detection of Lassa-X within each district, and the number of active infections in the population upon its detection.*

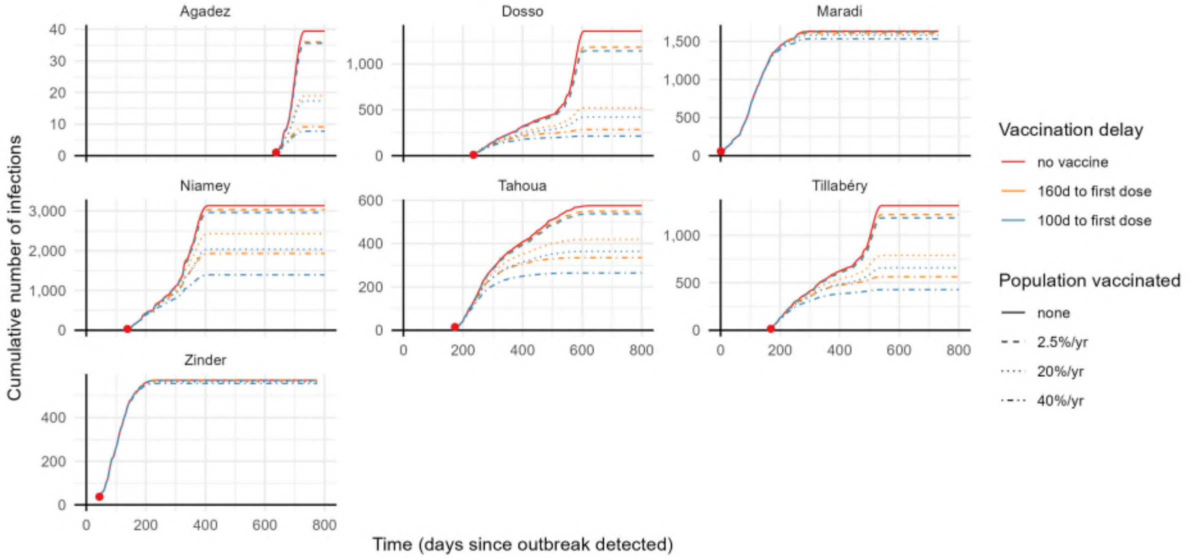


***Figure G.4. Example of how different vaccination strategies impact the cumulative burden of Lassa-X infection.*** *This plot shows the cumulative number of Lassa-X infections over time in the districts of Niger that experienced a Lassa-X outbreak in one randomly selected outbreak simulation. Lines show how a vaccine with 70% efficacy against infection influences infection dynamics, where line colour represents the delay to vaccine rollout and line dashing represents the rate of vaccination (the proportion of each district's population vaccinated over a 1-year period). Red dots indicate the*

*timing of the detection of Lassa-X within each district, and the cumulative number of infections in the population upon its detection.*

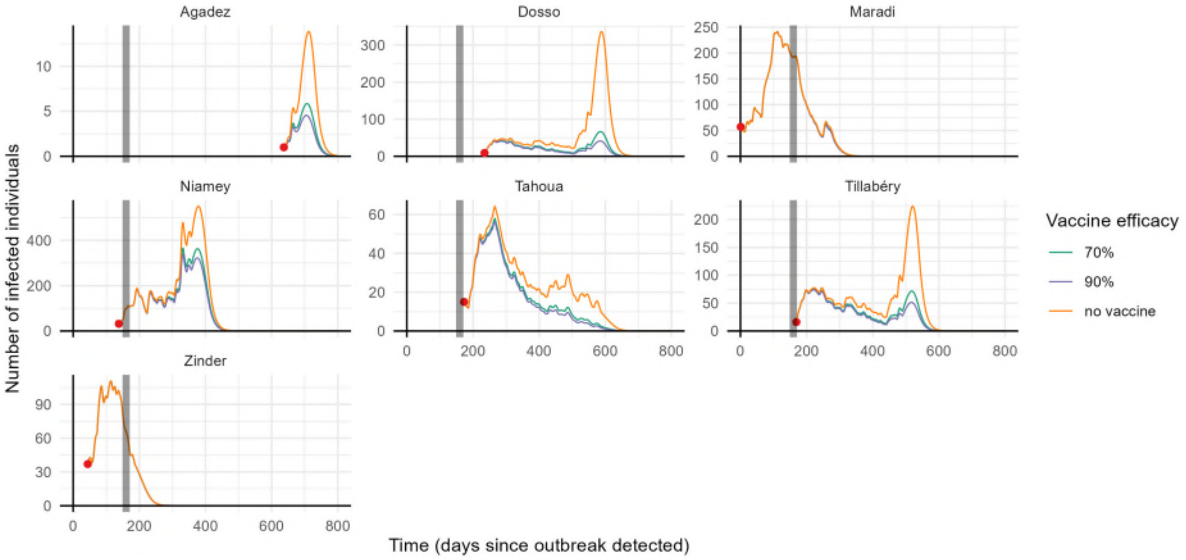


***Figure G.5. Example of how vaccine efficacy impacts Lassa-X infection dynamics.*** *This plot shows the number of active Lassa-X infections over time in the districts of Niger that experienced a Lassa-X outbreak in one randomly selected outbreak simulation. Line colour represents vaccine efficacy against infection (*$VE_{infect}$*). Here we assume a 160-day delay to the administration of the first vaccine in the population (demarcated by a grey vertical bar), and a vaccine uptake rate equivalent to 20% of the population of each district per year. Red dots indicate the timing of the detection of Lassa-X within each district, and the number of active infections upon its detection.*

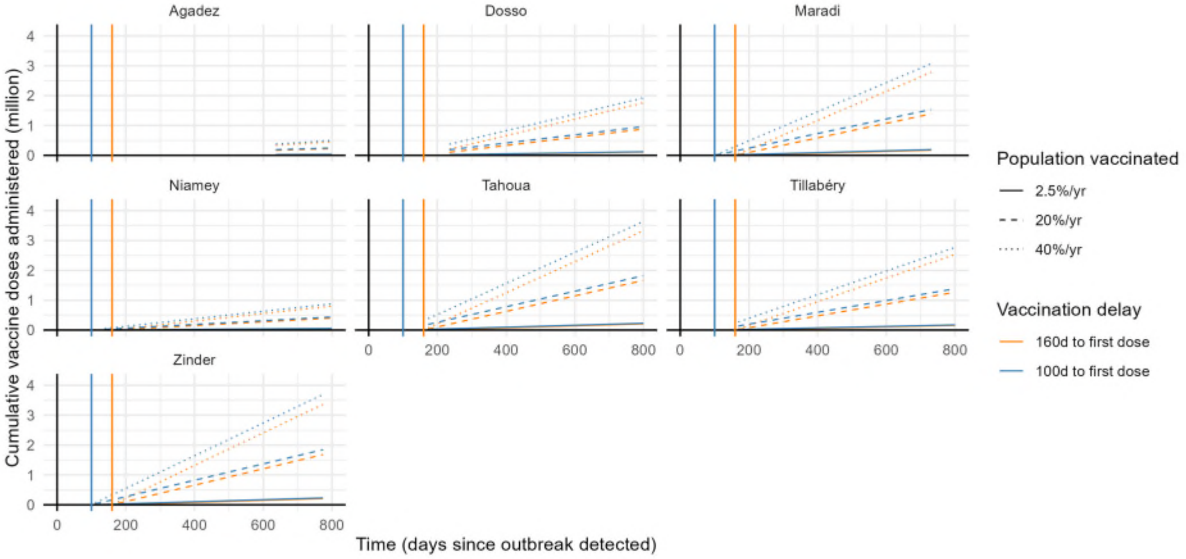


***Figure G.6. Example of vaccine administration in Lassa-X simulations.*** *The cumulative number of vaccine doses administered over time in the districts of Niger that experienced a Lassa-X outbreak in one randomly selected outbreak simulation. Lines show how the number of doses administered varies across different vaccination scenarios, where line colour represents the delay to vaccine rollout and line dashing represents the rate of vaccination (the proportion of the population vaccinated over a 1-year period). Coloured vertical bars indicate the date of administration of first vaccines, i.e. 100 days (blue line) and 160 days (orange line) from the first detection of the Lassa-X outbreak in West*

*Africa. Simulation time in each district begins with the initial detection of that district's outbreak, so the number of doses already administered upon outbreak detection is scaled accordingly.*

***Table G.5. Cumulative health burden of Lassa-X averted due to vaccine with 70% efficacy.*** *Columns represent the vaccination scenarios considered and rows represent the outcomes averted over approximately two years from the initiation of vaccine rollout. All figures represent means (95% uncertainty intervals) across 100 runs of the transmission model and 100 runs of the health-economic model simulations, for the baseline scenario assuming a probability of sequelae of 62% among patients discharged from hospital, and a probability of hospitalisation ten times greater than for LASV. LASV = Lassa virus, DALY = disability-adjusted life-year, VE = vaccine efficacy, K = thousand, M = million.*

| ***Outcome averted due to vaccination*** | **Vaccination scenario** | | | | | |
|---|---|---|---|---|---|---|
| | 2.5% of population vaccinated/year | | 20% of population vaccinated/year | | 40% of population vaccinated/year | |
| | 160d delay to first dose | 100d delay to first dose | 160d delay to first dose | 100d delay to first dose | 160d delay to first dose | 100d delay to first dose |
| **Vaccine 70% effective only against disease** ($VE_{infect} = 0\%, VE_{disease} = 70\%$) | | | | | | |
| *Lassa-X infections* | 0.0 (0.0-0.0) | 0.0 (0.0-0.0) | 0.0 (0.0-0.0) | 0.0 (0.0-0.0) | 0.0 (0.0-0.0) | 0.0 (0.0-0.0) |
| *Mild/moderate cases* | 3.9K (605.7-9.9K) | 4.8K (740.2-11.9K) | 31.1K (4.8K-78.9K) | 38.1K (5.9K-95.0K) | 62.1K (9.7K-157.7K) | 76.2K (11.8K-190.9K) |
| *Hospitalisations* | 1.7K (294.0-3.9K) | 2.1K (361.5-4.7K) | 13.6K (2.4K-31.2K) | 16.6K (2.9K-37.3K) | 27.1K (4.7K-62.3K) | 33.3K (5.8K-74.8K) |
| *Deaths* | 281.2 (35.6-811.0) | 344.5 (43.9-984.0) | 2.2K (284.6-6.5K) | 2.8K (350.9-7.9K) | 4.5K (569.2-13.0K) | 5.5K (704.9-15.7K) |
| *Sequelae* | 876.1 (150.4-2.0K) | 1.1K (185.0-2.4K) | 7.0K (1.2K-16.1K) | 8.6K (1.5K-19.5K) | 14.0K (2.4K-32.2K) | 17.2K (3.0K-39.1K) |
| *DALYs* | 14.1K (1.9K-39.2K) | 17.3K (2.4K-47.2K) | 112.9K (15.4K-313.3K) | 138.1K (18.9K-377.6K) | 225.9K (30.9K-626.2K) | 276.6K (38.0K-755.9K) |
| **Vaccine 70% effective against infection and disease** ($VE_{infect} = 70\%, VE_{disease} = 70\%$) | | | | | | |
| *Lassa-X infections* | 141.4K (27.5K-323.4K) | 183.8K (37.3K-399.2K) | 737.6K (146.5K-1.7M) | 916.0K (189.7K-2.0M) | 1.0M (200.5K-2.3M) | 1.2M (201.3K-2.7M) |
| *Mild/moderate cases* | 29.3K (4.8K-77.9K) | 37.9K (6.3K-98.2K) | 151.4K (23.2K-389.8K) | 187.2K (27.8K-469.9K) | 213.0K (30.2K-547.3K) | 253.0K (33.1K-651.2K) |
| *Hospitalisations* | 12.8K (2.4K-31.3K) | 16.6K (3.1K-38.8K) | 66.1K (11.6K-152.2K) | 81.8K (14.4K-184.8K) | 93.0K (15.4K-214.5K) | 110.5K (16.8K-252.3K) |
| *Deaths* | 2.1K (288.7-6.3K) | 2.7K (380.1-7.9K) | 11.0K (1.4K-31.4K) | 13.6K (1.6K-38.6K) | 15.4K (1.8K-44.6K) | 18.3K (2.0K-53.1K) |
| *Sequelae* | 6.6K (1.2K-16.2K) | 8.6K (1.6K-20.1K) | 34.2K (5.8K-78.9K) | 42.2K (7.2K-96.3K) | 48.1K (7.8K-112.0K) | 57.1K (8.5K-130.5K) |
| *DALYs* | 106.7K (15.6K-304.9K) | 138.0K (20.6K-383.0K) | 550.4K (74.1K-1.5M) | 679.1K (90.0K-1.9M) | 773.2K (97.8K-2.1M) | 916.4K (108.0K-2.6M) |

***Table G.6. Cumulative health burden of Lassa-X averted due to vaccine with 90% efficacy.*** *Columns represent the vaccination scenarios considered and rows represent the outcomes averted over approximately two years from the initiation of vaccine rollout. All figures represent means (95% uncertainty intervals) across 100 runs of the transmission model and 100 runs of the health-economic model simulations, for the baseline scenario assuming a probability of sequelae of 62% among patients discharged from hospital, and a probability of hospitalisation ten times greater than for LASV. LASV = Lassa virus, DALY = disability-adjusted life-year, VE = vaccine efficacy, K = thousand, M = million.*

| ***Outcome averted due to vaccination*** | **Vaccination scenario** | | | | | |
|---|---|---|---|---|---|---|
| | 2.5% of population vaccinated/year | | 20% of population vaccinated/year | | 40% of population vaccinated/year | |
| | 160d delay to first dose | 100d delay to first dose | 160d delay to first dose | 100d delay to first dose | 160d delay to first dose | 100d delay to first dose |
| **Vaccine 90% effective only against disease** ($VE_{infect} = 0\%, VE_{disease} = 90\%$) | | | | | | |
| *Lassa-X infections* | 0.0 (0.0-0.0) | 0.0 (0.0-0.0) | 0.0 (0.0-0.0) | 0.0 (0.0-0.0) | 0.0 (0.0-0.0) | 0.0 (0.0-0.0) |
| *Mild/moderate cases* | 5.0K (779.0-12.7K) | 6.1K (951.3-15.3K) | 39.9K (6.2K-101.4K) | 48.9K (7.6K-122.1K) | 79.9K (12.5K-202.7K) | 97.8K (15.2K-245.1K) |
| *Hospitalisations* | 2.2K (378.1-5.0K) | 2.7K (465.0-6.0K) | 17.4K (3.0K-40.1K) | 21.4K (3.7K-48.0K) | 34.9K (6.0K-80.1K) | 42.7K (7.4K-96.1K) |
| *Deaths* | 361.5 (45.8-1.0K) | 442.9 (56.4-1.3K) | 2.9K (365.9-8.3K) | 3.5K (451.1-10.1K) | 5.8K (731.9-16.7K) | 7.1K (902.1-20.2K) |
| *Sequelae* | 1.1K (193.4-2.6K) | 1.4K (237.7-3.1K) | 9.0K (1.5K-20.7K) | 11.0K (1.9K-25.0K) | 18.0K (3.1K-41.4K) | 22.1K (3.8K-50.2K) |
| *DALYs* | 18.1K (2.5K-50.4K) | 22.2K (3.0K-60.7K) | 145.2K (19.8K-402.8K) | 177.6K (24.3K-485.5K) | 290.3K (39.7K-805.2K) | 355.2K (48.6K-970.8K) |
| **Vaccine 90% effective against infection and disease** ($VE_{infect} = 90\%, VE_{disease} = 90\%$) | | | | | | |
| *Lassa-X infections* | 179.0K (34.7K-411.1K) | 232.2K (47.3K-505.4K) | 851.8K (169.5K-1.9M) | 1.0M (199.3K-2.4M) | 1.1M (201.0K-2.5M) | 1.3M (201.9K-2.9M) |
| *Mild/moderate cases* | 36.2K (5.9K-96.4K) | 46.8K (7.7K-121.1K) | 171.4K (25.8K-438.0K) | 209.7K (30.0K-529.6K) | 230.6K (31.5K-594.0K) | 270.2K (34.8K-699.6K) |
| *Hospitalisations* | 15.8K (2.9K-38.8K) | 20.5K (3.8K-48.0K) | 74.9K (13.1K-172.6K) | 91.6K (15.2K-206.8K) | 100.7K (15.7K-232.6K) | 118.0K (17.1K-271.0K) |
| *Deaths* | 2.6K (352.5-7.7K) | 3.4K (467.3-9.8K) | 12.4K (1.5K-35.7K) | 15.2K (1.8K-43.4K) | 16.7K (1.9K-48.8K) | 19.6K (2.1K-57.2K) |
| *Sequelae* | 8.2K (1.5K-20.1K) | 10.6K (1.9K-24.8K) | 38.7K (6.5K-89.2K) | 47.3K (7.7K-107.5K) | 52.0K (8.1K-120.6K) | 60.9K (8.7K-140.5K) |
| *DALYs* | 131.8K (19.1K-377.2K) | 170.4K (25.3K-474.0K) | 622.9K (82.9K-1.7M) | 760.5K (96.1K-2.1M) | 836.7K (102.5K-2.3M) | 978.1K (111.9K-2.7M) |

***Table G.7. Cumulative economic burden of Lassa-X averted due to vaccine with 70% efficacy.*** *Columns represent the vaccination scenarios considered and rows represent the outcomes averted over approximately two years from the initiation of vaccine rollout. All figures represent means (95% uncertainty intervals) across 100 runs of the transmission model and 100 runs of the health-economic model simulations, for the baseline scenario assuming a probability of sequelae of 62% among patients discharged from hospital, and a probability of hospitalisation ten times greater than for LASV. Monetary costs are reported in International dollars (2021) and future costs are discounted at 3%/year. Catastrophic and impoverishing expenditures are reported as the number of individuals facing those costs. DALY = disability-adjusted life-year, VSL = value of statistical life, VE = vaccine efficacy, K = thousand, M = million, B = billion.*

| ***Outcome averted due to vaccination*** | **Vaccination scenario** | | | | | |
|---|---|---|---|---|---|---|
| | 2.5% of population vaccinated/year | | 20% of population vaccinated/year | | 40% of population vaccinated/year | |
| | 160d delay to first dose | 100d delay to first dose | 160d delay to first dose | 100d delay to first dose | 160d delay to first dose | 100d delay to first dose |
| **Vaccine 70% effective only against disease** ($VE_{infect} = 0\%, VE_{disease} = 70\%$) | | | | | | |
| *Treatment costs, government-reimbursed ($ 2021)* | 2.5M (436.9K-5.7M) | 3.0M (542.6K-6.9M) | 19.7M (3.5M-45.8M) | 24.0M (4.3M-55.0M) | 39.4M (7.0M-91.6M) | 48.1M (8.7M-110.2M) |
| *Treatment costs, out-of-pocket ($ 2021)* | 1.3M (210.8K-3.0M) | 1.6M (255.7K-3.7M) | 10.4M (1.7M-24.2M) | 12.8M (2.0M-29.4M) | 20.8M (3.4M-48.4M) | 25.7M (4.1M-58.9M) |
| *Catastrophic expenditures (N)* | 1.7K (288.3-3.8K) | 2.0K (352.5-4.6K) | 13.3K (2.3K-30.6K) | 16.3K (2.8K-36.7K) | 26.7K (4.6K-61.1K) | 32.7K (5.6K-73.7K) |
| *Impoverishing expenditures (N)* | 1.2K (206.2-2.7K) | 1.4K (255.7-3.2K) | 9.3K (1.6K-21.3K) | 11.4K (2.0K-25.3K) | 18.7K (3.3K-42.7K) | 22.9K (4.1K-50.5K) |
| *Productivity losses ($ 2021)* | 8.1M (948.1K-24.6M) | 10.0M (1.2M-30.1M) | 64.9M (7.6M-196.9M) | 79.9M (9.3M-240.6M) | 129.8M (15.2M-393.7M) | 159.9M (18.6M-481.7M) |
| *Monetised DALYs ($ 2021)* | 2.2M (325.8K-6.6M) | 2.7M (398.8K-7.9M) | 17.9M (2.6M-52.8M) | 21.8M (3.2M-63.2M) | 35.8M (5.3M-105.8M) | 43.6M (6.4M-126.5M) |
| *VSL ($ 2021)* | 109.8M (11.3M-346.3M) | 135.7M (13.8M-420.9M) | 878.2M (90.6M-2.8B) | 1.1B (110.3M-3.4B) | 1.8B (181.6M-5.5B) | 2.2B (221.7M-6.7B) |
| **Vaccine 70% effective against infection and disease** ($VE_{infect} = 70\%, VE_{disease} = 70\%$) | | | | | | |
| *Treatment costs, government-reimbursed ($ 2021)* | 18.7M (3.5M-46.5M) | 24.1M (4.6M-57.2M) | 96.2M (17.2M-222.1M) | 118.3M (21.6M-268.2M) | 134.7M (23.4M-314.4M) | 159.0M (25.3M-370.3M) |
| *Treatment costs, out-of-pocket ($ 2021)* | 9.6M (1.7M-24.3M) | 12.6M (2.3M-30.0M) | 50.5M (8.2M-119.2M) | 63.0M (9.4M-147.6M) | 71.7M (10.1M-172.1M) | 86.1M (10.8M-206.5M) |
| *Catastrophic expenditures (N)* | 12.5K (2.3K-30.7K) | 16.3K (3.0K-38.2K) | 64.9K (11.3K-148.8K) | 80.3K (13.9K-182.1K) | 91.4K (14.6K-210.5K) | 108.7K (16.0K-247.7K) |
| *Impoverishing expenditures (N)* | 8.8K (1.7K-21.9K) | 11.4K (2.2K-26.9K) | 45.5K (8.1K-105.0K) | 56.2K (10.0K-126.9K) | 63.8K (10.5K-147.3K) | 75.9K (11.6K-173.4K) |
| *Productivity losses ($ 2021)* | 60.4M (7.9M-186.4M) | 79.1M (10.4M-238.7M) | 317.7M (36.8M-991.6M) | 395.6M (41.1M-1.2B) | 449.7M (43.7M-1.4B) | 537.8M (47.1M-1.7B) |
| *Monetised DALYs ($ 2021)* | 17.0M (2.6M-51.6M) | 21.9M (3.5M-64.4M) | 88.1M (12.2M-267.6M) | 107.9M (13.9M-321.8M) | 123.2M (14.9M-374.0M) | 144.5M (16.1M-429.3M) |

| | | | | | | |
|---|---|---|---|---|---|---|
| *VSL ($ 2021)* | 809.8M (95.2M-2.6B) | 1.1B (126.4M-3.3B) | 4.3B (423.4M-13.8B) | 5.4B (482.0M-16.9B) | 6.1B (518.6M-19.7B) | 7.3B (557.3M-23.5B) |

***Table G.8. Cumulative economic burden of Lassa-X averted due to vaccine with 90% efficacy.*** *Columns represent the vaccination scenarios considered and rows represent the outcomes averted over approximately two years from the initiation of vaccine rollout. All figures represent means (95% uncertainty intervals) across 100 runs of the transmission model and 100 runs of the health-economic model simulations, for the baseline scenario assuming a probability of sequelae of 62% among patients discharged from hospital, and a probability of hospitalisation ten times greater than for LASV. Monetary costs are reported in International dollars (2021) and future costs are discounted at 3%/year. Catastrophic and impoverishing expenditures are reported as the number of individuals facing those costs. DALY = disability-adjusted life-year, VSL = value of statistical life, VE = vaccine efficacy, K = thousand, M = million, B = billion.*

| ***Outcome averted due to vaccination*** | **Vaccination scenario** | | | | | |
|---|---|---|---|---|---|---|
| | 2.5% of population vaccinated/year | | 20% of population vaccinated/year | | 40% of population vaccinated/year | |
| | 160d delay to first dose | 100d delay to first dose | 160d delay to first dose | 100d delay to first dose | 160d delay to first dose | 100d delay to first dose |
| **Vaccine 90% effective only against disease** ($VE_{infect} = 0\%, VE_{disease} = 90\%$) | | | | | | |
| *Treatment costs, government-reimbursed ($ 2021)* | 3.2M (561.7K-7.4M) | 3.9M (697.7K-8.8M) | 25.3M (4.5M-58.9M) | 30.9M (5.6M-70.7M) | 50.6M (9.0M-117.7M) | 61.8M (11.2M-141.4M) |
| *Treatment costs, out-of-pocket ($ 2021)* | 1.7M (271.1K-3.9M) | 2.1M (328.9K-4.7M) | 13.4M (2.2M-31.1M) | 16.5M (2.6M-37.8M) | 26.7M (4.3M-62.2M) | 33.0M (5.3M-75.6M) |
| *Catastrophic expenditures (N)* | 2.1K (370.5-4.9K) | 2.6K (453.1-5.9K) | 17.1K (3.0K-39.3K) | 21.0K (3.6K-47.2K) | 34.3K (5.9K-78.6K) | 42.0K (7.2K-94.5K) |
| *Impoverishing expenditures (N)* | 1.5K (265.1-3.4K) | 1.8K (328.8-4.1K) | 12.0K (2.1K-27.4K) | 14.7K (2.6K-32.5K) | 24.0K (4.2K-54.8K) | 29.4K (5.3K-64.9K) |
| *Productivity losses ($ 2021)* | 10.4M (1.2M-31.6M) | 12.8M (1.5M-38.7M) | 83.4M (9.7M-253.2M) | 102.7M (11.9M-309.3M) | 166.8M (19.5M-506.1M) | 205.5M (23.9M-618.4M) |
| *Monetised DALYs ($ 2021)* | 2.9M (418.7K-8.5M) | 3.5M (512.4K-10.2M) | 23.0M (3.3M-67.8M) | 28.0M (4.1M-81.3M) | 46.0M (6.7M-135.6M) | 56.0M (8.2M-162.6M) |
| *VSL ($ 2021)* | 141.2M (14.6M-445.2M) | 174.4M (17.7M-541.2M) | 1.1B (116.5M-3.6B) | 1.4B (141.8M-4.3B) | 2.3B (233.1M-7.1B) | 2.8B (283.6M-8.7B) |
| **Vaccine 90% effective against infection and disease** ($VE_{infect} = 90\%, VE_{disease} = 90\%$) | | | | | | |
| *Treatment costs, government-reimbursed ($ 2021)* | 23.1M (4.3M-57.7M) | 29.8M (5.7M-70.5M) | 108.8M (19.5M-251.2M) | 132.3M (23.4M-304.4M) | 145.5M (24.3M-340.9M) | 169.5M (25.8M-397.2M) |
| *Treatment costs, out-of-pocket ($ 2021)* | 11.9M (2.1M-30.0M) | 15.6M (2.8M-37.1M) | 57.3M (8.9M-135.4M) | 70.9M (10.1M-168.3M) | 77.9M (10.6M-189.7M) | 92.3M (11.1M-224.1M) |
| *Catastrophic expenditures (N)* | 15.5K (2.9K-38.2K) | 20.1K (3.7K-47.2K) | 73.5K (12.6K-169.8K) | 90.0K (14.5K-204.2K) | 99.0K (15.2K-229.0K) | 116.1K (16.3K-265.8K) |
| *Impoverishing expenditures (N)* | 10.9K (2.0K-27.0K) | 14.1K (2.7K-33.3K) | 51.4K (9.1K-119.3K) | 62.9K (10.4K-142.8K) | 69.1K (11.0K-160.4K) | 81.0K (11.8K-186.1K) |
| *Productivity losses ($ 2021)* | 74.7M (9.7M-230.5M) | 97.7M (12.6M-296.0M) | 360.4M (39.5M-1.1B) | 444.0M (43.2M-1.4B) | 487.7M (45.5M-1.5B) | 575.6M (48.7M-1.8B) |
| *Monetised DALYs ($ 2021)* | 21.0M (3.2M-63.9M) | 27.1M (4.3M-79.9M) | 99.6M (13.2M-301.9M) | 120.6M (14.7M-362.2M) | 132.9M (15.7M-405.2M) | 153.7M (16.5M-449.5M) |

| | | | | | | |
|---|---|---|---|---|---|---|
| *VSL ($ 2021)* | 1.0B (115.6M-3.2B) | 1.3B (154.2M-4.0B) | 4.9B (452.1M-15.7B) | 6.0B (506.8M-19.2B) | 6.6B (537.7M-21.5B) | 7.9B (567.5M-25.4B) |

#### Appendix H. GATHER checklist

**Table H.1**. GATHER checklist.[55]

| Item | Checklist item | Reporting |
|---|---|---|
| **Objectives and funding** | | |
| 1 | Define the indicator(s), populations (including age, sex, and geographic entities), and time period(s) for which estimates were made. | Results ¶ 1-2, Methods ¶ 8, Supplementary appendix D ¶ 1 |
| 2 | List the funding sources for the work. | *Role of the funder* statement |
| **Data inputs** | | |
| *For all data inputs from multiple sources that are synthesized as part of the study:* | | |
| 3 | Describe how the data were identified and how the data were accessed. | Supplementary appendices B, C.1 and D.3 |
| 4 | Specify the inclusion and exclusion criteria. Identify all ad-hoc exclusions. | Supplementary appendix D.3 |
| 5 | Provide information on all included data sources and their main characteristics. For each data source used, report reference information or contact name/institution, population represented, data collection method, year(s) of data collection, sex and age range, diagnostic criteria or measurement method, and sample size, as relevant. | Supplementary appendix D.3 |
| 6 | Identify and describe any categories of input data that have potentially important biases (e.g., based on characteristics listed in item 5). | Supplementary appendix D.3 |
| *For data inputs that contribute to the analysis but were not synthesized as part of the study:* | | |
| 7 | Describe and give sources for any other data inputs. | Methods ¶ 1,4,7 and Supplementary appendices F.1 and F.2 |
| *For all data inputs:* | | |
| 8 | Provide all data inputs in a file format from which data can be efficiently extracted (e.g., a spreadsheet rather than a PDF), including all relevant meta-data listed in item 5. For any data inputs that cannot be shared because of ethical or legal reasons, such as third-party ownership, provide a contact name or the name of the institution that retains the right to the data. | *Data sharing* statement |
| **Data analysis** | | |
| 9 | Provide a conceptual overview of the data analysis method. A diagram may be helpful. | Results ¶ 1-2, Extended Data Figure 1 |
| 10 | Provide a detailed description of all steps of the analysis, including mathematical formulae. This description should cover, as relevant, data cleaning, data pre-processing, data adjustments and weighting of data sources, and mathematical or statistical model(s). | Supplementary appendices B, C.1, C.3, D.1 and D.2 |
| 11 | Describe how candidate models were evaluated and how the final model(s) were selected. | Supplementary appendix D.4 |
| 12 | Provide the results of an evaluation of model performance, if done, as well as the results of any relevant sensitivity analysis. | Supplementary appendix D.4 |
| 13 | Describe methods for calculating uncertainty of the estimates. State which sources of uncertainty were, and were not, accounted for in the uncertainty analysis. | Methods ¶ 11, Supplementary appendix D.4 |
| 14 | State how analytic or statistical source code used to generate estimates can be accessed. | *Data sharing* statement |
| **Results and Discussion** | | |
| 15 | Provide published estimates in a file format from which data can be efficiently extracted. | Results tables |
| 16 | Report a quantitative measure of the uncertainty of the estimates (e.g. uncertainty intervals). | Reported throughout |
| 17 | Interpret results in light of existing evidence. If updating a previous set of estimates, describe the reasons for changes in estimates. | Discussion ¶ 1,3 |
| 18 | Discuss limitations of the estimates. Include a discussion of any modelling assumptions or data limitations that affect interpretation of the estimates. | Discussion ¶ 6,7 |

#### Appendix I. Appendix references


1. Basinski AJ, Fichet-Calvet E, Sjodin AR, et al. Bridging the gap: Using reservoir ecology and human serosurveys to estimate Lassa virus spillover in West Africa. *PLoS Comput Biol* 2021; **17**(3): e1008811.
2. Global Administrative Areas. GADM database of Global Administrative Areas, version 4.1. 2022. https://gadm.org/data.html (accessed 20/02/2023.
3. Attfield LA. Mathematical modelling of the environmental and ecological drivers of zoonotic disease with an application to Lassa fever: Imperial College London; 2022.
4. World Bank. Birth rate, crude (per 1,000 people). 2022. https://data.worldbank.org/indicator/SP.DYN.CBRT.IN (accessed 03/04/2022.
5. World Bank. Death rate, crude (per 1,000 people). 2022. https://data.worldbank.org/indicator/SP.DYN.CDRT.IN (accessed 03/04/2022.
6. World Bank. Population, total. 2022. https://data.worldbank.org/indicator/SP.POP.TOTL (accessed 03/04/2022.
7. Lerch A, Ten Bosch QA, L'Azou Jackson M, et al. Projecting vaccine demand and impact for emerging zoonotic pathogens. *BMC Med* 2022; **20**(1): 202.
8. Lerch A. VaccineCampaign. 2022. https://github.com/lerch-a/VaccineCampaign (accessed 16/10/2023.
9. US CDC. Lassa Fever Outbreak Distribution Map. 2014. https://www.cdc.gov/vhf/lassa/outbreaks/index.html (accessed 16/10/2023.
10. World Health Organization. Geographic distribution of Lassa fever in West African affected countries, 1969-2018. 2018. https://cdn.who.int/media/images/default-source/health-topics/lassa-fever/lassa-fever-countries-2018png.tmb-1024v.png?sfvrsn=10af107d_7 (accessed 16/10/2023.
11. CEPI. 2021 Annual Progress Report. Oslo, Norway, 2022.
12. World Health Organization. Oral Cholera Vaccine stockpile for cholera emergency response, 2013.
13. Massing LA, Aboubakar S, Blake A, et al. Highly targeted cholera vaccination campaigns in urban setting are feasible: The experience in Kalemie, Democratic Republic of Congo. *PLoS Negl Trop Dis* 2018; **12**(5): e0006369.
14. Ngwa MC, Alemu W, Okudo I, et al. The reactive vaccination campaign against cholera emergency in camps for internally displaced persons, Borno, Nigeria, 2017: a two-stage cluster survey. *BMJ Glob Health* 2020; **5**(6).
15. World Health Organization. Oral cholera vaccines in mass immunization campaigns: guidance for planning and use. Geneva, Switzerland, 2010.
16. Ali M, Nelson AR, Lopez AL, Sack DA. Updated global burden of cholera in endemic countries. *PLoS Negl Trop Dis* 2015; **9**(6): e0003832.
17. World Health Organization. Disease Outbreak News; Cholera - Global Situation. 11/02/2023 2023. https://www.who.int/emergencies/disease-outbreak-news/item/2023-DON437 (accessed 20/02/2023.
18. WorldPop. WorldPop. 2023. https://www.worldpop.org/ (accessed 16/10/2023.
19. World development report 1993--investing in health. *Commun Dis Rep CDR Wkly* 1993; **3**(30): 137.
20. Larson BA. Calculating disability-adjusted-life-years lost (DALYs) in discrete-time. *Cost Eff Resour Alloc* 2013; **11**(1): 18.
21. McCormick JB, Webb PA, Krebs JW, Johnson KM, Smith ES. A prospective study of the epidemiology and ecology of Lassa fever. *J Infect Dis* 1987; **155**(3): 437-44.

22. Penfold S, Adegnika AA, Asogun D, et al. A prospective, multi-site, cohort study to estimate incidence of infection and disease due to Lassa fever virus in West African countries (the Enable Lassa research programme)-Study protocol. *PLoS One* 2023; **18**(3): e0283643.
23. Battle KE, Bisanzio D, Gibson HS, et al. Treatment-seeking rates in malaria endemic countries. *Malar J* 2016; **15**: 20.
24. Naidoo D, Ihekweazu C. Nigeria's efforts to strengthen laboratory diagnostics - Why access to reliable and affordable diagnostics is key to building resilient laboratory systems. *Afr J Lab Med* 2020; **9**(2): 1019.
25. Saleh F, Popoola BO, Arinze C, et al. Adapting public health response through lessons learnt: Nigeria's experience from Lassa fever and COVID-19. *BMJ Glob Health* 2022; **7**(Suppl 7).
26. Simons D. Lassa fever cases suffer from severe underreporting based on reported fatalities. *Int Health* 2023; **15**(5): 608-10.
27. Camacho A, Ndiaye A, Amevoin Y, Mousset M. Enable interim analysis: preliminary results (data as of 24 October 2022), 2022.
28. Duvignaud A, Jaspard M, Etafo IC, et al. Lassa fever outcomes and prognostic factors in Nigeria (LASCOPE): a prospective cohort study. *Lancet Glob Health* 2021; **9**(4): e469-e78.
29. Ficenec SC, Percak J, Arguello S, et al. Lassa Fever Induced Hearing Loss: The Neglected Disability of Hemorrhagic Fever. *Int J Infect Dis* 2020; **100**: 82-7.
30. Bria YP, Yeh CH, Bedingfield S. Significant symptoms and nonsymptom-related factors for malaria diagnosis in endemic regions of Indonesia. *Int J Infect Dis* 2021; **103**: 194-200.
31. Wan X, Wang W, Liu J, Tong T. Estimating the sample mean and standard deviation from the sample size, median, range and/or interquartile range. *BMC Med Res Methodol* 2014; **14**: 135.
32. Global Burden of Disease Collaborative Network. Global Burden of Disease Study 2019 (GBD 2019) Disability Weights. In: (IHME) IfHMaE, editor. Seattle, United States of America; 2020.
33. Merson L, Bourner J, Jalloh S, et al. Clinical characterization of Lassa fever: A systematic review of clinical reports and research to inform clinical trial design. *PLoS Negl Trop Dis* 2021; **15**(9): e0009788.
34. Haagsma JA, Maertens de Noordhout C, Polinder S, et al. Assessing disability weights based on the responses of 30,660 people from four European countries. *Popul Health Metr* 2015; **13**: 10.
35. Ginsberg GM, Kark JD, Einav S. Cost-utility analysis of treating out of hospital cardiac arrests in Jerusalem. *Resuscitation* 2015; **86**: 54-61.
36. John D, Narassima MS, Bhattacharya P, Mukherjee N, Banerjee A, Menon J. Model-based estimation of burden of COVID-19 with disability-adjusted life years and value of statistical life in West Bengal, India. *BMJ Open* 2023; **13**(1): e065729.
37. Global Burden of Disease Hearing Loss Collaborators. Hearing loss prevalence and years lived with disability, 1990-2019: findings from the Global Burden of Disease Study 2019. *Lancet* 2021; **397**(10278): 996-1009.
38. Ochalek J, Lomas J, Claxton K. Estimating health opportunity costs in low-income and middle-income countries: a novel approach and evidence from cross-country data. *BMJ Glob Health* 2018; **3**(6): e000964.
39. Moses MW, Pedroza P, Baral R, et al. Funding and services needed to achieve universal health coverage: applications of global, regional, and national estimates of utilisation of outpatient visits and inpatient admissions from 1990 to 2016, and unit costs from 1995 to 2016. *Lancet Public Health* 2019; **4**(1): e49-e73.

40. The World Bank. World Bank Open Data. 2023. https://databank.worldbank.org/ (accessed 16/10/2023.
41. Asogun D, Tobin E, Momoh J, et al. Medical cost of Lassa fever treatment in Irrua Specialist Teaching Hospital, Nigeria. *Int J Basic Appl Innov Res* 2016; **5**(3): 62-73.
42. World Health Organization. The Global Health Observatory. SDG 3.8.2 Catastrophic health spending (and related indicators). 2023. https://www.who.int/data/gho/data/themes/topics/topic-details/GHO/financial-protection (accessed 16/10/2023.
43. The World Bank. Poverty and Inequality Platform. 2023. https://pip.worldbank.org/home (accessed 16/10/2023.
44. International Labour Organization. International Labour Organization Statistics ILOSTAT. 2023. https://ilostat.ilo.org/ (accessed 16/10/2023.
45. Colmer J. What is the meaning of (statistical) life? Benefit-cost analysis in the time of COVID-19. *Oxford Rev Econ Pol* 2020; **36**: S56-S63.
46. Robinson LA, Hammitt JK, Cecchini M, et al. Reference Case Guidelines for Benefit-Cost Analysis in Global Health and Development, 2019.
47. Assistant Secretary for Planning and Evaluation (ASPE). Appendix D: Updating Value per Statistical Life (VSL) Estimates for Inflation and Changes in Real Income. 2021. https://aspe.hhs.gov/reports/updating-vsl-estimates (accessed 16/10/2023.
48. Kramer AM, Pulliam JT, Alexander LW, Park AW, Rohani P, Drake JM. Spatial spread of the West Africa Ebola epidemic. *R Soc Open Sci* 2016; **3**(8): 160294.
49. Garske T, Cori A, Ariyarajah A, et al. Heterogeneities in the case fatality ratio in the West African Ebola outbreak 2013-2016. *Philos Trans R Soc Lond B Biol Sci* 2017; **372**(1721).
50. Dudas G, Carvalho LM, Bedford T, et al. Virus genomes reveal factors that spread and sustained the Ebola epidemic. *Nature* 2017; **544**(7650): 309-15.
51. GeoNames. GeoNames. 2023. https://www.geonames.org/ (accessed 17/06/2023.
52. Thompson RN, Stockwin JE, van Gaalen RD, et al. Improved inference of time-varying reproduction numbers during infectious disease outbreaks. *Epidemics* 2019; **29**: 100356.
53. W. H. O. Ebola Response Team, Aylward B, Barboza P, et al. Ebola virus disease in West Africa--the first 9 months of the epidemic and forward projections. *N Engl J Med* 2014; **371**(16): 1481-95.
54. Soetaert K, Petzoldt T, Setzer RW. Solving Differential Equations in R: Package deSolve. *J Stat Softw* 2010; **33**(9): 1-25.
55. Stevens GA, Alkema L, Black RE, et al. Guidelines for Accurate and Transparent Health Estimates Reporting: the GATHER statement. *Lancet* 2016; **388**(10062): e19-e23.
